# An umbrella review and meta-analysis of the use of renin-angiotensin system drugs and COVID-19 outcomes: what do we know so far?

**DOI:** 10.1101/2022.03.20.22272664

**Authors:** Amanj Kurdi, Natalie Weir, Tanja Mueller

**Author notes:** **Corresponding author:** Amanj Kurdi, Strathclyde Institute of Pharmacy and Biomedical Science, University of Strathclyde, Glasgow, 161 Cathedral Street, G4 0RE. Funding sources/sponsors: None.

## Abstract

**Backgrounds:** Evidence from several meta-analyses are still controversial about the effects of angiotensin-converting enzyme inhibitors (ACEIs)/angiotensin-receptor blockers (ARBs) on COVID-19 outcomes.

**Purpose:** Umbrella review of systematic reviews/meta-analysis to provide comprehensive assessment of the effect of ACEIs/ARBs on COVID-19 related outcomes by summarising the currently available evidence.

**Data Source:** Medline (OVID), Embase, Scopus, Cochrane library and medRxiv from inception to 1^st^ February 2021.

**Study Selection:** Systematic reviews with meta-analysis that evaluated the effect of ACEIs/ARBs on COVID-19 related clinical outcomes

**Data Extraction:** Two reviewers independently extracted the data and assessed studies’ risk of bias using AMSTAR 2 Critical Appraisal Tool.

**Data Synthesis:** Pooled estimates were combined using the random-effects meta-analyses model including several sub-group analyses. Overall, 47 reviews were eligible for inclusion. Out of the nine COVID-19 outcomes evaluated, there was significant associations between ACEIs/ARBs use and each of death (OR=0.80, 95%CI=0.75-0.86; I^2^=51.9%), death/ICU admission as composite outcome (OR=0.86, 95%CI=0.80-0.92; I^2^=43.9%), severe COVID-19 (OR=0.86, 95%CI=0.78-0.95; I^2^=68%), and hospitalisation (OR=1.23, 95%CI=1.04-1.46; I^2^= 76.4%). The significant reduction in death/ICU admission, however, was higher among studies which presented adjusted measure of effects (OR=0.63, 95%CI=0.47-0.84) and were of moderate quality (OR=0.74, 95%CI=0.63-0.85).

**Limitations:** The effect of unmeasured confounding could not be ruled out. Only 21.3% (n=10) of the studies were of ‘moderate’ quality.

**Conclusion:** Collective evidence from observational studies indicate a good quality evidence on the significant association between ACEIs/ARBs use and reduction in death and death/ICU admission, but poor-quality evidence on both reducing severe COVID-19 and increasing hospitalisation. Our findings further support the current recommendations of not discontinuing ACEIs/ARBs therapy in patients with COVID-19.

**Registration:** The study protocol was registered in PROSPERO (CRD42021233398).

**Funding Source:** None

## Introduction

A new coronavirus variant, the “severe acute respiratory syndrome coronavirus 2” (SARS-CoV-2), first emerged in Wuhan, China, in late 2019, and has since spread globally. The disease caused by this virus is now commonly known as COVID-19 and presents with a range of symptoms, including fever and a persistent cough; in severe cases, patients require hospitalisation and ventilation.

Several risk factors linked to poor disease outcomes have been identified early on, including age, sex, and the presence of certain conditions such as cardiovascular disease, including hypertension (1). Consequently, the possible impact of renin-angiotensin-aldosterone system (RAAS) inhibitors on COVID-19 related outcomes has emerged as a topic of interest, based on their widespread use among patients at risk of poor disease outcomes (2) and their mechanisms of action – in particular, the potential upregulation of angiotensin-converting enzyme 2 (ACE2) which is associated with viral entry into bronchial cells (3).) This has resulted in the rapid dissemination of numerous studies, mostly retrospective observational in nature, focusing on the risk of COVID-19 infection, disease severity, and/or disease outcomes in patients being treated with either angiotensin-converting-enzyme inhibitors (ACEIs) or angiotensin receptor blockers (ARBs) since early 2020 (4-6).

As was the case in most early COVID-19 related research, the evidence comprised observational studies with notably small sample sizes and short durations of follow-up. Resultantly, to draw ultimate conclusions, a number of systematic reviews were swiftly published in attempt to offer a more substantial view by aggregating findings of these small-scale studies. These meta-analyses have offered tentative insights into all three areas of interest with regards to the use of RAAS inhibitors in times of COVID-19: (i) risk of infection, usually measured as the share of positive PCR tests within a study cohort; (ii) risk of severe COVID-19, with various underlying definitions ranging from hospitalisation due to the disease to the requirement for mechanical ventilation; and (iii) the risk of mortality, identified by using recorded cause of death during hospitalisation. While there were similarities between some of the published results – e.g. indicating, in general, no association between RAAS inhibitor use and risk of COVID-19 infection – other results were more varied (4-6).

There are many possible reasons for the variability in the published results based on conducted meta-analyses, the main one potentially being the varying timeframes of the underlying systematic reviews and, therefore, the diverging inclusion of potentially relevant studies. Although each publication, in and off itself, may offer some interesting information, the observed discrepancies between previously published findings and, even more so, the limited number of primary studies and – consequently – the limited number of patients and events of interest included in each of them results in an overall impression of inconclusiveness, warranting further scrutiny.

A logical next step, besides conducting additional systematic reviews/meta-analyses, is to perform a systematic review of systematic reviews (also known as umbrella review), thereby taking advantage of the availability of high-level evidence and providing an opportunity to contrast and compare (7). The aim of this umbrella review and meta-analysis, therefore, was to assess the effect of ACEIs and ARBs on COVID-19 related outcomes by summarising the currently available, aggregate evidence.

## Methods

An umbrella literature review and subsequent meta-analysis was conducted. The protocol was informed by Joanna Briggs Reviewer’s Manual for ‘Development of an Umbrella review protocol’ (8) and published on PROSPERO (CRD42021233398).

### Eligibility criteria

Eligible studies were systematic reviews which conducted a meta-analysis to explore the effect of ACEIs and ARBs on COVID-19 related outcomes. Eligible study populations were adults (≥18 years) in real-world contexts with and without COVID-19 diagnosis. The exposure of interest was treatment with RAAS inhibitors (i.e. ACEIs and/or ARBs) compared to those not exposed to RAAS inhibitors. Reviews conducting a comparison between patients exposed to ACEIs and patients exposed to ARBs were also eligible for inclusion. Outcomes of interest were COVID-19 infection risk and COVID-19 related clinical outcome, including but not limited to: death; severity of COVID-19 infection; admission to intensive care unit (ICU); hospitalisation; hospital discharge; ventilator use; length of hospital stay; hospital re-admission; dialysis; acute respiratory distress syndrome; septic shock; acute kidney injury; cardiac injury; pneumonia severity; as well as other relevant outcomes identified iteratively throughout study selection and data extraction.

### Search strategy

The databases Medline, EMBASE, Scopus, Cochrane, and medRxiv were searched in February 2021. Publications were searched from 2019 onwards to reflect the date with which COVID-related reviews could have been published. The search was limited to the English language and for systematic review articles. Search terms for “renin-angiotensin system”, “angiotensin-converting enzyme inhibitors”, “angiotensin II receptor antagonists”, “COVID-19” were used with various synonyms, truncation codes and Boolean operators (Supplementary file **1**). When full texts were not obtainable the author(s) were contacted up to two times to request full texts. The reference lists of included reviews were also screened to identify eligible reviews.

### Article selection

Article selection was conducted using Covidence software (9). To ensure consistency in the study selection process 10% of the articles’ titles/abstracts and full texts were randomly selected and screened independently by two researchers (NW and TM). The percentage of agreement was calculated for all independent validation, with >80% considered adequate (10). Where dubiety arose over an article’s eligibility a third reviewer was consulted (AK).

### Data extraction

Data were extracted from the reviews using Microsoft Excel. A data extraction template was piloted with 10% of reviews by NW and agreed for use by all authors. 10% of reviews were randomly selected and underwent independent data extraction by NW and TM; the percentage of agreement was calculated. Again, agreement >80% was considered adequate (10). Where dubiety arose over data extraction a second reviewer was consulted (AK). Data extracted from the reviews included: title; authors; year review published; study design; sample size; setting; population; exposure (e.g. ACEIs/ARBs, ACEIs, or ARBs); and outcomes (e.g. death, COVID-19 infection, hospitalisation). Data was extracted from the published reviews only; the primary studies were not referred to and authors were not contacted for further data.

### Quality Assessment

Quality assessment was conducted independently by NW and TM using the AMSTAR 2 tool (11).Studies were categorised as having high, moderate, low and critically low confidence in the results based on the number of ‘critical domains’. Critical domains related to each review containing: an explicit statement that the methods were established a priori within a protocol; if a satisfactory technique for assessing the risk of bias (RoB) was conducted and sufficiently discussed; if the meta-analysis used appropriate methods; and if publication bias (small study bias) was conducted.

### Data analysis and synthesis

The random-effects meta-analysis model was used to statistically combine the measure of effects for those outcomes that were reported by more than one study to obtain one pooled estimate for each outcome, stratified by the three level of exposure (ACEIs/ARBs, ACEIs, ARBs). We used random-effects model because it allows the results to be generalisable to other populations as well as addresses the likely heterogeneity between the included studies; hence it is the most commonly used meta-analysis model (12). In order to explore the potential source of heterogeneity as well as the effect of potential confounders on the sensitivity and robustness of the combined pooled estimates, we conducted several sub-group analyses based on numerous variables including: whether the reported measure of effects was crude or adjusted, the study was peer-reviewed or not, and the study’s methodological quality as per the quality assessment. Furthermore, to assess the impact of ACEIs/ARBs among patients with hypertension (the most common indication for ACEIs/ARBs), we also conducted sub-group analysis based on whether the studies had included either patients with hypertension only or at least had hypertension as one of the comorbidities versus those studies which did not recorded the hypertension status of their study population. The combined pooled estimates were presented as odds ratios and 95%CI and graphically as forest plots. I^2^ statistic (13) was used to assess heterogeneity between the studies, to check whether the variability is more likely to be due to chance or heterogeneity in the studies; I^2^ values ranged between 0%-100% with 0% indicating lack of heterogeneity, whereas 25%, 50%, and 75% indicating low, moderate and high heterogeneity, respectively (13). Publication bias was assessed using funnel plots and Egger’s asymmetry test (14) for those outcomes where >10 studies were included in the analysis as recommended by Cochrane guidelines (15). Furthermore, we evaluated the influence of individual reviews on the summary pooled estimate for each outcome by conducting influential analyses (16) whereby the pooled meta-analysis estimates for each outcome were computed by omitting one study at a time. Data were analysed using STATA 12.

## Results

Out of an initial 157 publications, 66 systematic reviews underwent full text screening; after further exclusions based on pre-specified criteria, 47 studies were identified to be relevant for this project (Figure **1**) (4-6, 17-60).

**Figure 1.**
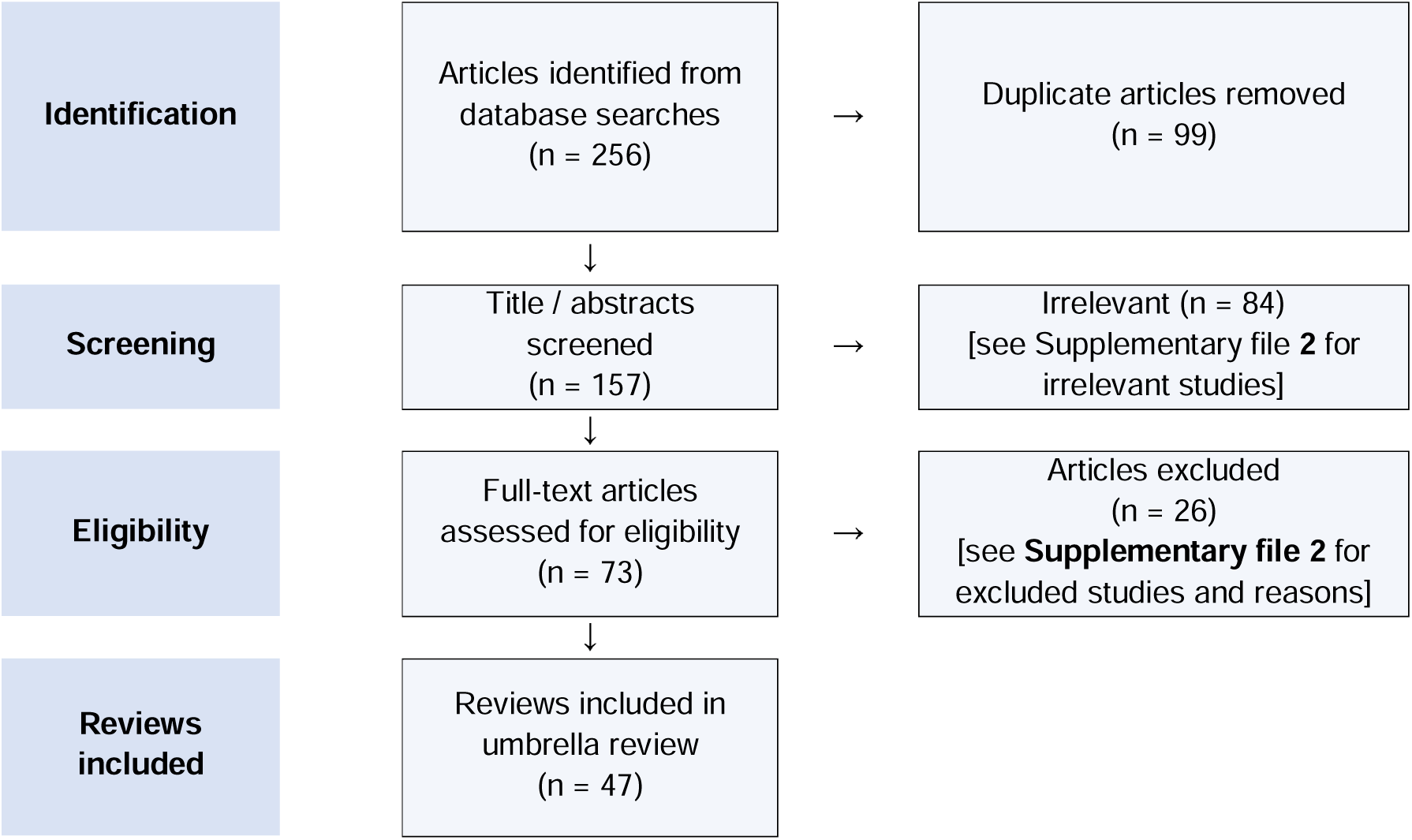
PRISMA flow diagram of the review selection process.

### Review characteristics

Forty-six reviews (97.9%) compared COVID-19 related outcomes between ACEI/ARB users vs. non-users among patients with COVID-19 (4-6, 17-52, 54-60), one study (2.12%%) compared outcomes between ACEIs/ARBs users in patients with and without COVID-19 infection (53)), and 16 studies (34.0%) explored both (6, 19, 25-27, 40, 41, 43, 44, 48, 50, 51, 54, 56, 58, 60). Most of the included reviews were peer-reviewed publications (68.1%; n=32), whereas the remining 15 (31.9%) reviews were non-peer reviewed publications (i.e. were published in a pre-print database) (17-19, 21-23, 30, 32-34, 36, 46, 50, 54, 60). The time the searches were conducted ranged from April 2020 to October 2020, with 21 (44.7%) review searches conducted in the month of May 2020 (4-6, 17, 21, 23, 24, 28, 30-32, 35, 36, 40-42, 44, 46, 48, 50, 54) Pre-print articles were included in 28 (59.6%) reviews (4, 17, 19-22, 25, 26, 30, 33, 37, 41-45, 47-53, 55, 56, 59, 60), and 10 (21.3%) reviews adjusted for retracted studies (4, 18, 31, 40, 45, 47-50, 56). Full details of the 47 reviews are presented in Supplementary file **3**.

A total of 213 meta-analyses were conducted by the 47 reviews (**Supplementary file 4**). In terms of number of COVID-19 related outcomes reported in each review, one outcome was reported by 13 reviews (27.7%) (18, 20, 21, 23, 24, 28, 29, 38, 39, 47, 52, 53, 61), two outcomes by 15 reviews (31.9%) (4, 17, 26, 31, 32, 34-37, 40, 42, 49, 54, 55, 58), three outcomes by 11 reviews (23.4%) (6, 22, 25, 27, 33, 44-46, 50, 56, 60) and 4-9 outcomes by eight reviews (17%) (19, 30, 41, 43, 48, 51, 57, 59). Overall, the 47 eligible reviews reported data on 18 unique pooled outcome estimates including death in 36 reviews, reviews (4, 6, 17-19, 22, 24, 25, 27, 30-39, 41-49, 54-56, 58-60), ICU admission in nine reviews (27, 28, 30, 41, 43, 48, 51, 56, 59), death/ICU admission as a composite outcome in 16 reviews (4, 20, 21, 23, 26, 29, 31, 32, 40, 41, 43, 45, 51, 55, 59), risk of acquiring COVID-19 infection in 15 reviews (19, 25, 27, 40, 41, 43, 44), severe COVID-19 infection in 22 reviews (6, 17, 19, 22, 25, 30, 33-37, 41-46, 48, 59, 60), hospitalisation in nine reviews (19, 30, 41, 43, 48, 59), length of hospital stay in five reviews (19, 22, 30, 46, 59), use of mechanical ventilator in three reviews (30, 41), risk of severe acute respiratory syndrome (SARS) in two reviews (26, 59), and each of hospital discharge (30), ICU admission/mechanical ventilator use (41), risk of COVID-19 infection/hospitalisation (53), severe pneumonia (41), level of serum creatinine (57), d-dimer (57), cough (57), fever (57) and renal dialysis (59) in one review; accordingly, nine out of these 18 outcomes were included in the meta-analysis as they were reported by at least two reviews. In terms of the exposure, ACEIs and ARBs were evaluated as one class (ACEIs/ARBs) in all the eligible 47 reviews but three (26, 53, 57), and as separate classes in 17 (4, 6, 23, 25-27, 30, 31, 38, 40, 41, 43, 47, 50, 53, 54, 58) and 16 (4, 6, 23, 25-27, 30, 31, 38, 40, 41, 43, 50, 53, 54, 58) reviews, respectively. Majority of the reviews (66%; n=31) only evaluated one exposure, mainly ACEIs/ARBs combined as one class (n=30); whereas one third of them (29.8%; n=14) reported data for the three level of exposure (ACEIs/ARBs, ACEIs, ARBs).

### Quality assessment

Overall confidence in the results was ‘moderate’ for 10 (21.3%) reviews (19, 25, 26, 30, 37, 41-43, 56, 59), ‘low’ for 15 (30.6%) reviews (4, 5, 20-22, 27, 28, 31, 34, 45, 49-51, 55, 60), and ‘critically low’ for 22 (44.9%) reviews (6, 17, 18, 23, 24, 29, 32, 33, 35, 36, 38-40, 44, 46-48, 52-54, 57, 58) (Supplementary file **5**). Considering the critical domains, most reviews were considered to have had a satisfactory technique for the statistical combination of results (n=45, 95.7%) (4-6, 17-22, 24-57, 59, 60) and for assessing risk of bias (n=38, 80.1%) (4-6, 17, 19-23, 25-28, 30, 31, 34-38, 40-46, 48-53, 55-57, 59, 60). Less reviews were favourably considered in terms of accounting for risk of bias when interpreting and discussing the results (n=32, 68.1%), with appropriate conduct of publication bias (n=33) (4-6, 17, 19-21, 23-27, 30-33, 37, 38, 41-45, 47, 49-51, 53, 56, 57, 59, 60), and only 15 (31.9%) reviews referred to the review methods being established a priori (19, 22, 25, 26, 28, 30, 34, 37, 41-43, 52, 55, 56, 59).

### Effect of ACEIs/AEBs (as a one group) on the study outcomes

Overall, the effect of ACEIs/ARBs on nine COVID-19 related clinical outcomes were evaluated (**Table 1**). The combined pooled meta-analysis estimates indicated that ACEIs/ARBs used was associated with a significant reduction in three clinical outcomes including death (OR=0.80, 95%CI=0.75-0.86; I^2^ = 51.9%) (Figure **2**) death/ICU admission as composite outcome (OR=0.86, 95%CI= 0.80-0.92; I2= 43.9%) (Figure **3**) and severe COVID-19 infection (OR=0.86, 95% CI=0.78-0.95; I2 = 68%) (Figure **4**); on the other hand, ACEIs/ARBs was associated with a significant increase in hospitalisation (OR=1.23, 95%CI=1.04-1.46; I2= 76.4%) (Figure **5**). However, there was insignificant association with each of ICU admission (Figure **6**), risk of acquiring COVID-19 infection (Figure **7**), use of mechanical ventilator (Figure **8**), risk of SARS (Figure **9**), and risk of severe pneumonia (Figure **10**).

**Table 1.** Meta-analyses pooled estimates with 95%CI of the effects of ACEIs/ARBs on COVID-19 related clinical outcomes.

| Outcomes | ACEIs/ARBs | p-value | ACEIs | p-value | ARBs | P-value |
| --- | --- | --- | --- | --- | --- | --- |
| **Death** | 0.80 (0.75, 0.86) | <0.001 | 0.91 (0.89, 1.12) | 0.984 | 1.10 (0.94, 1.25) | 0.263 |
| Number of studies | 47 |  | 7 |  | 6 |  |
| I-squared | 51.9% | 0.001 | 29.1% | 0.206 | 41.5% | 0.129 |
| **ICU** | 1.03 (0.86, 1.19) | 0.721 | 0.96 (0.87, 1.1) | 0.406 | 1.21 (0.93, 1.47) | 0.312 |
| Number of studies | 10 |  | 4 |  | 4 |  |
| I-squared (p-value) | 58.7% | 0.01 | 0% | 0.882 | 76.5% | 0.005 |
| **Death/ICU** | 0.86 (0.80, 0.92) | <0.001 | 0.94 (0.86, 1.03) | 0.167 | 0.98 (0.92, 1.05) | 0.530 |
| Number of studies | 22 |  | 8 |  | 8 |  |
| I-squared (p-value) | 43.9% | 0.015 | 29.5% | 0.193 | 0% | 0.614 |
| **Risk of COVID-19** | 0.99 (0.97, 1.02) | 0.560 | 0.97 (0.93, 1.01) | 0.058 | 1.01 (0.97, 1.04) | 0.726 |
| Number of studies | 19 |  | 11 |  | 10 |  |
| I-squared (p-value) | 24.7% | 0.159 | 31.7% | 0.146 | 0% | 0.757 |
| **Severe COVID-19** | 0.86 (0.78, 0.95) | 0.003 | 0.92 (0.81, 1.05) | 0.232 | 0.94 (0.84, 1.05) | 0.281 |
| Number of studies | 28 |  | 8 |  | 8 |  |
| I-squared (p-value) | 68% | <0.001 | 0% | 0.951 | 53.7% | 0.580 |
| **Severe pneumonia** | 0.82 (0.22, 3.05) | 0.765 | NA |  | NA |  |
| Number of studies | 2 |  |  |  |  |  |
| I-squared (p-value) | 0% | 0.405 |  |  |  |  |
| **Hospitalisation** | 1.23 (1.04, 1.46) | 0.019 | 1.18 (1.04, 1.35) | 0.012 | 1.17 (0.84, 1.61) | 0.354 |
| Number of studies | 11 |  | 5 |  | 5 |  |
| I-squared (p-value) | 76.4% | <0.001 | 6.7% | 0.368 | 86.9% | <0.001 |
| **Ventilator use** | 1.18 (0.84, 1.66) | 0.347 | 1.01 (0.03, 34.52) | 0.994 | 0.985 (0.084, 11.57) | 0.990 |
| Number of studies | 3 |  | 1 |  | 1 |  |
| I-squared (p-value) | 53.9% | 0.114 | NA |  | NA |  |
| **Acute SARS infection** | 0.71 (0.49, 1.02) | 0.064 | 1.06 (0.84, 1.34) | 0.633 | 1.11 (0.95, 1.29) | 0.493 |
| Number of studies | 1 |  | 2 |  | 2 |  |
| I-squared (p-value) | NA |  | 81% | 0.022 | 48.9% | 0.162 |

(Note) NA: not applicable indicating not enough studies to perform meta-analyses

**Figure 2.**
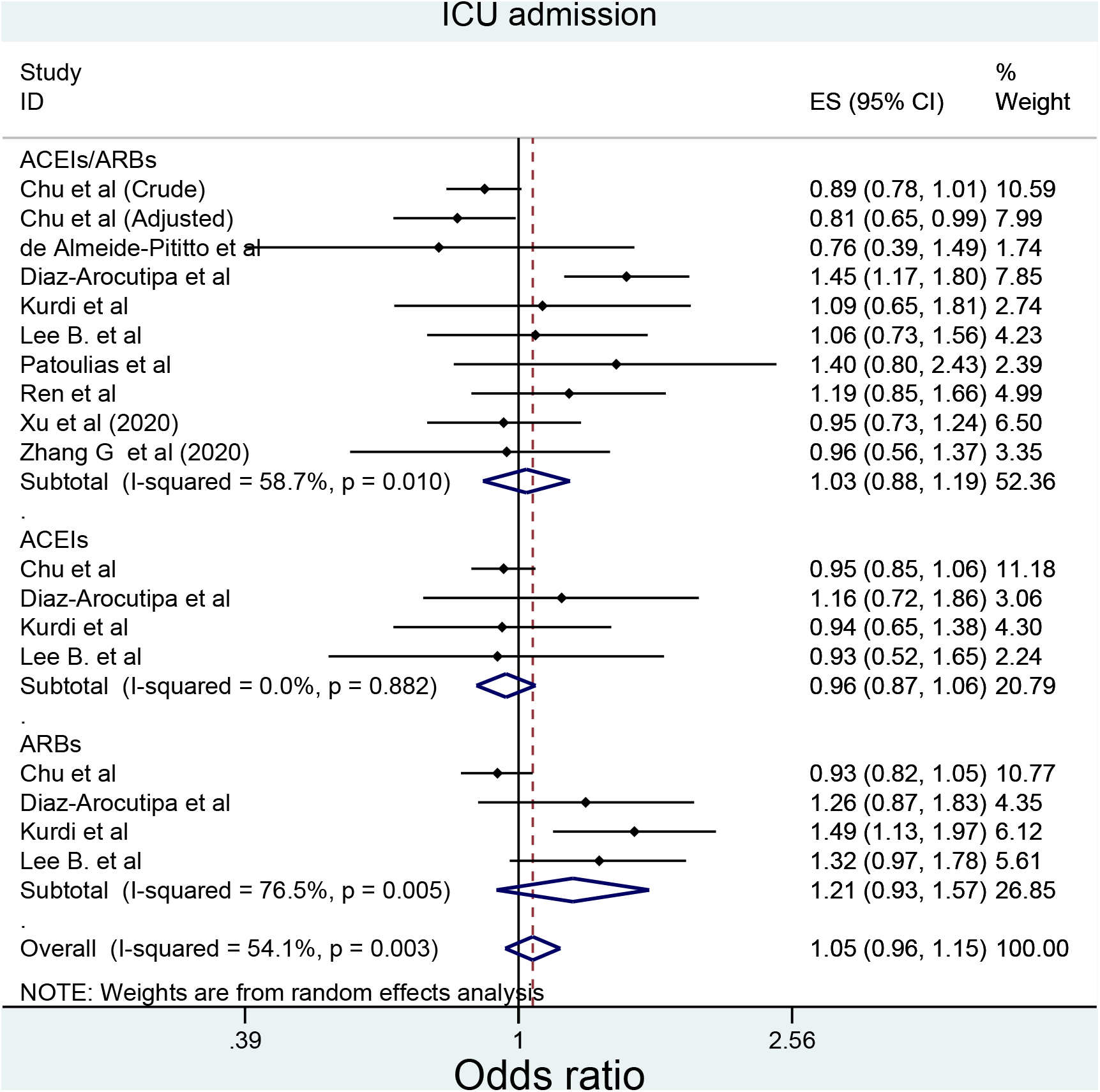
Forest plot depicting pooled estimates for the association between mortality and the three level of renin-angiotensin system drug exposure (ACEIs/ARBs, ACEIs, ARBs)

**Figure 3.**
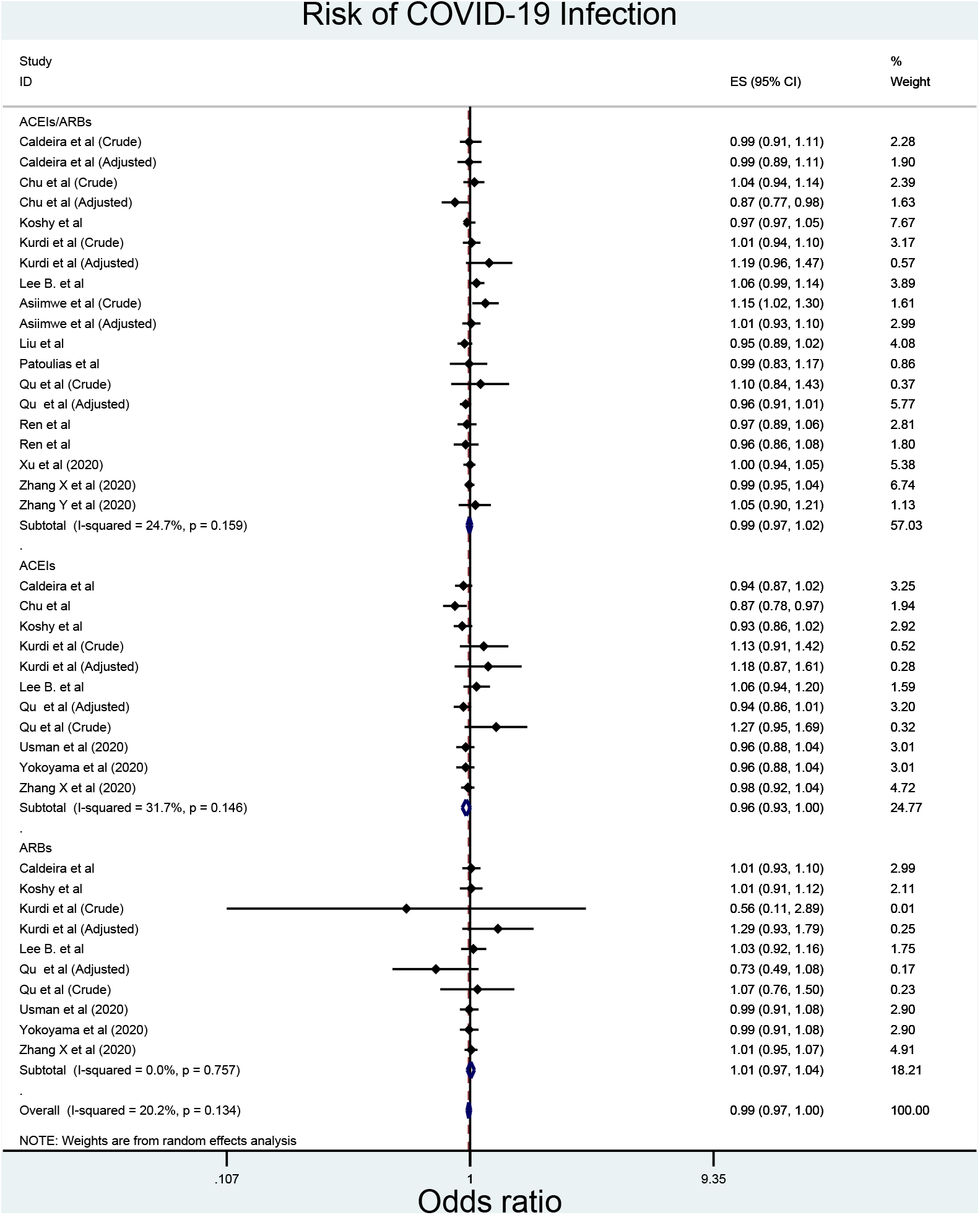
Forest plot depicting pooled estimates for the association between death/Intensive Care Unit (as a composite outcome) and the three level of renin-angiotensin system drug exposure (ACEIs/ARBs, ACEIs, ARBs)

**Figure 4.**
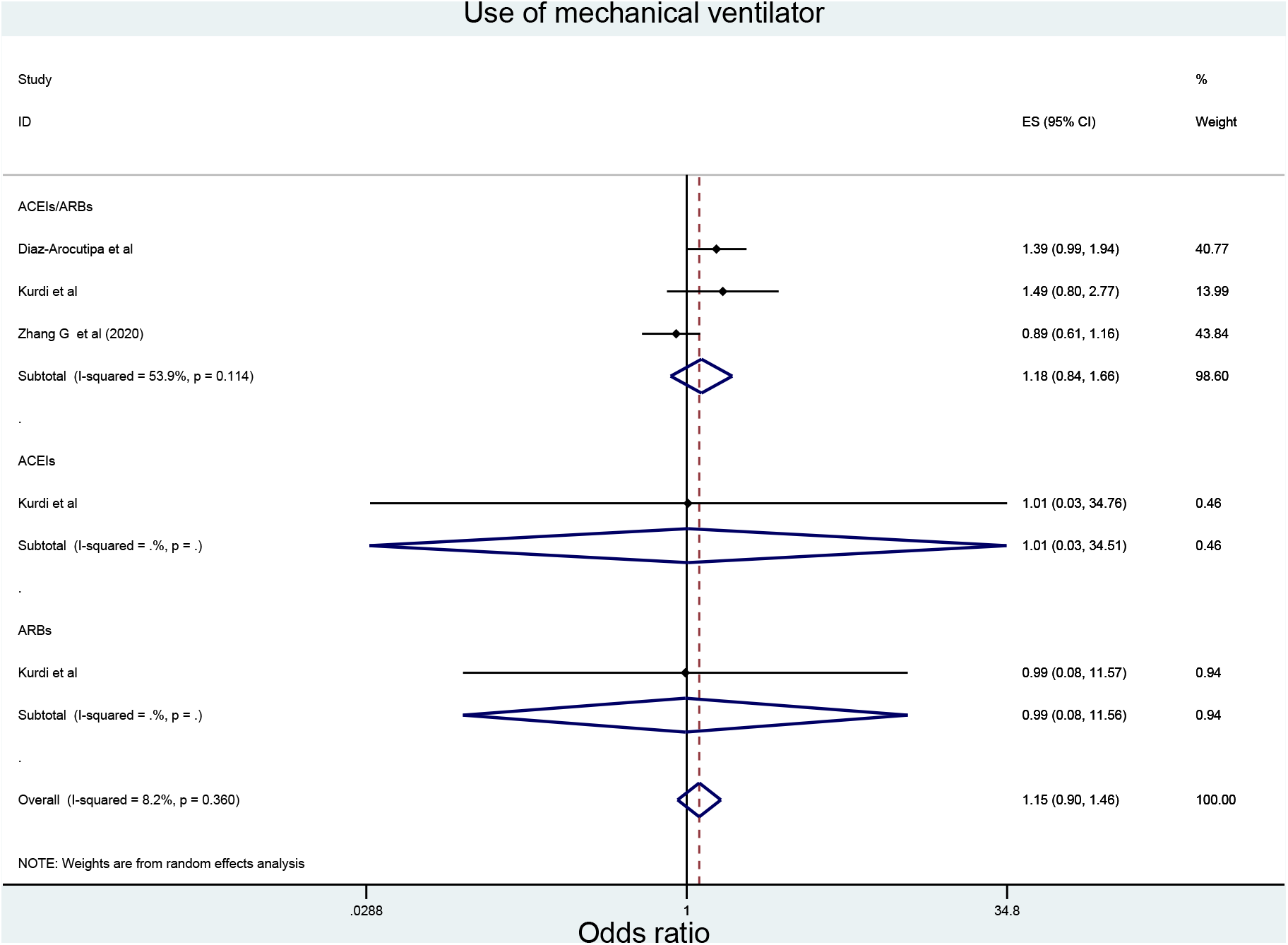
Forest plot depicting pooled estimates for the association between severe COVID-19 infection and the three level of renin-angiotensin system drug exposure (ACEIs/ARBs, ACEIs, ARBs)

**Figure 5.**
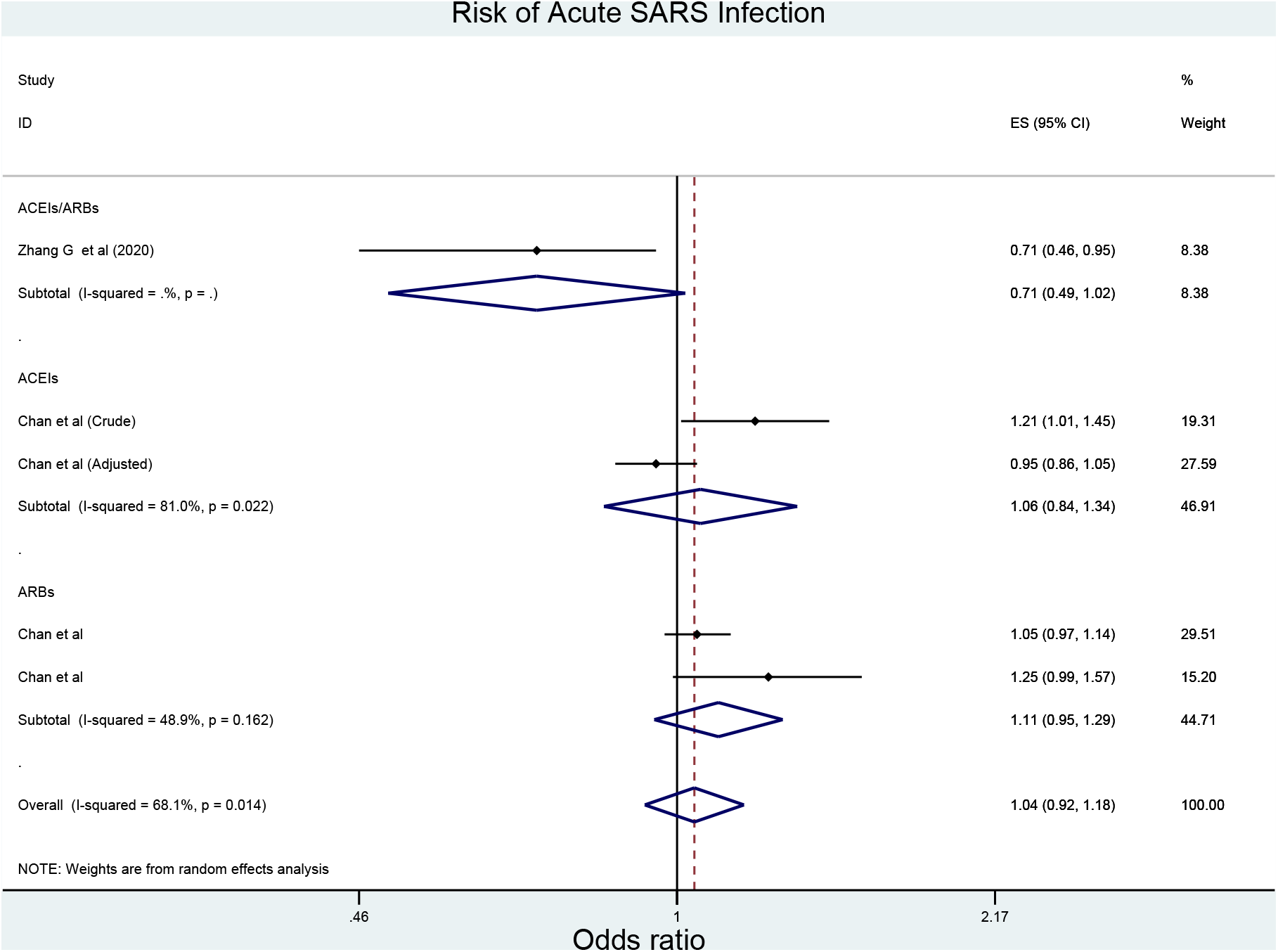
Forest plot depicting pooled estimates for the association between hospitalisation and the three level of renin-angiotensin system drug exposure (ACEIs/ARBs, ACEIs, ARBs)

**Figure 6.**
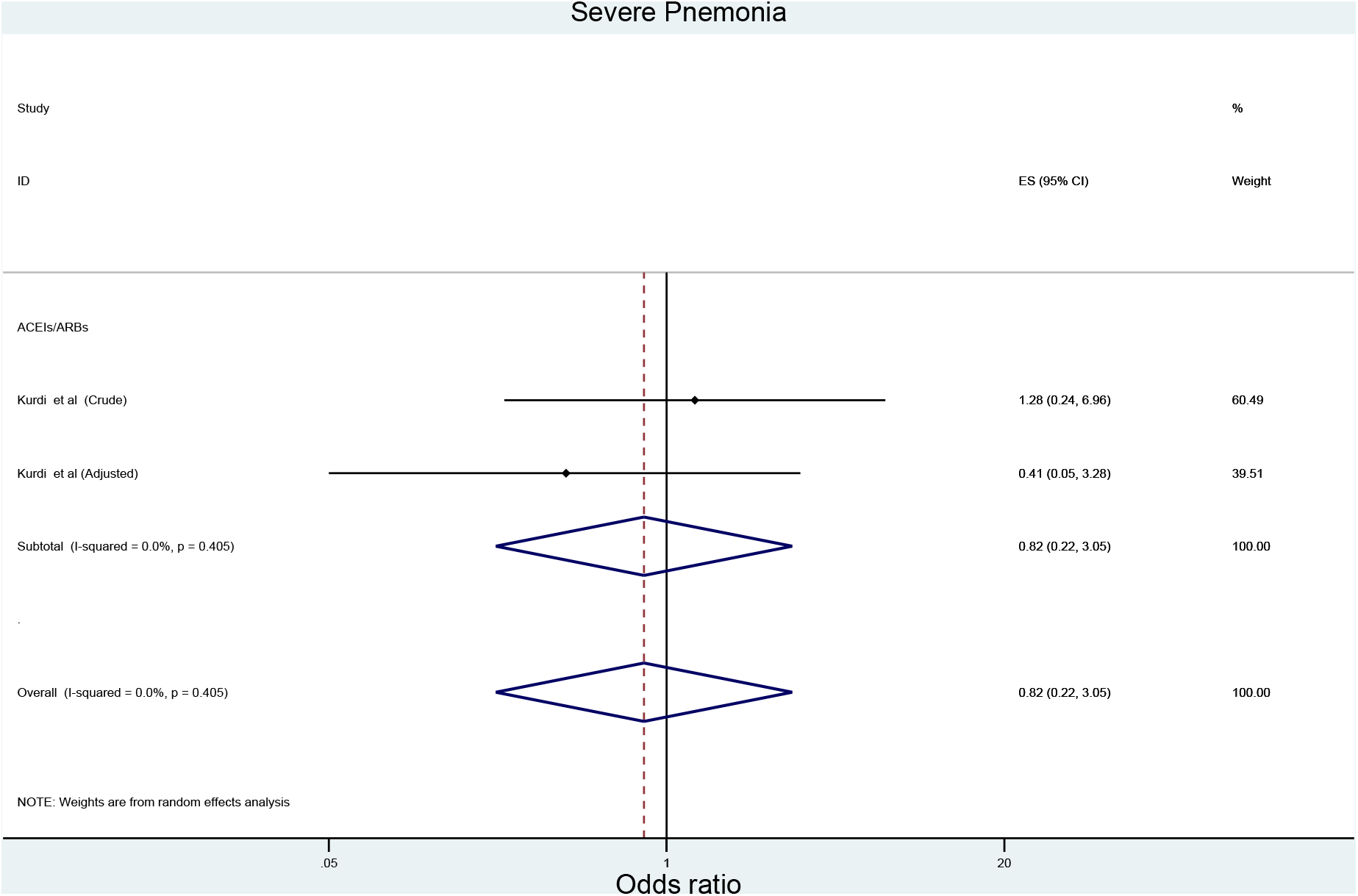
Forest plot depicting pooled estimates for the association between developing Intensive Care Unit admission and the three level of renin-angiotensin system drug exposure (ACEIs/ARBs, ACEIs, ARBs)

**Figure 7.**
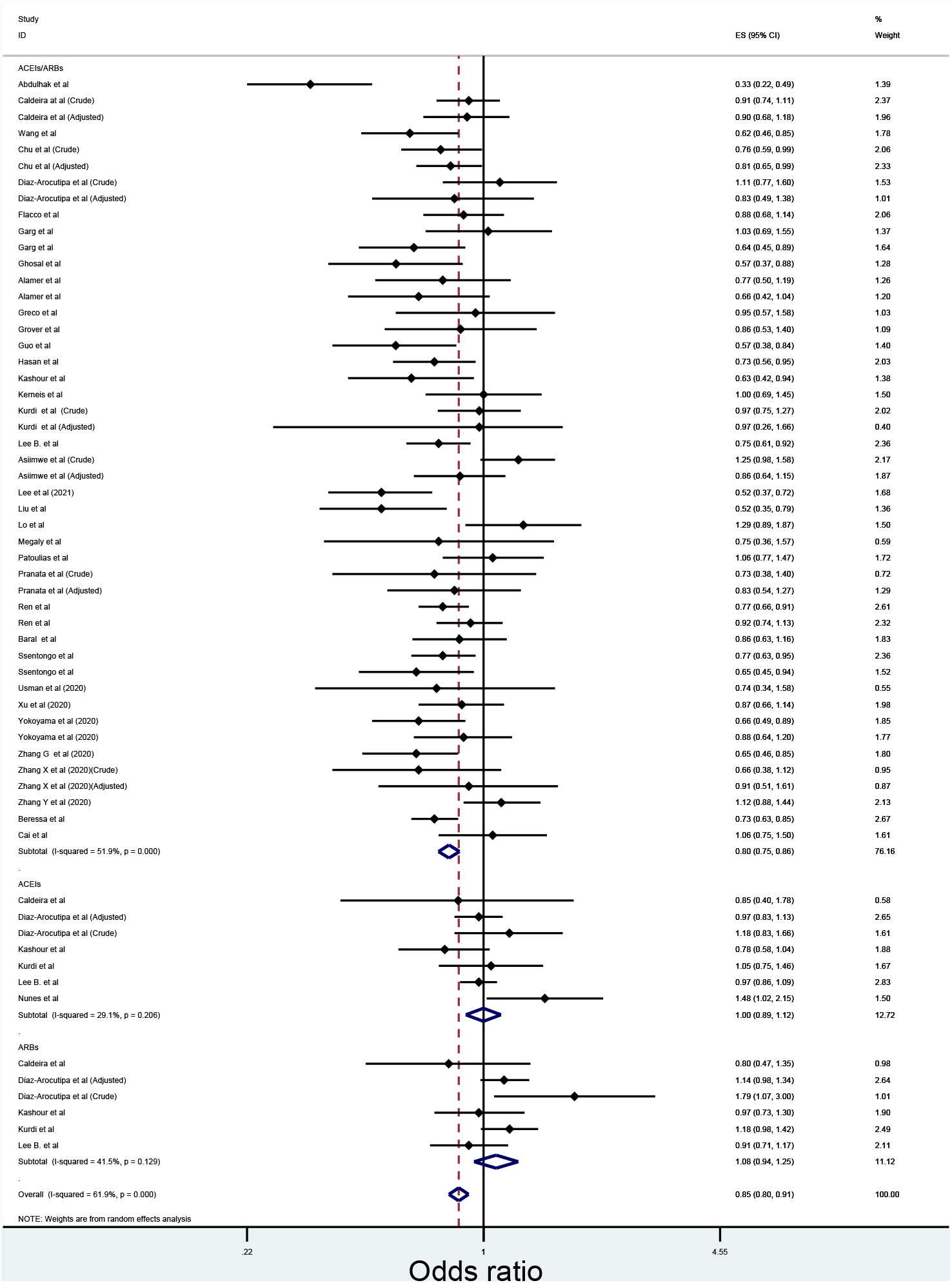
Forest plot depicting pooled estimates for the association between between risk of acquiring COVID-19 infection and the three level of renin-angiotensin system drug exposure (ACEIs/ARBs, ACEIs, ARBs)

**Figure 8.**
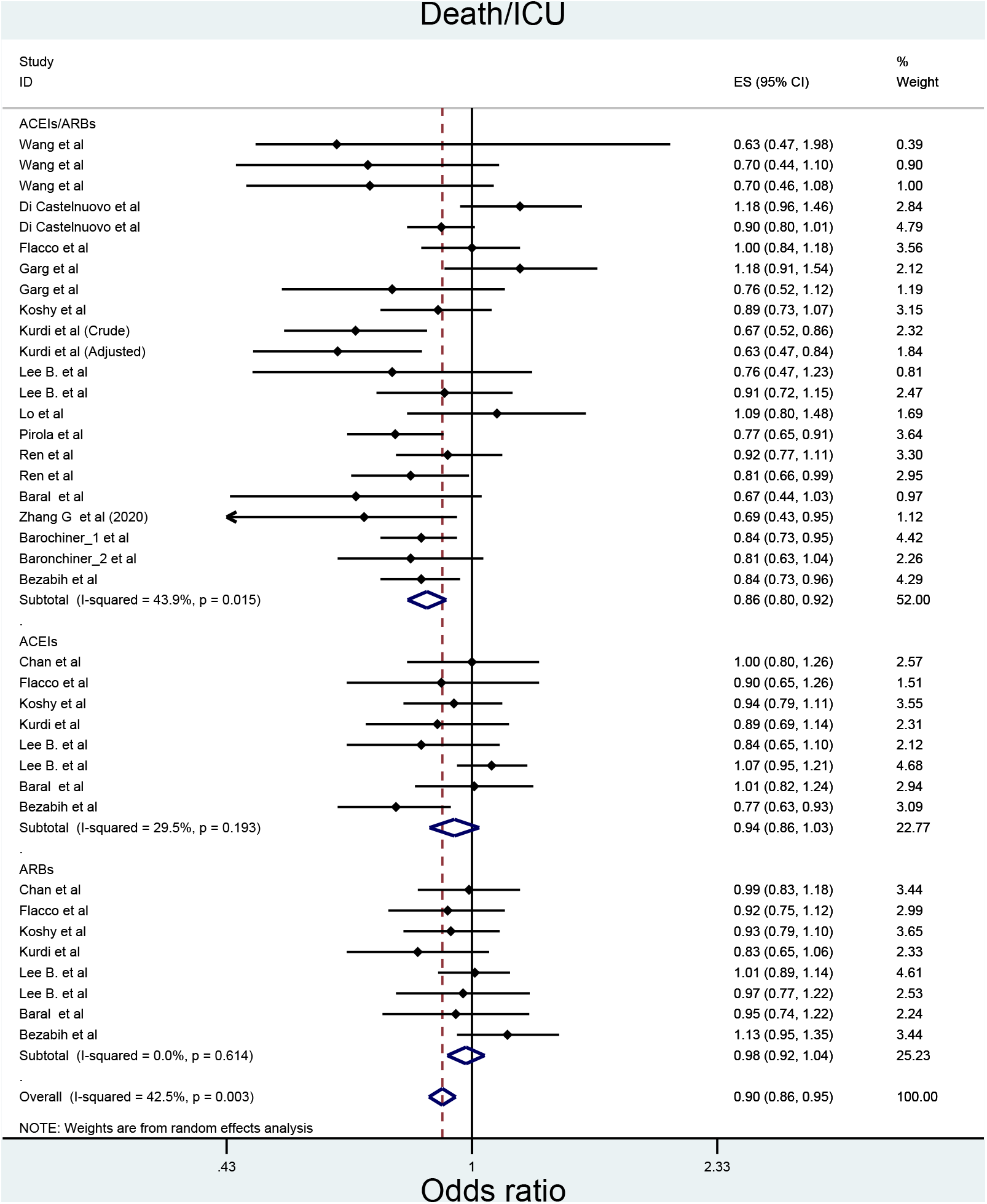
Forest plot depicting pooled estimate for the association between use of mechanical ventilator and ACEIs/ARBs use

**Figure 9.**
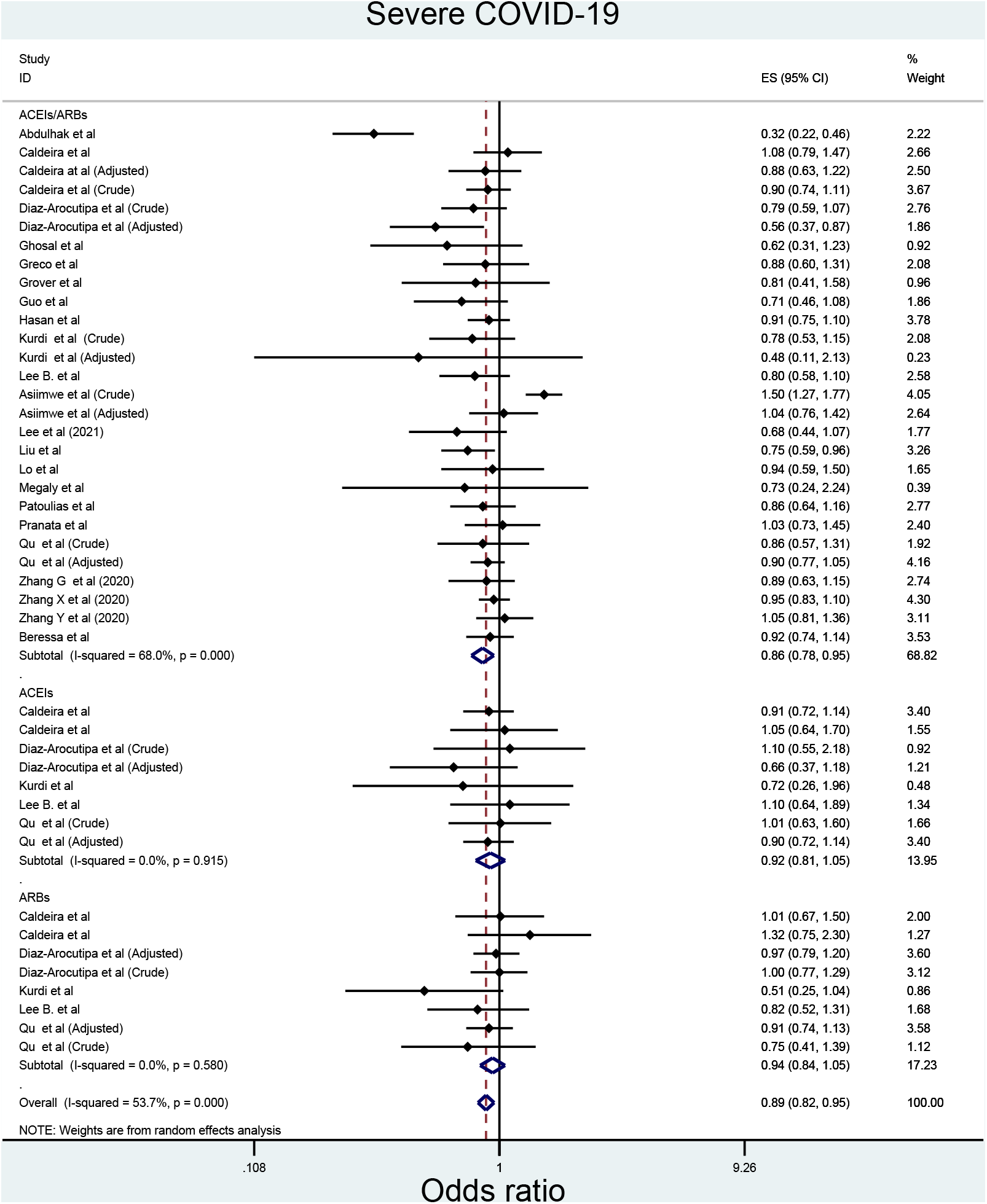
Forest plot depicting pooled estimates for the association between risk of severe acute respiratory syndrome (SARS)and the three level of renin-angiotensin system drug exposure (ACEIs/ARBs, ACEIs, ARBs)

**Figure 10.**
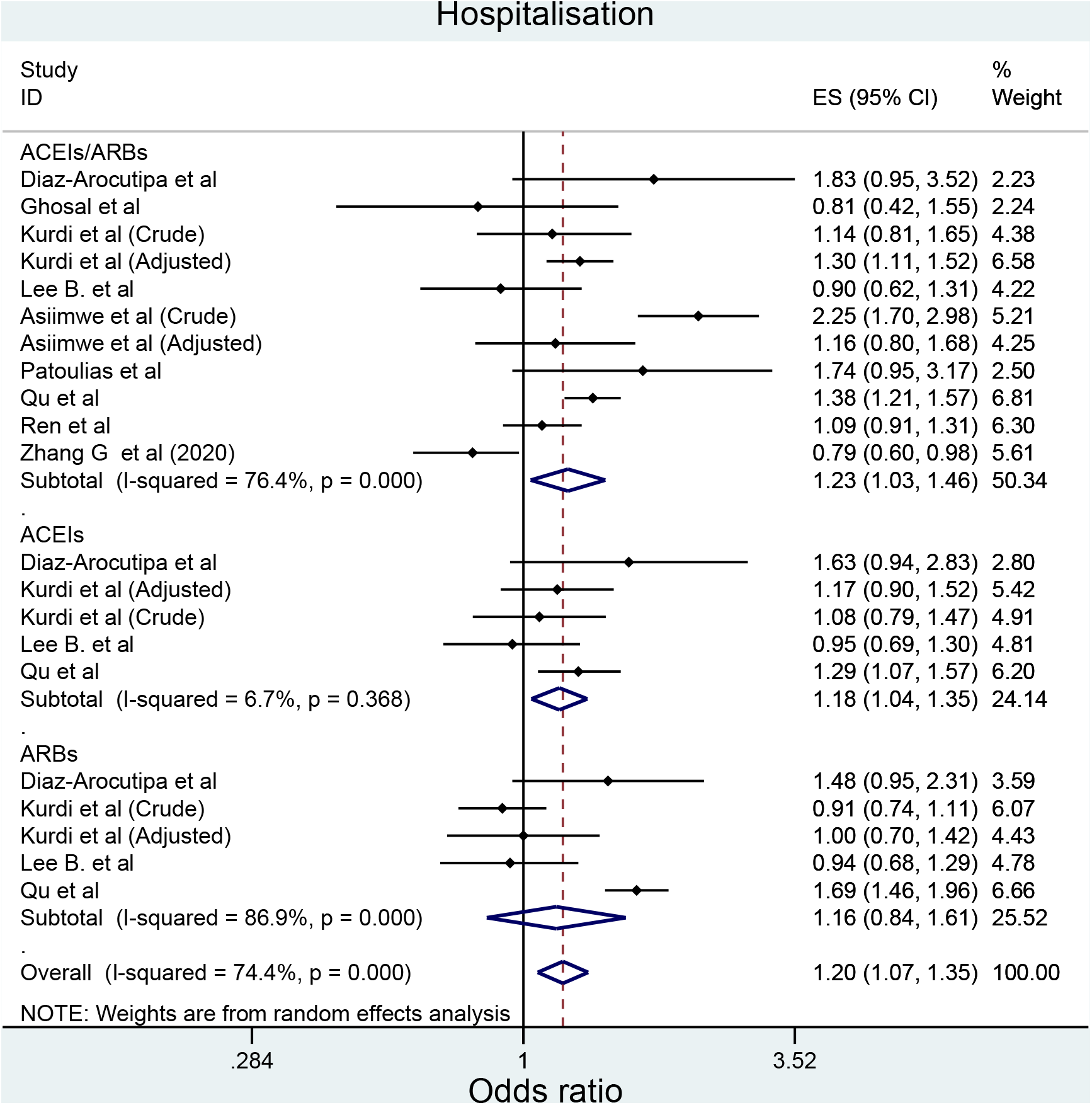
Forest plot depicting pooled estimates for the association between severe pneumonia and ACEIs/ARBs use

However, the sub-group analyses indicated different results for some of the outcomes (**Table 2**). Firstly, despite the consistent significant reduction in death in association with ACEIs/ARBs use regardless of studies’ crude/adjusted measure of effects, peer-review status and hypertension use status, there was a trend toward lower protective effective of ACEIs/ARBs on death as the quality of the studies enhanced from critically low (OR=0.75, 95%CI=0.66-0.85; I^2^= 60.4%) to moderate (OR=0.85, 95%CI=0.75-0.96; I^2^= 53.4%) (Supplementary file **6A**; **Table 2**). Similarly, the significant reduction in death/ICU admission associated with ACEIs/ARBs appeared to be higher among the studies which presented adjusted measure of effects (adjusted: OR=0.63, 95%CI=0.47-0.84 vs. crude: OR=0.87, 95%CI=0.81-0.93); and the pooled estimates for association ranged from insignificant association among the critically low-quality studies (OR=0.94, 95%CI=0.84-1.06; I^2^ = 57.4%) to a significantly higher reduction among the moderate quality studies (OR=0.74, 95%CI=0.63-0.85; I^2^ = 18.9%); (Supplementary file **7A**; **Table 2)**; besides, the significant protective impact of ACEIs/ARBs on death/ICU admission was observed only among peer-reviewed studies (peer-reviewed: OR=0.85, 95%CI=0.79-0.92 vs. non-peer reviewed: OR=0.89, 95%CI=0.75-1.10) and studies included hypertension patients (OR=0.85, 95%CI=0.80-0.90) Supplementary file **7A**; **Table 2**). Likewise, the protective effect of ACEIs/ARBs use on severe COVID-19 infection was observed only among: peer-reviewed studies (peer-reviewed: OR=0.89, 95%CI=0.83-0.96 vs. non-peer reviewed: OR=0.82, 95%CI=0.66-1.01), studies that did not recorded the hypertension status of their patients (OR=0.85, 95%CI=0.76-0.96) and critically low-quality studies (OR=0.69, 95%CI=0.53-0.92) and in fact the protective effect disappeared completely as the quality of the studies improved since insignificant association was observed among both low and moderate quality studies (OR=0.93, 95%CI=0.85-1.03; OR=0.89, 95%CI=0.77-1.04, respectively) (Supplementary file **8A**; **Table 2)**. In terms of ACEIs/ARBs’ increasing impact on hospitalisation, this impact was demonstrated only among the studies which: presented adjusted measure of effects (adjusted: OR=1.33, 95%CI=1.21-1.47 vs. crude: OR=1.21, 95%CI=0.91-1.61), were not peer-reviewed (OR=1.45, 95%CI=1.10-10.20 vs. peer-reviewed: OR=1.11, 95%CI=0.90-1.31) and did not record the hypertension status of their patients (OR=1.35, 95%CI=1.15-1.58) (Supplementary file **9A**; **Table 2**).

**Table 2.** Sub-group meta-analyses pooled estimates with 95%CI of the effects of ACEIs/ARBs on COVID-19 related clinical outcomes.

|  | **Death (n=60)** |  |  |
| --- | --- | --- | --- |
|  | **ACEIs/ARBs** | **ACEIs** | **ARBs** |
| **Adjusted outcome measure** |  |  |  |
| Adjusted OR | 0.80 (0.74, 0.91) | 0.90 (0.89, 1.12) | 1.1 (0.96, 1.26) |
| Crude OR | 0.80 (0.73, 0.86) | 1.10 (0.92, 1.25) | 1.1 (0.85, 1.42) |
| Number of studies | 10 vs. 37 | 2 vs. 5 | 2 vs. 4 |
| I-squared (p-value) | 0.0% (0.947) vs. 61% (<0.001) | 40.3% (0.196) vs. 26.7% (0.244) | 0.0% (0.335) vs. 60.6% (0.055) |
| **Peer reviewed article?** |  |  |  |
| Yes | 0.80 (0.76, 0.85) | 1.0 (0.83, 1.2) | 1.02 (0.87, 1.19) |
| No | 0.79 (0.66, 0.95) | 1.0 (0.87, 1.16) | 1.33 (0.88, 2.03) |
| Number of studies | 33 vs. 14 | 5 vs. 2 | 4 vs. 2 |
| I-squared (p-value) | 25.3% (0.095) vs. 75.3% (>0.001) | 45.7% (0.117) vs. 2.5% (0.331) | 27.2% (0.249) vs. 62.9% (0.101) |
| **Study's quality** |  |  |  |
| Critically low | 0.75 (0.66, 0.85) | 1.06 (0.57, 1.99) | 0.97 (0.37, 1.29) |
| Low | 0.81 (0.75, 0.88) | NA | NA |
| Moderate | 0.85 (0.75, 0.96) | 0.99 (0.90, 1.10) | 1.11 (0.94, 1.30) |
| Number of studies | 21 vs. 12 vs. 14 | 2 vs. 0 vs. 5 | 1 vs. 0 vs. 5 |
| I-squared (p-value) | 60.4% (>0.001) vs. 18.8% (0.259) vs. 53.4% (0.009) | 85.8% (0.008) vs. NA vs. 29.1% (0.206) | NA vs. NA vs. 48.4% (0.101) |
| **Hypertension use status** |  |  |  |
| Hypertensive patients | 0.74 (0.69, 0.79) | 0.97 (0.86, 1.09) | 0.91 (0.71, 1.17) |
| Not-recorded | 0.84 (0.77, 0.92) | 1.02 (0.87, 1.21) | 1.13 (0.98, 1.31) |
| Number of studies | 15 vs. 32 | 1 vs. 6 | 1 vs. 5 |
| I-squared (p-value) | 0.0% (0.617) vs. 57.3% (>0.001) | NA vs. 39.9% (0.140) | NA vs. 33.5% (0.129) |
|  | **ICU admission (n=18)** |  |  |
| **Adjusted outcome measure** |  |  |  |
| Adjusted OR | 0.86 (0.73, 1.02) | NA | NA |
| Crude OR | 1.09 (0.91, 1.32) | 0.96 (0.87, 1.06)* | 1.21 (0.93, 1.57)* |
| Number of studies | 2 vs. 8 | 0 vs. 4 | 0 vs. 4 |
| I-squared (p-value) | 0.0% (0.356) vs. 59.8% (0.015) | NA vs. 0.0% (0.882) | NA vs. 76.5% (0.005) |
| **Peer reviewed article?** |  |  |  |
| Yes | 0.93 (0.85, 1.01) | 0.95 (0.86, 1.05) | 1.20 (0.87, 1.66) |
| No | 1.45 (1.17, 1.80) | 1.16 (0.72, 1.86) | 1.26 (0.87, 1.83) |
| Number of studies | 9 vs. 1 | 3 vs. 1 | 3 vs. 1 |
| I-squared (p-value) | 0.0% (0.488) vs. NA | 0.0% (0.997) vs. NA | 83.1% (0.003) vs. NA |
| **Study's quality** |  |  |  |
| Critically low | 1.40 (0.80, 2.44) | NA | NA |
| Low | 0.90 (0.78, 1.03) | 0.95 (0.85, 1.06) | 0.93 (0.82, 1.05) |
| Moderate | 1.12 (0.92, 1.37) | 1.0 (0.77, 1.30) | 1.37 (1.15, 1.64) |
| Number of studies | 1 vs. 4 vs. 5 | 0 vs. 1 vs. 3 | 0 vs. 1 vs. 3 |
| I-squared (p-value) | NA vs. 22.6% (0.275) vs. 45% (0.122) | NA vs. NA vs. 0.0% (0.770) | NA vs. NA vs. 0.0% (0.742) |
| **Hypertension use status** |  |  |  |
| Hypertensive patients | 0.97 (0.75, 1.27) | 0.93 (0.52, 1.66) | 1.32 (0.97, 1.79) |
| Not-recorded | 1.05, 0.87, 1.27) | 0.96 (0.87, 1.06) | 1.18 (0.85, 1.64) |
| Number of studies | 3 vs. 7 | 1 vs. 3 | 1 vs. 3 |
| I-squared (p-value) | 0.0% (0.697) vs. 71.5% (0.002) | NA vs. 0.0% (0.722) | NA vs. 80.8% (0.006) |
| **Death/ICU admission (n=38)** |  |  |  |
| **Adjusted outcome measure** |  |  |  |
| Adjusted OR | 0.63 (0.47, 0.84) | 1.0 (0.80, 1.26) | 1.0 (0.83, 1.18) |
| Crude OR | 0.87 (0.81, 0.93) | 0.93 (0.85, 1.03) | 0.98 (0.91, 1.05) |
| Number of studies | 1 vs. 21 | 1 vs. 7 | 1 vs. 7 |
| I-squared (p-value) | NA vs. 38.9% (0.036) | NA vs. 38.5% (0.135) | NA vs. 0.0% (0.498) |
| **Peer reviewed article?** |  |  |  |
| Yes | 0.85 (0.79, 0.92) | 0.99 (0.92, 1.10) | 0.96 (0.89, 1.03) |
| No | 0.89 (0.75, 1.10) | 0.77 (0.63, 0.94) | 1.13 (0.95, 1.34) |
| Number of studies | 18 vs. 4 | 7 vs. 1 | 7 vs. 1 |
| I-squared (p-value) | 45.5% (0.019) vs. 51.5% (0.103) | 0.0% (0.605) vs. NA | 0.0% (0.874) vs. NA |
| **Study's quality** |  |  |  |
| Critically low | 0.94 (0.84, 1.06) | 0.86 (0.70, 1.04) | 1.02 (0.85, 1.24) |
| Low | 0.85 (0.79, 0.92) | 0.98 (0.82, 1.16) | 0.93 (0.80, 1.10) |
| Moderate | 0.74 (0.63, 0.85) | 0.99 90.88, 1.10) | 0.98 (0.89, 1.06) |
| Number of studies | 6 vs. 11. vs. 5 | 2 vs. 2 vs. 4 | 2 vs. 2 vs. 4 |
| I-squared (p-value) | 57.4% (0.038) vs. 15.8% (0.293) vs. 18.9% (0.294) | 56.3% (0.130) vs. 0.0% (0.568) vs. 20.7% (0.286) | 60% (0.114) vs. 0.0% (0.865) vs. 0.0% (0.572) |
| **Hypertension use status** |  |  |  |
| Hypertensive patients | 0.85 (0.80, 0.9) | 0.9 (0.75, 1.08) | 1.01 (0.93, 1.10) |
| Not-recorded | 0.88 (0.76, 1.03) | 0.96 (0.87, 1.06) | 0.93 (0.85, 1.03) |
| Number of studies | 13 vs. 9 | 4 vs. 4 | 4 vs. 4 |
| I-squared (p-value) | 0.0% (0.595) vs. 69% (0.001) | 67.1% (0.028) vs. 0.0% (0.852) | 0.0% (0.473) vs. 0.0% (0.723) |
| **Risk of COVID-19 infection (n=40)** |  |  |  |
| **Adjusted outcome measure** |  |  |  |
| Adjusted OR | 0.98 (0.94, 1.03) | 1.0 (0.82, 1.2) | 0.98 (0.56, 1.7) |
| Crude OR | 1.0 (0.97, 1.02) | 0.97 (0.93, 1.01) | 1.0 (0.97, 1.04) |
| Number of studies | 6 vs. 13 | 2 vs. 9 | 2 vs. 8 |
| I-squared (p-value) | 41.7% (0.127) vs. 18.7% (0.255) | 49% (0.161) vs. 36.6% (0.125) | 78.9% (0.03) vs. 0.0% (0.993) |
| **Peer reviewed article?** |  |  |  |
| Yes | 0.99 (0.97, 1.01) | 0.96 (0.92, 1.01) | 1.01 (0.98, 1.05) |
| No | 1.03 (0.96, 1.10) | 0.97 (0.89, 1.10) | 0.97 (0.85, 1.11) |
| Number of studies | 14 vs. 5 | 8 vs. 3 | 7 vs. 3 |
| I-squared (p-value) | 14.6% (0.294) vs. 52.5% (0.077) | 34.8% (0.150) vs. 48.6% (0.143) | 0.0% (0.814) vs. 18.1% (0.295) |
| **Study's quality** |  |  |  |
| Critically low | 0.97 (0.95, 1.0) | 0.96 (0.93, 0.99) | 1.0 (0.96, 1.04) |
| Low | 0.97 (0.93, 1.01) | 0.95 (0.84, 1.09) | 0.90 (0.62, 1.30) |
| Moderate | 1.03 (0.99, 1.06) | 1.03 (0.93, 1.14) | 1.03 (0.96, 1.10) |
| Number of studies | 4 vs. 7 vs. 8 | 4 vs. 3 vs. 4 | 4 vs. 2 vs. 4 |
| I-squared (p-value) | 0.0% (0.780) vs. 17.5% (0.296) vs. 12.7% (0.331) | 0.0% (0.811) vs. 66.7% (0.050) vs. 45.3% (0.140) | 0.0% (0.970) vs. 51.6% (0.151) vs. 0.0% (0.467) |
| **Hypertension use status** |  |  |  |
| Hypertensive patients | 1.02 (0.93, 1.11) | 1.0 (0.91, 1.11) | 1.0 (0.94, 1.08) |
| Not-recorded | 0.99 (0.97, 1.01) | 0.96 (0.92, 0.99) | 1.0 (0.97, 1.05) |
| Number of studies | 2 vs. 17 | 2 vs. 9 | 2 vs. 8 |
| I-squared (p-value) | 58.3% (0.122) vs. 19.7% (0.224) | 42.0% (0.189) vs. 33.5% (0.150) | 0.0% (0.590) vs. 0.0% (0.595) |
| **Severe COVID-19 (n=44)** |  |  |  |
| **Adjusted outcome measure** |  |  |  |
| Adjusted OR | 0.88 (0.78, 0.99) | 0.86 (0.70, 1.07) | 0.94 (0.81, 1.10) |
| Crude OR | 0.86 (0.75, 0.97) | 0.96 (0.81, 1.14) | 0.93 (0.78, 1.13) |
| Number of studies | 6 vs. 22 | 2 vs. 6 | 2 vs. 6 |
| I-squared (p-value) | 19.3% (0.287) vs. 73% (>0.001) | 0.0% (0.330) vs. 0.0% (0.954) | 0.0% (0.674) vs. 8.8% (0.360) |
| **Peer reviewed article?** |  |  |  |
| Yes | 0.89 (0.83, 0.96) | 0.94 (0.78, 1.14) | 0.91 (0.66, 1.25) |
| No | 0.82 (0.66, 1.01) | 0.9 (0.75, 1.10) | 0.95 (0.83, 1.10) |
| Number of studies | 15 vs. 13 | 4 vs. 4 | 4 vs. 4 |
| I-squared (p-value) | 0.0% (0.885) vs. 84% (>0.001) | 0.0% (0.832) vs. 0.0% (0.646) | 36.3% (0.194) vs. 0.0% (0.821) |
| **Study's quality** |  |  |  |
| Critically low | 0.69 (0.53, 0.92) | NA | NA |
| Low | 0.93 (0.85, 1.03) | 0.92 (0.75, 1.31) | 0.89 (0.73, 1.09) |
| Moderate | 0.89 (0.77, 1.04) | 0.92 (0.78, 1.10) | 0.96 (0.84, 1.10) |
| Number of studies | 7 vs. 7 vs. 14 | 0 vs. 2 vs. 6 | 0 vs. 2 vs. 6 |
| I-squared (p-value) | 80.5% (>0.001) vs. 0.0% (0.954) vs. 69.8% (>0.001) | NA vs. 0.0% (0.664) vs. 0.0% (0.782) | NA vs. 0.0% (0.557) vs. 0.0% (0.426) |
| **Hypertension use status** |  |  |  |
| Hypertensive patients | 0.89 (0.77, 1.01) | 1.10 (0.64, 1.89) | 0.82 (0.52, 1.30) |
| Not-recorded | 0.85 (0.758, 0.96) | 0.91 (0.79, 1.10) | 0.95 (0.84, 1.10) |
| Number of studies | 5 vs. 23 | 1 vs. 7 | 1 vs. 7 |
| I-squared (p-value) | 0.0% (0.684) vs. 73.1% (>0.001) | NA vs. 0.0% (0.899) | Na vs. 0.0% (0.506) |
| **Hospitalisation (n=21)** |  |  |  |
| **Adjusted outcome measure** |  |  |  |
| Adjusted OR | 1.33 (1.21, 1.47) | 1.25 (1.10, 1.46) | 1.33 (0.80, 2.23) |
| Crude OR | 1.21 (0.91, 1.61) | 1.10 (0.86, 1.41) | 1.02 (0.79, 1.31) |
| Number of studies | 3 vs. 8 | 2 vs. 3 | 2 vs. 3 |
| I-squared (p-value) | 0.0% (0.634) vs. 81.5% (>0.001) | 0.0% (0.556) vs. 27.9% (0.250) | 86.1% (0.007) vs. 49% (0.141) |
| **Peer reviewed article?** |  |  |  |
| Yes | 1.11 (0.90, 1.31) | 1.11 (0.91, 1.27) | 0.93 (0.80, 1.10) |
| No | 1.45 (1.10, 2.0) | 1.32 (1.10, 1.59) | 1.67 (1.45, 1.92) |
| Number of studies | 6 vs. 5 | 3 vs. 2 | 3 vs. 2 |
| I-squared (p-value) | 66.2% (0.011) vs. 73.1% (0.005) | 0.0% (0.611) vs. 0.0% (0.432) | 0.0% (894) vs. 0.0% (0.578) |
| **Study's quality** |  |  |  |
| Critically low | 1.20 (0.57, 2.54) | NA | NA |
| Low | 1.24 (0.98, 1.56) | 1.29 (1.07, 1.56) | 1.69 (1.46, 1.96) |
| Moderate | 1.24 (0.94, 1.63) | 1.12 (0.95, 1.31) | 0.99 (0.94, 1.19) |
| Number of studies | 2 vs. 2 vs. 7 | 0 vs. 1 vs. 4 | 0 vs. 1 vs. 4 |
| I-squared (p-value) | 64.8% (0.092) vs. 76.5% (0.039) vs. 82.9% (>0.001) | NA vs. NA vs. 0.0% (0.368) | NA vs. NA vs. 23.9% (0.268) |
| **Hypertension use status** |  |  |  |
| Hypertensive patients | 0.82 (0.67, 1.01) | 0.95 (0.69, 1.30) | 0.94 (0.68, 1.31) |
| Not-recorded | 1.35 (1.15, 1.58) | 1.23 (1.10, 1.41) | 1.23 (0.84, 1.78) |
| Number of studies | 2 vs. 9 | 1 vs. 4 | 1 vs. 4 |
| I-squared (p-value) | 0.0% (0.568) vs. 66% (0.003) | NA vs. 0.0% (0.553) | NA vs. 88.7% (>0.001) |
| **Ventilator use (n=5)** |  |  |  |
| **Adjusted outcome measure** |  |  |  |
| Adjusted OR | NA | NA | NA |
| Crude OR | 1.18 (0.84, 1.66)* | 1.01 (0.03, 34.52)* | 0.985 (0.084, 11.57)* |
| Number of studies | 0 vs. 3 | 0 vs. 1 | 0 vs. 1 |
| I-squared (p-value) | NA vs. 53.4% (0.114) | NA | NA |
| **Peer reviewed article?** |  |  |  |
| Yes | 1.10 (0.66, 1.75) | 1.01 (0.03, 34.52)* | 0.985 (0.084, 11.57)* |
| No | 1.39 (0.99, 1.95) | NA | NA |
| Number of studies | 2 vs. 1 | 1 vs. 0 | 1 vs. 0 |
| I-squared (p-value) | 52.6% (0.146) vs. NA | NA | NA |
| **Study's quality** |  |  |  |
| Critically low | NA | NA | NA |
| Low | NA | NA | NA |
| Moderate | 1.18 (0.84, 1.66)* | 1.01 (0.03, 34.52)* | 0.985 (0.084, 11.57)* |
| Number of studies | 0 vs. 0 vs. 3 | 0 vs. 0 vs. 1 | 0 vs. 0 vs. 1 |
| I-squared (p-value) | NA vs. NA vs. 53.4% (0.114) | NA | NA |
| **Hypertension use status** |  |  |  |
| Hypertensive patients | 0.89 (0.65, 1.23) | NA | NA |
| Not-recorded | 1.41 (1.10, 1.90) | 1.014 (0.030, 34.758)* | 0.985 (0.084, 11.570)* |
| Number of studies | 1 vs. 2 | 0 vs. 1 | 0 vs. 1 |
| I-squared (p-value) | NA vs. 0.0% (0.844) | NA | NA |
| **Acute SARS (n=5)** |  |  |  |
| **Adjusted outcome measure** |  |  |  |
| Adjusted OR | NA | 0.95 (0.86, 1.05) | 1.05 (0.97, 1.14) |
| Crude OR | 0.71 (0.49, 1.02) | 1.21 (1.01, 1.45) | 1.25 (0.99, 1.57) |
| Number of studies | 0 vs. 1 | 1 vs. 1 | 1 vs. 1 |
| I-squared (p-value) | NA | NA | NA |
| **Peer reviewed article?** |  |  |  |
| Yes | 0.71 (0.49, 1.02)* | 1.06 (0.84, 1.34)* | 1.11 (0.95, 1.29)* |
| No | NA | NA | NA |
| Number of studies | 1 vs. 0 | 2 vs. 0 | 2 vs. 0 |
| I-squared (p-value) | NA | 81% (0.022) vs. NA | 48.9% (0.162) vs. NA |
| **Study's quality** |  |  |  |
| Critically low |  |  |  |
| Low | NA | NA | NA |
| Moderate | NA | NA | NA |
| Number of studies | 0.71 (0.49, 1.02) | 1.06 (0.84, 1.34)* | 1.11 (0.95, 1.29)* |
| I-squared (p-value) | 0 vs. 0 vs. 1 | 0 vs. 0 vs. 2 | 0 vs. 0 vs. 2 |
| Hypertension use status |  | NA vs. NA. vs. 81% (0.022) | NA vs. NA. vs. 48.9% (0.162) |
| **Hypertensive patients** | 0.71 (0.49, 1.02) | NA | NA |
| Not-recorded | NA | 1.06 (0.84, 1.34) | 1.11 (0.95, 1.29) |
| Number of studies | 1 vs. 0 | 0 vs. 2 | 0 vs. 2 |
| I-squared (p-value) | NA | NA vs. 81% (0.022) | NA vs. 48.9% (0.162) |
| (Note) *Indicates that the pooled estimate is the same as the overall analyses because all the studies were in one group; NA: not applicable indicating that no studies were available to perform meta-analyses for these outcomes; |  |  |  |

### Effect of ACEIs and AEBs (as a separate group) on the study outcomes

Overall, the effect of ACEIs and ARBs on seven COVID-19 related clinical outcomes (death, ICU admission, death/ICU admission, risk of acquiring COVID-19 infection, severe COVID-19 infection, hospitalisation, and acute SARS) were evaluated. Neither ACEIs nor ARBs had any significant impact on any of the seven studied outcomes (**Figures 2-10; Table 1)** except for hospitalisation whereby ACEIs use was associated with a significant increase in COVID-19 related hospitalisation (OR=1.18, 95%CI=1.04-1.35; I^2^ = 6.7%) (Figure **5**; **Table 1**). These results were mostly consistent across all the sub-group analyses (**Supplementary Files 6B&C, 7B&C, 8B&C; Table 2)** except for the increasing effect of ACEIs on hospitalisation which was only observed among those studies which did not record the hypertension status of their patients (OR=1.23, 95%CI=1.10-1.41) (**Supplementary Files 9B&C; Table 2)**

### Publication bias

Results from the funnel plots (Supplementary file **10**) and Egger’s asymmetry tests for the six outcomes (death, ICU admission, death/ICU admission, risk of acquiring COVID-19 infection, severe COVID-19 infection, and hospitalisation) that were reported by at least 10 studies indicated no evidence of significant publication bias in all of them except for death/ICU admission and severe COVID-19 infection (p-value=0.022 and 0.019, respectively).

### Influential analyses

The results from the influential analyses indicated that none of the combined pooled meta-analysis estimates for the nine outcomes were dominated/influenced by an individual study since the omission of any of these individual studies one at a time made no difference to the pooled meta-analysis estimate because all of pooled meta-analysis estimates were overlapping (Supplementary file **11**).

## Discussion

This umbrella review for the first time combined all the available evidence so far from observational studies on the impact of ACEIs/ARBs on COVID-19 clinical outcomes (47 systematic review studies which reported 213 meta-analyses) into one pooled estimate using an umbrella review and meta-analysis approach. The collective, combined pooled estimates indicated evidence of statistically significant reduction in mortality, death/ICU admission (as a composite endpoint) and severe COVID-19 infection in association with ACEIs/ARBs use, but significant increase in the risk of hospitalisation (Table 1). Interestingly, when analysing ACEIs and ARBs as a two separate groups, there was no evidence of any significant association between ACEIs, or ARBs and any of the nine COVID-19 related clinical outcomes analysed in our study.

Although the magnitude of observed impact of ACEIs/ARBs use on reducing mortality was decreasing as the quality of studies improved (ranged from 25% reduction death- OR=0.75; 95%CI: 0.66, 0.85-among critically low-quality studies to 15% reduction- OR=0.85; 95%CI: 0.75, 0.96-among moderate-quality studies) (Table 2), the evidence were overall mostly consistent across all the sub-group analyses including a greater impact among studies that included hypertensive patients (26% reduction- OR=0.74; 95%CI: 0.69, 0.79) compared with studies that did not record the hypertension status of their study population (14% reduction-OR=0.84; 95%CI: 0.77, 0.92). In terms of death/ICU admission, the quality of the evidence was even better because the impact of ACEIs/ARBs use was greater and significant only among: moderate-quality studies (26% reduction- OR=0.63, 0.85), peer-reviewed studies (15% reduction- OR=0.85; 95%CI: 0.79, 0.92), and studies with hypertensive patients (15% reduction; OR=0.85; 95%CI: 0.80, 0.90); however, the impact was significant regardless of whether the measure of effects was crude or adjusted, even though the impact was greater among studies with adjusted measure of effects (37% reduction- OR=0.63; 95%CI: 0.47, 0.84) compared with 13% reduction (OR=0.87; 95%CI: 0.81, 0.93) among studies with crude measure of effects. In contrast, the quality of the evidence for the impact of ACEIs/ARBs use on severe COVID-19 was low since a significant reduction was only observed among critically-low quality studies (31% reduction- OR=0.69; 95%CI: 0.53, 0.92) and in fact, the significant association disappeared as the quality of the studied enhanced from critically low quality to either low or moderate quality.

In terms of the impact of ACEIs/ARBs on hospitalisation, the quality of the evidence was low because the significant association was not apparent when the data were analysed by the quality of the studies, even though the magnitude of the effect was almost consistent across the various quality of the studies; besides, the significant increase in hospitalisation was observed only among: studies that reported adjusted measure of effects (33% increase- OR=0.1.33; 95%CI; 1.21, 1.47), non-peer reviewed studies (45% increase- OR=1.45; 95%CI: 1.10, 2.0) and studies that did not recorded the hypertensive status of their study population (35% increase- OR=1.35; 95%CI: 1.15, 1.58).

Furthermore, the sub-group analyses demonstrated some low-quality evidence regarding the impact of ACEIs and ARBs (as separate groups) whereby ARBs use was associated with a significant increase in hospitalisation only among the studies that were of low-quality (69% increase- OR=1.69; 95%CI: 1.46, 1.96) and non-peer reviewed (67% increase- OR=1.67; 95%CI: 1.45, 1.92); whereas ACEIs use increased hospitalisation significantly by 23% (OR=1.23; 95%CI: 1.10, 1.41) only among studies that did not report the hypertensive status of their study population. This observed difference between ARBs and ACEIs in their impact on COVID-19 clinical outcomes has been suggested to be due to the increased level of angiotensin-II, which occurs following ARBs treatment but not ACEIs, which in turn imposes an increased substrate load on ACE2 enzyme (the key cell entry point for COVID-19) requiring its upregulation (62); hence facilitates COVID-19 virus cell entry and its subsequent infectivity/pathogenicity (63). Furthermore, the increase in ACE2 activity demonstrated in patients with hypertension, either due to the pathophysiology of hypertension itself (64) or administration ACEIs/ARBs as antihypertensive medications (65), could at least partially explain some of our study findings as why ACEIs/ARBs had significant impact on certain COVID-19 clinical outcomes only among studies that included patient with hypertension.

Several hypotheses have been suggested to explain the potential negative and positive effects of ACEIs/ARBs use on COVID-19 clinical outcomes. The negative effects are hypothesised to be due to ACEIs/ARBs induced upregulation of ACE2 expression; hence enhancing viral binding and cell entry (65); whereas the positive protective effects could be through ACEIs/ARBs effects on angiotensin II expression leading to subsequent increase in the protective angiotensin 1-7 and 1-9 which have anti-inflammatory and vasodilatory effects; hence potentially attenuating the cardiac and pulmonary damages (2). Genetic ACE2 polymorphism among some individuals has been also suggested as potential factor explaining, at least partially, the harmful effects on ACEIs/ARBs on COVID-19 outcomes (66).

Our study findings are in contrast to the findings from a recent randomised clinical trial (RCT) (67) which found insignificant differences in the mean number of days alive and out of the hospital between those assigned to discontinue vs continue ACEIs or ARBs. However, there are certain points that should be considered when interpreting the findings from this clinical trial in comparison to our study findings. First, this RCT was designed to evaluate the impact of continuing ACEIs or ARBs vs. their discontinuation after contracting COVID-19 rather than evaluating ACEIs/ARBs use vs. non-use of these medication which was the focus of most of the observational studies involved in our current study. Secondly, the RCT included only patients with mild or moderate COVID-19 with more than half of the participants (57%; n=376) having mild COVID-19, and evaluated only two COVID-19 related clinical outcomes, namely days alive (mortality) and out of hospital days; hence leaving a big gap in the evidence around ACEIs/ARBs’ impact on other important COVID-19 clinical outcomes such is ICU admission, hospitalisation, acquiring COVID-19 infection and severe COVID-19 as well as limiting the findings’ external validity (generalisability) to patients with severe COVID-19. Furthermore, although the RCT’s participants were all hypertensive patients, about one-third (∼31%) and ∼1% had diabetes and heart failure, respectively, which further limits the generalisability of the RCT’s findings to these conditions for which ACEIs/ARBs are commonly indicated. Moreover, the RCT’s participants were all from Brazil and hence extending the findings to other races or ethnicities will be limited; this is particularly importantly because there are evidence demonstrating that there are potential genetic variants of renin, angiotensinogen, ACE, angiotensin II and ACE2 among various populations that influence the function of the renin-angiotensin aldosterone system; hence affecting someone’ response to the COVID-19 infection (68). Finally, it is not entirely clear how long it takes for the ACE2 upregulation (induced by ACEIs/ARBs treatment) to return to its normal level after discontinuing ACEIs/ARBs therapy, suggesting that measuring any clinical outcome within 30 days might not be long enough for the ACE2 level to return back to its pre-ACEIs/ARBs treatment level (i.e., ACE2 level would be comparable between those continued or discontinued ACEIs/ARBs treatment) which could potentially explain the insignificant difference in the study outcomes between the two groups in the RCT; however, this requires further investigation.

It is rather surprising and unusual to have such high number of published systematic reviews and meta-analysis (47 studies) on the same topic. Circumstances associated with the pandemic may have influenced researchers’ decisions and overall study quality. For example, researchers may have decided not to submit a published protocol to quicken the review process for rapid dissemination of results to clinicians and COVID-19 policy makers (41).

### Strengths and limitations

This review presents the most comprehensive and systematic overview on the impact using RAAS inhibitors on COVID-19 related clinical outcomes, with a wide range of sensitivity (sub-group) analyses to assess the strength, validity and robustness of the evidence while accounting for potential confounding variables. Furthermore, none of the pooled meta-analysis estimates for the nine studied outcomes was affected/dominated by a single individual study. Although most of the included studies were classified as ‘low’ or ‘critically low’ quality when assessed using AMSTAR 2 tool, it is widely acknowledged that the AMSTAR 2 tool has a high standard with most reviews rated as ‘critically low’ (69, 70). The AMSTAR 2 tool is also prone to subjective biases (71), and assessment results are at the discretion of the reviewers regarding what is a “comprehensive” literature search or “satisfactory” explanation of heterogeneity or risk of bias assessment (71); therefore, quality assessment was conducted fully independent in this review and further criteria were set by the assessors to ensure inter-rater consistency. Alternatives tools to AMSTAR 2 exist such as the ROBIS tool, however the measurement categories are found to be broadly similar with the AMSTAR 2 tool considered more reliable (71). Additionally, we accounted for this issue by conducting a sub-group analysis based on the level of studies’ quality.

## Conclusion

Collective evidence so far from observational studies indicate a good quality evidence on the significant association between ACEIs/ARBs use and reduction in death and death/ICU admission (as a composite outcome). Additionally, ACEIs/ARBs use was found to be associated with a significant reduction in severe COVID-19 but a significant increase in hospitalisation; however, the evidence for these two outcomes was of poor quality; hence, cautious interpretation of these findings is required. Interestingly, findings for some of the clinical outcomes were dependent on whether the included patients had hypertension or not. Overall, our study findings further support the current recommendations of not discontinuing ACEIs/ARBs therapy in patients with COVID-19 due to the lack of good quality evidence on their harm but rather it could be beneficial to patients.

## Supporting information

PRISMA checklist

## Data Availability

All data produced in the present study are available upon reasonable request to the authors

## Supplementary files’ captions and legends

Supplementary file 1. Search strategy used in the database searches

Supplementary file 2. List and details of the irrelevant studies excluded at the stage of abstract and title screening

Supplementary file 3. Study characterises of the 47 eligible reviews included in the current umbrella systematic review

**Supplementary file 4. Details of all the 213 meta-analyses conducted by the eligible 47 reviews**

Supplementary file 5. Quality assessment score of the 47 eligible reviews included in the current umbrella systematic review using AMSTAR 2 tool

Supplementary file 6. Forest plot depicting sub-group analyses pooled estimates for the association between mortality and ACEIs/ARBs use sub-grouped by A) type of analyses (crude vs. adjusted); B) peer-review status; C) methodological quality; and D) hypertension stats

Supplementary file 6A. Forest plot depicting sub-group analyses pooled estimates for the association between mortality and ACEIs use sub-grouped by A) type of analyses (crude vs. adjusted); B) peer-review status; C) methodological quality; and D) hypertension stats

Supplementary file 6B. Forest plot depicting sub-group analyses pooled estimates for the association between mortality and ARBs use sub-grouped by A) type of analyses (crude vs. adjusted); B) peer-review status; C) methodological quality; and D) hypertension stats

Supplementary file 7. Forest plot depicting sub-group analyses pooled estimates for the association between death/ICU admission (as a composite outcome) and ACEIs/ARBs use sub-grouped by A) type of analyses (crude vs. adjusted); B) peer-review status; C) methodological quality; and D) hypertension stats

Supplementary file 7A. Forest plot depicting sub-group analyses pooled estimates for the association between death/ICU admission (as a composite outcome) and ACEIs use sub-grouped by A) type of analyses (crude vs. adjusted); B) peer-review status; C) methodological quality; and D) hypertension stats

Supplementary file 7B. Forest plot depicting sub-group analyses pooled estimates for the association between death/ICU admission (as a composite outcome) and ARBs use sub-grouped by A) type of analyses (crude vs. adjusted); B) peer-review status; C) methodological quality; and D) hypertension stats

Supplementary file 8. Forest plot depicting sub-group analyses pooled estimates for the association between severe COVID-19 and ACEIs/ARBs use sub-grouped by A) type of analyses (crude vs. adjusted); B) peer-review status; C) methodological quality; and D) hypertension stats

Supplementary file 8A. Forest plot depicting sub-group analyses pooled estimates for the association between severe COVID-19 and ACEIs use sub-grouped by A) type of analyses (crude vs. adjusted); B) peer-review status; C) methodological quality; and D) hypertension stats

Supplementary file 8B. Forest plot depicting sub-group analyses pooled estimates for the association between severe COVID-19 and ARBs use sub-grouped by A) type of analyses (crude vs. adjusted); B) peer-review status; C) methodological quality; and D) hypertension stats

Supplementary file 9. Forest plot depicting sub-group analyses pooled estimates for the association between hospitalisation and ACEIs/ARBs use sub-grouped by A) type of analyses (crude vs. adjusted); B) peer-review status; C) methodological quality; and D) hypertension stats

Supplementary file 9A. Forest plot depicting sub-group analyses pooled estimates for the association between hospitalisation and ACEIs use sub-grouped by A) type of analyses (crude vs. adjusted); B) peer-review status; C) methodological quality; and D) hypertension stats

Supplementary file 9B. Forest plot depicting sub-group analyses pooled estimates for the association between hospitalisation and ARBs use sub-grouped by A) type of analyses (crude vs. adjusted); B) peer-review status; C) methodological quality; and D) hypertension stats

Supplementary file 10. Publication bias funnel plot for the outcomes with >=10 studies

Supplementary file 11. Results of the influential analyses

