## Supplementary figures and images for "An umbrella review and meta-analysis of the use of renin-angiotensin system drugs and COVID-19 outcomes: what do we know so far?"

### Supplementary file 6

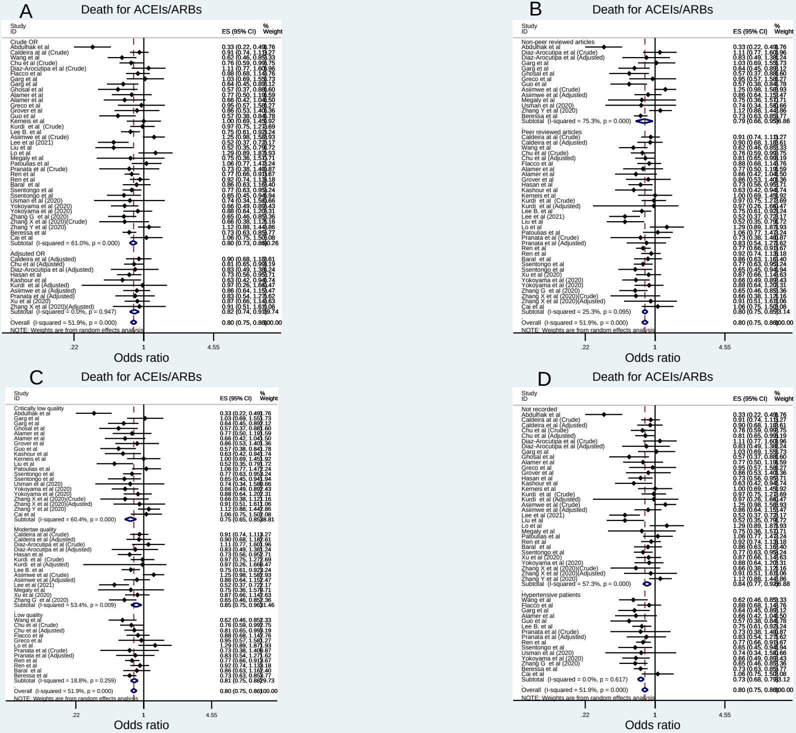

### Supplementary file 6A

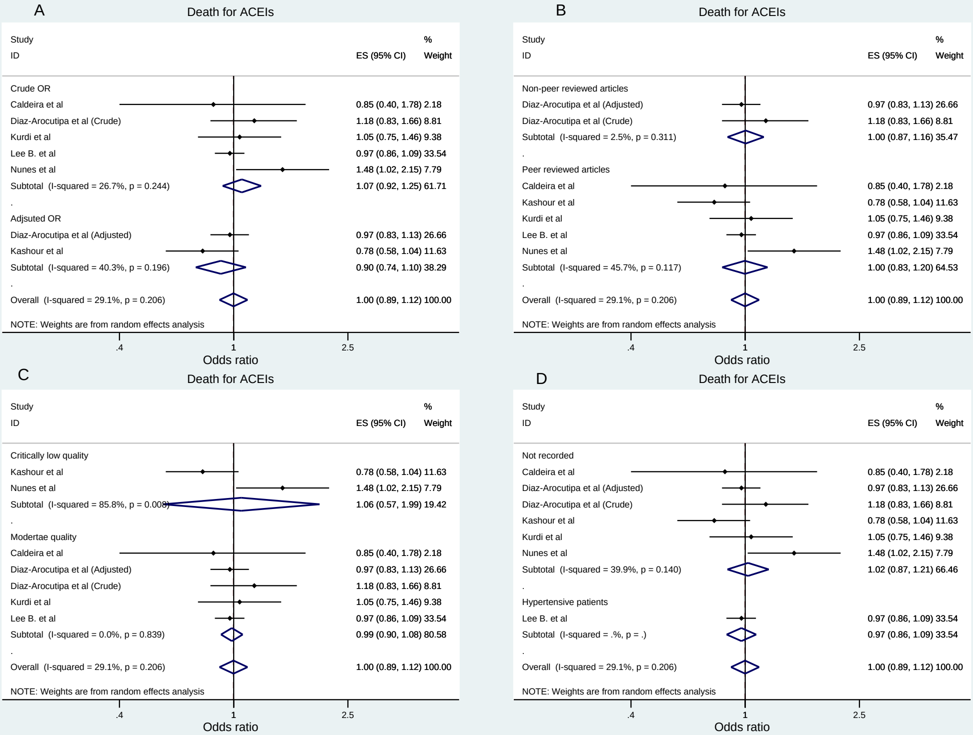

### Supplementary file 6B

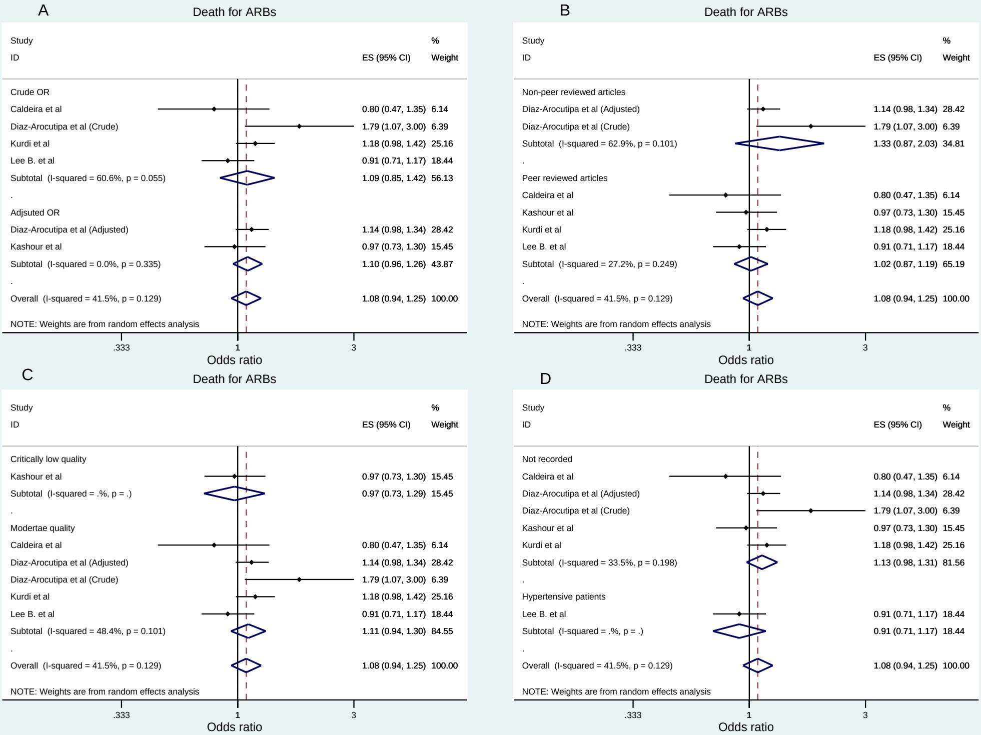

### Supplementary file 7

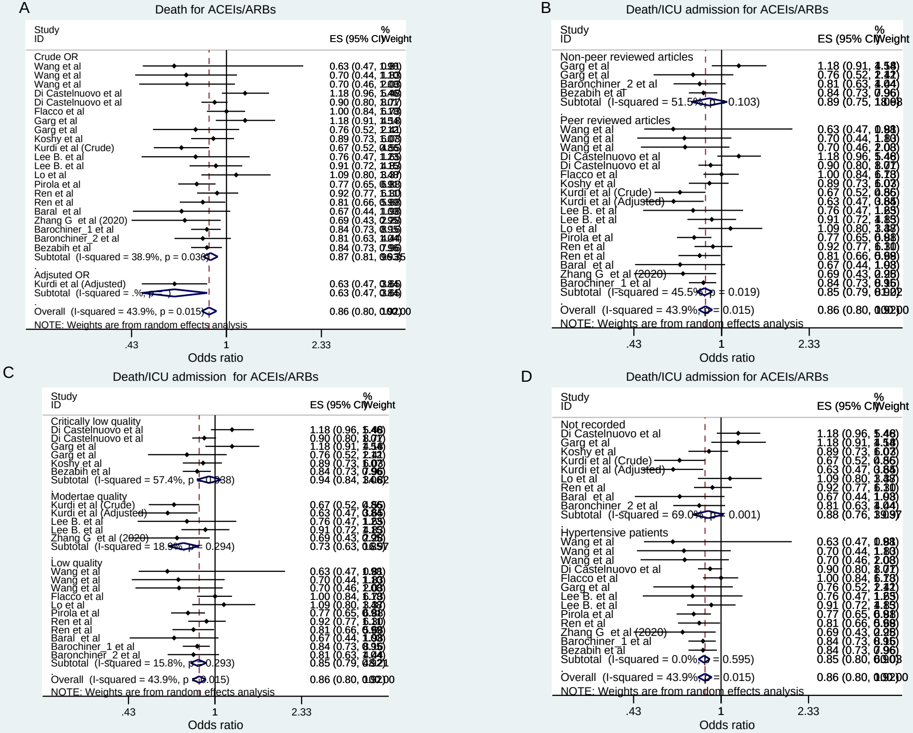

### Supplementary file 7A

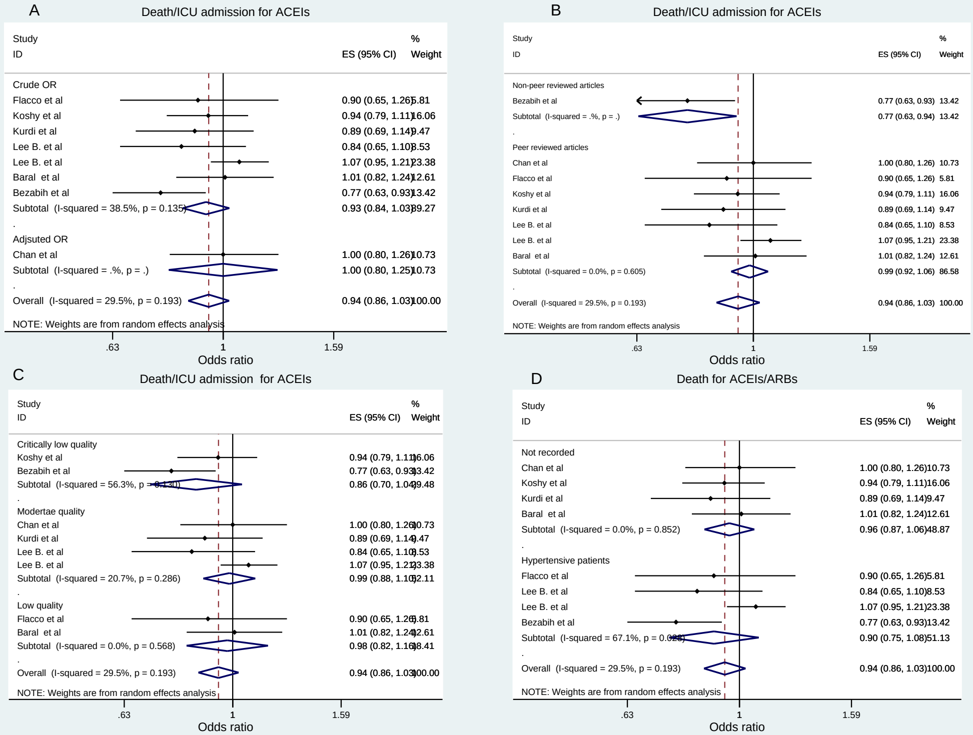

### Supplementary file 7B

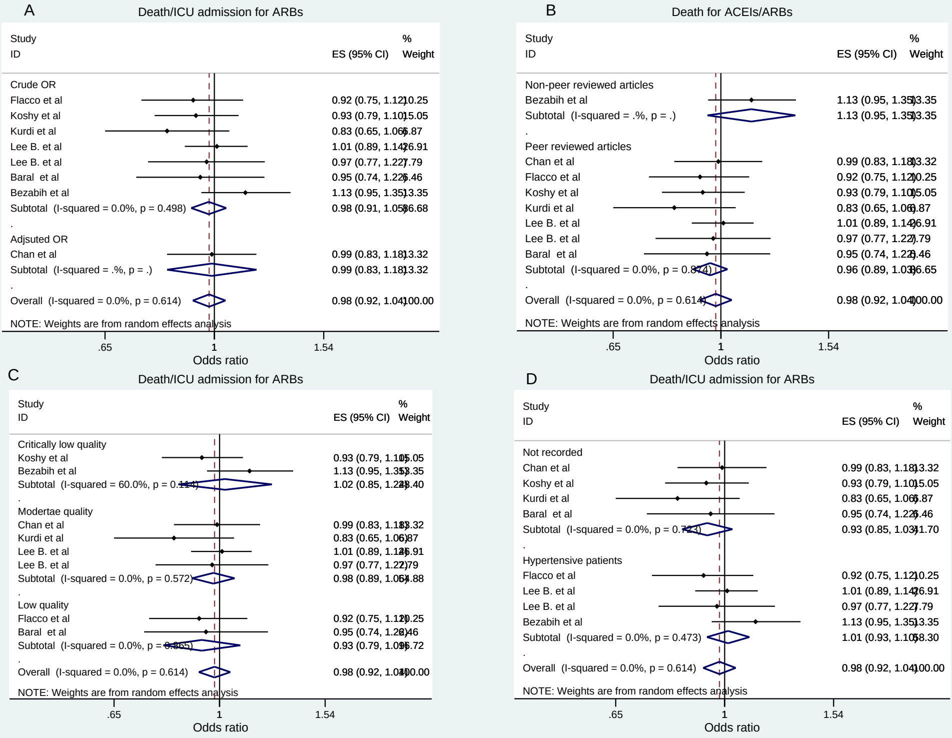

### Supplementary file 8

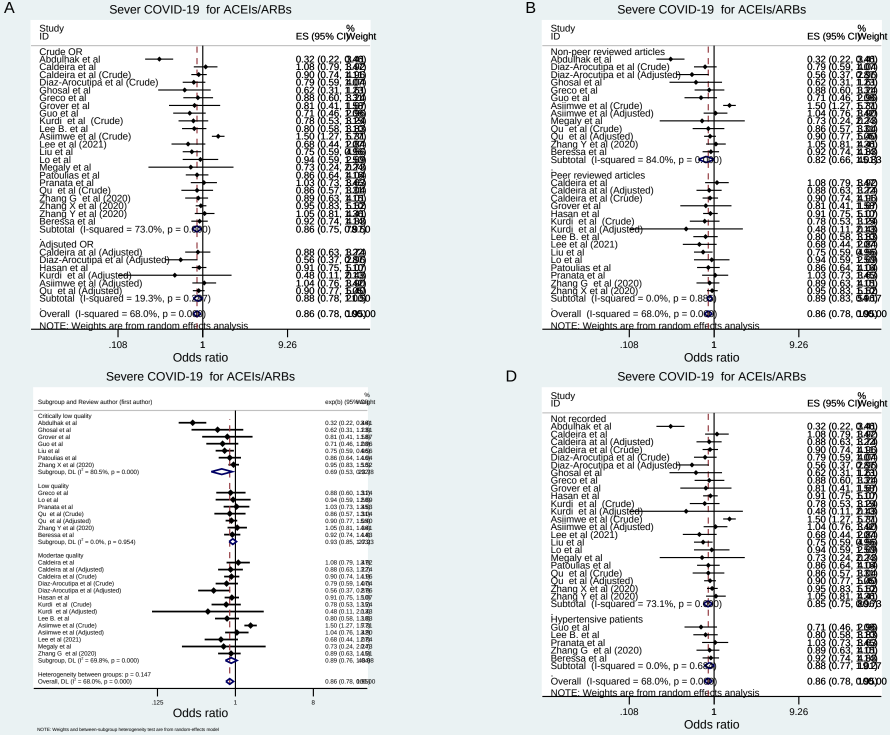

### Supplementary file 8A

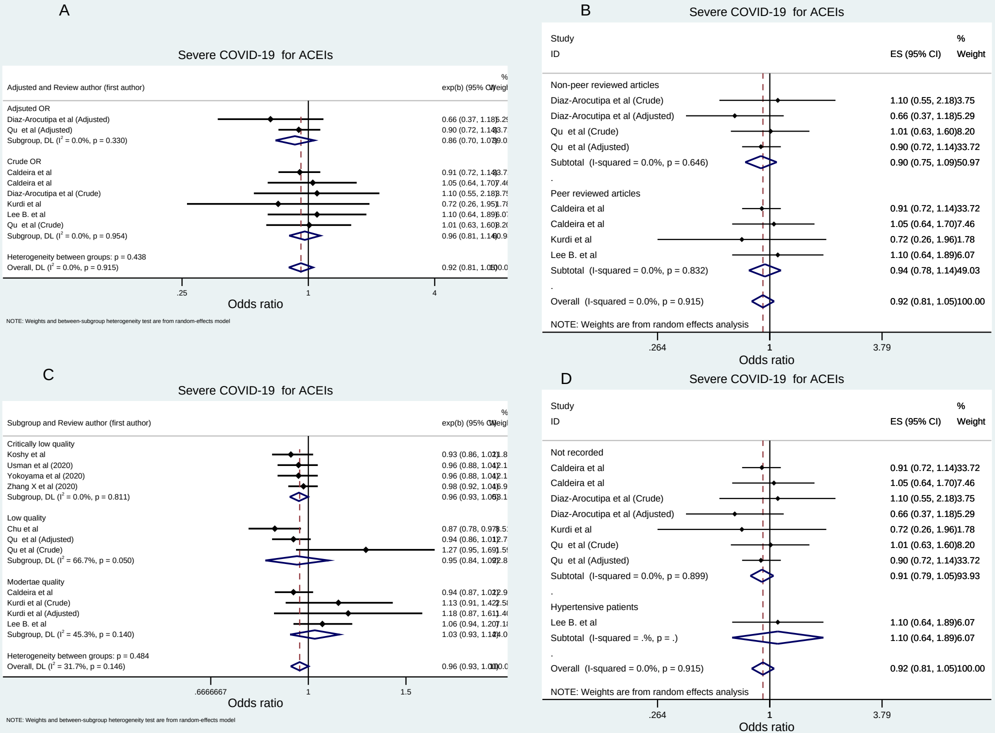

### Supplementary file 8B

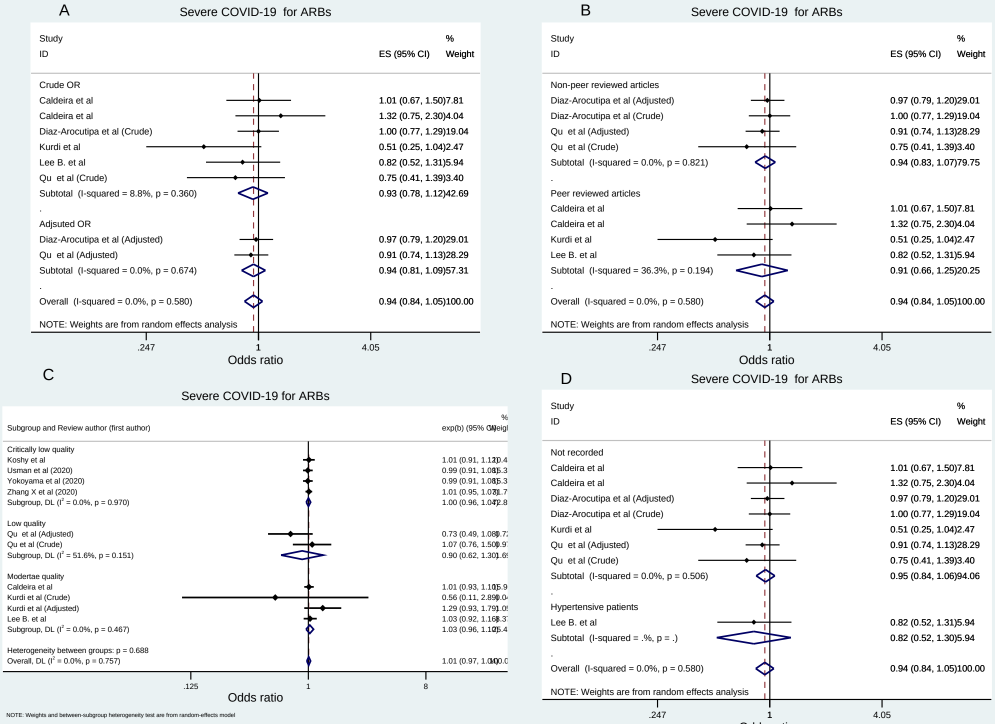

### Supplementary file 9

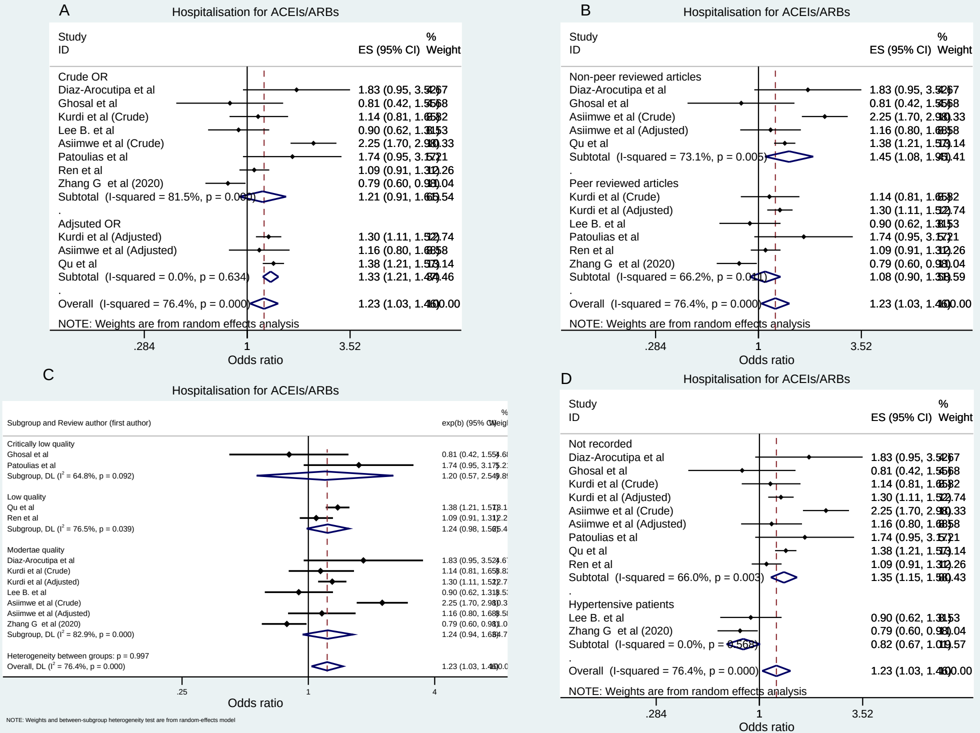

### Supplementary file 9A

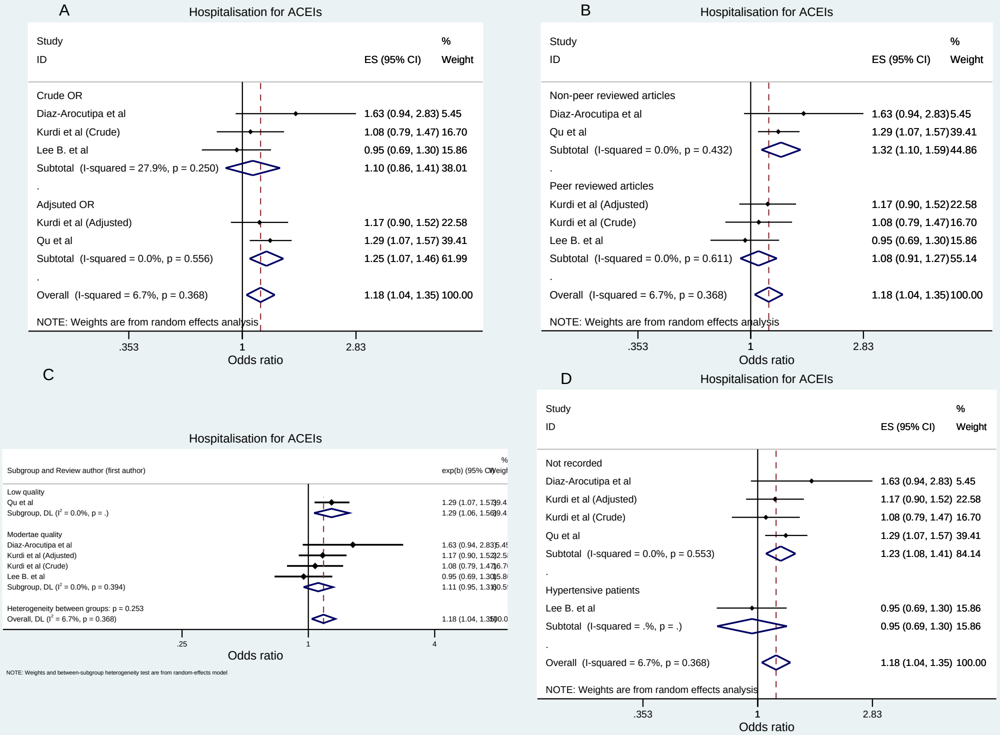

### Supplementary file 9B

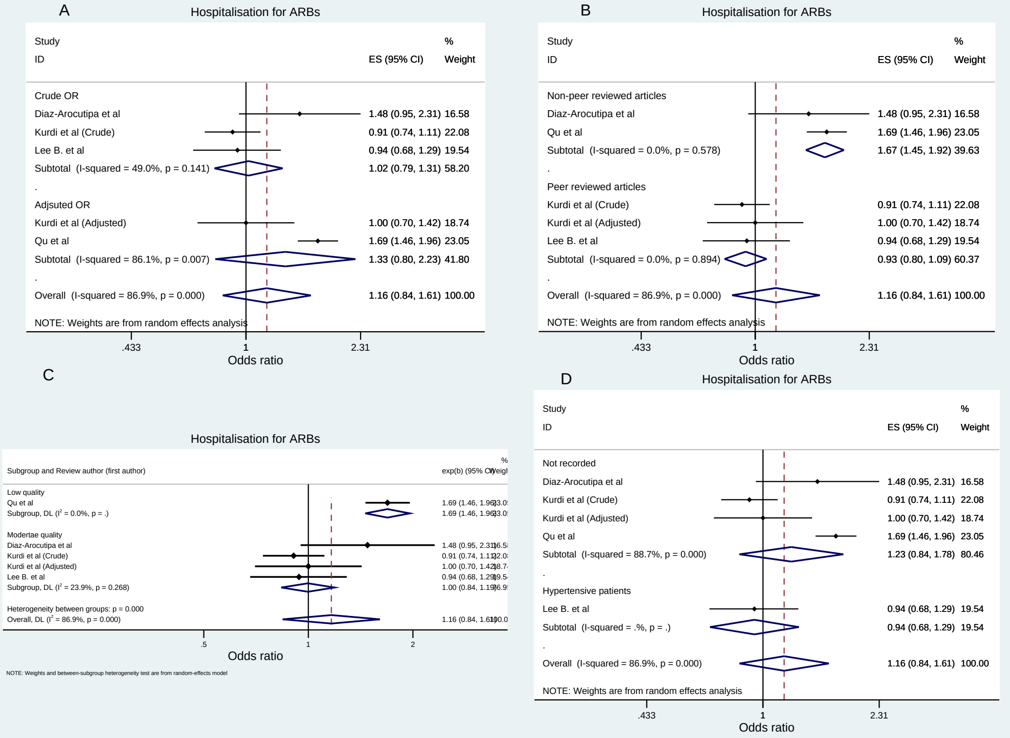

### Supplementary file 10

Supplementary file 10. Publication bias funnel plot for the outcomes with >=10 studies

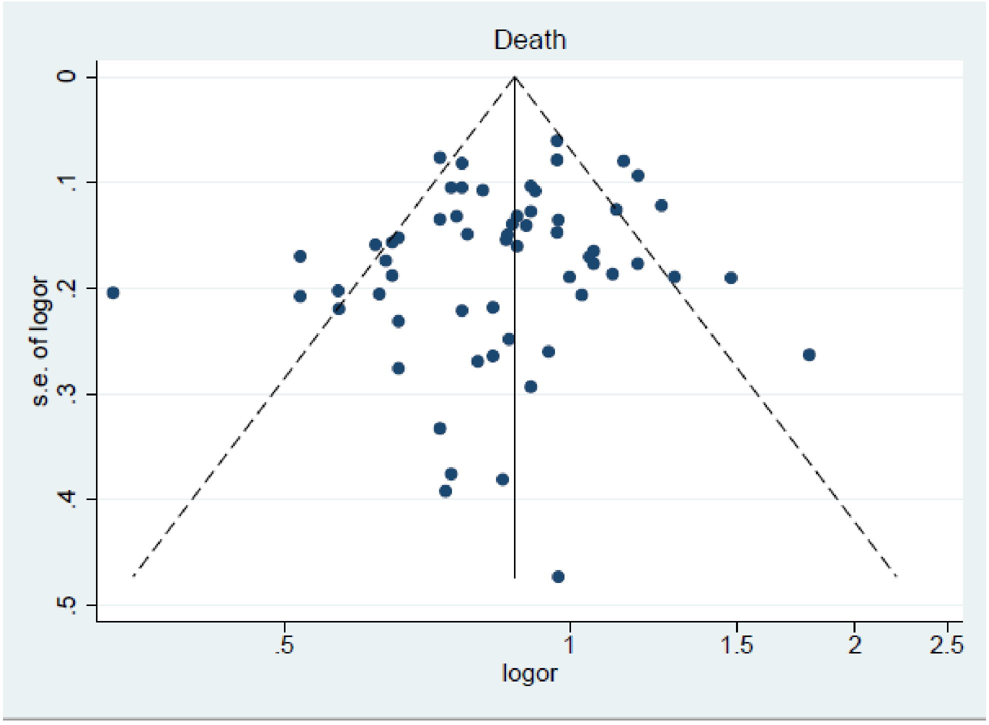


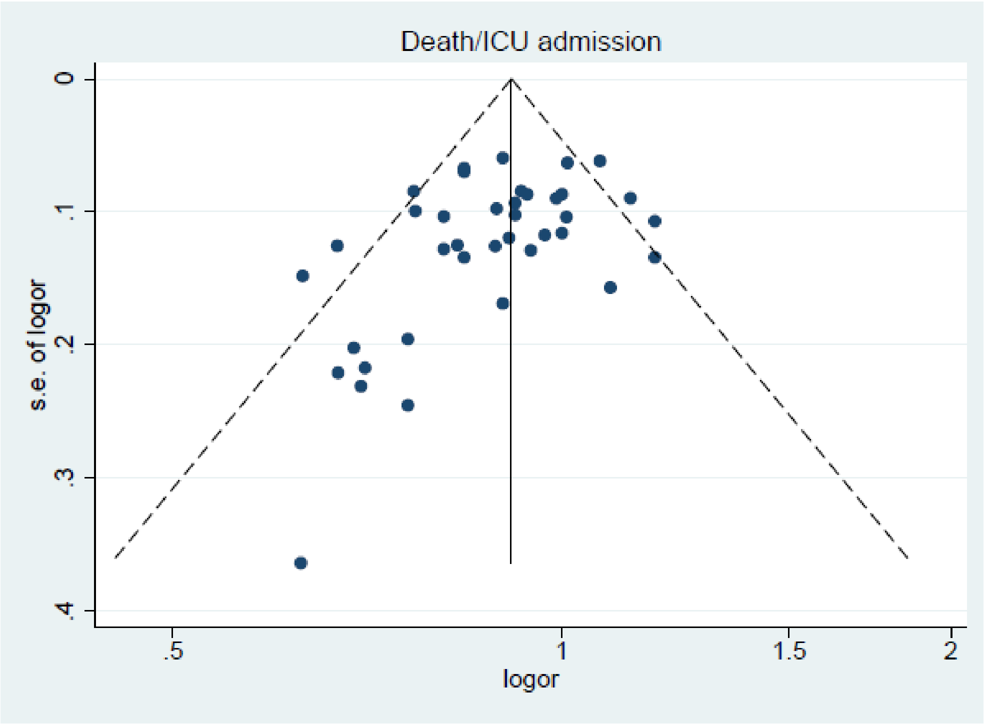

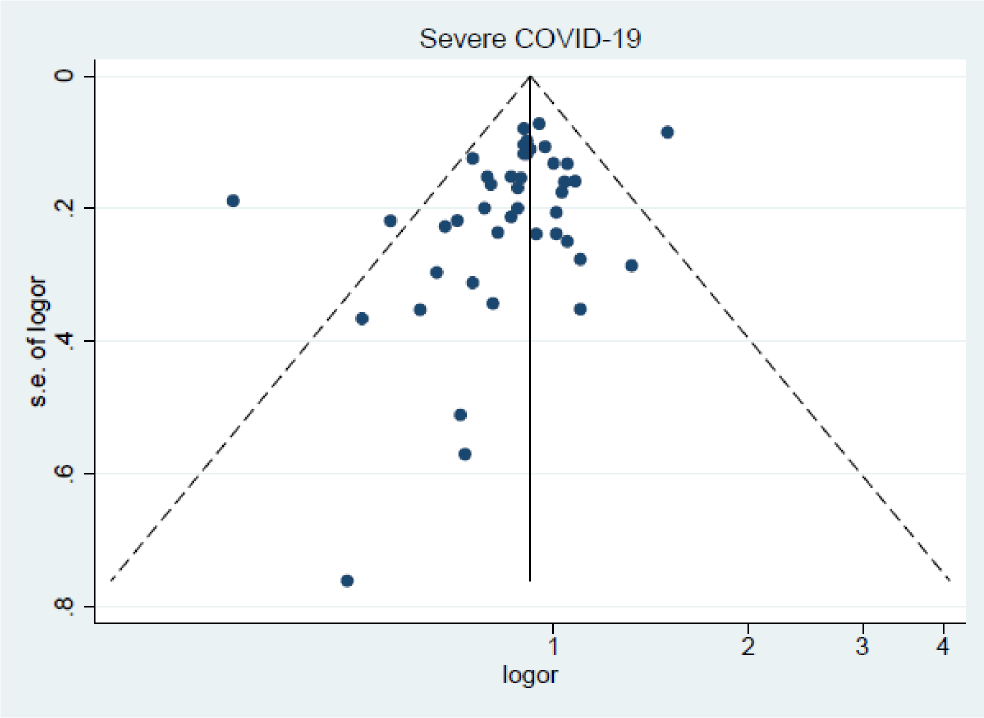

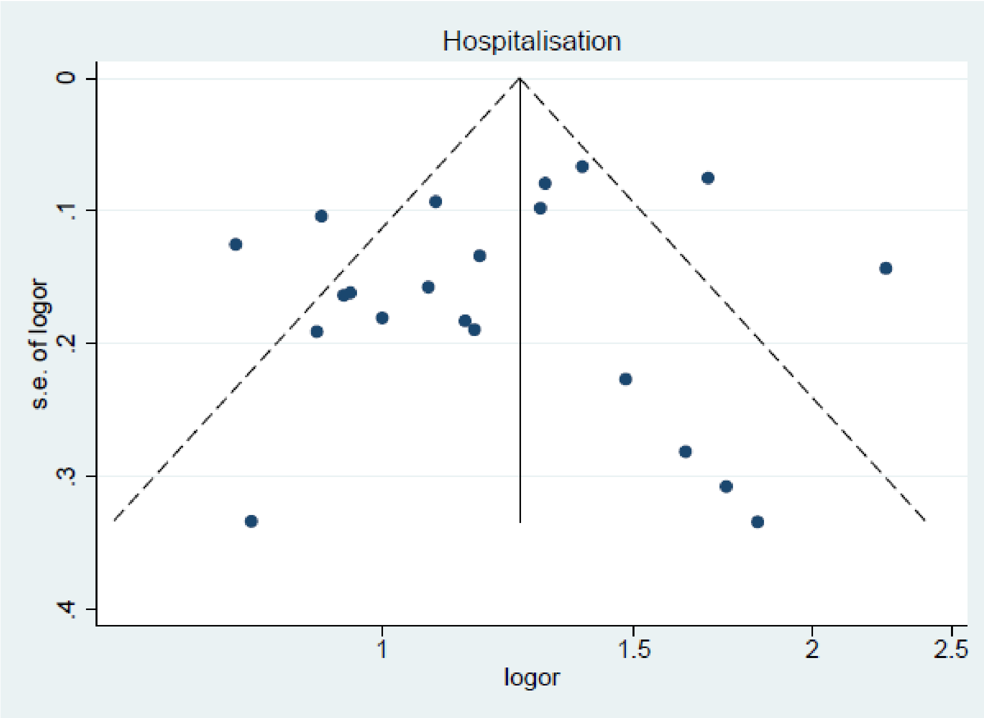

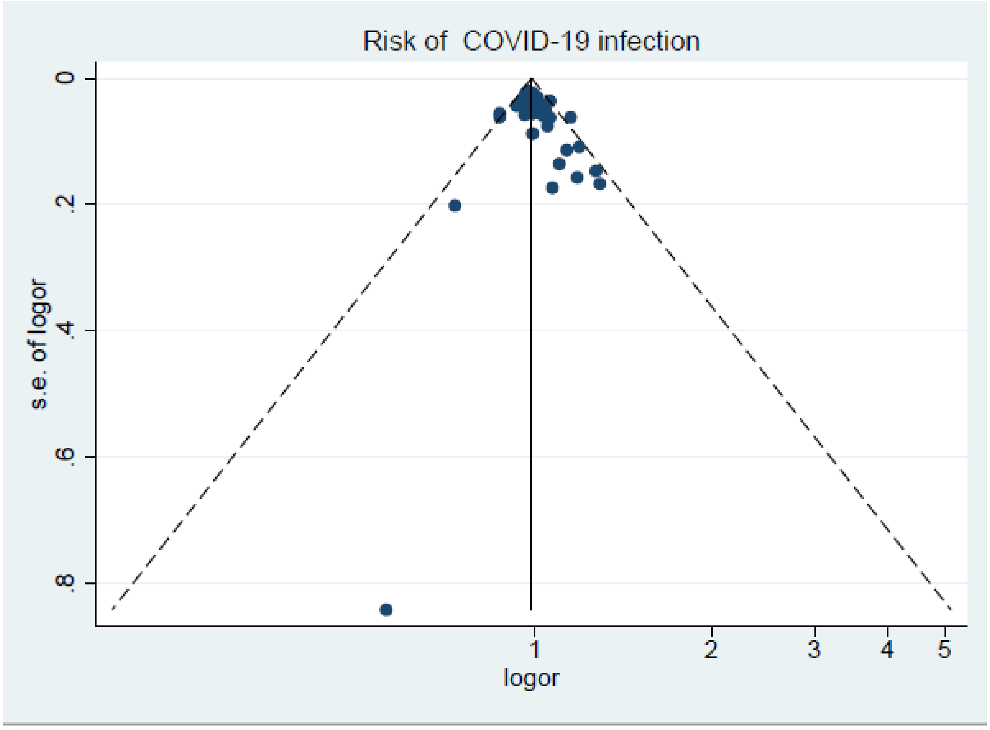

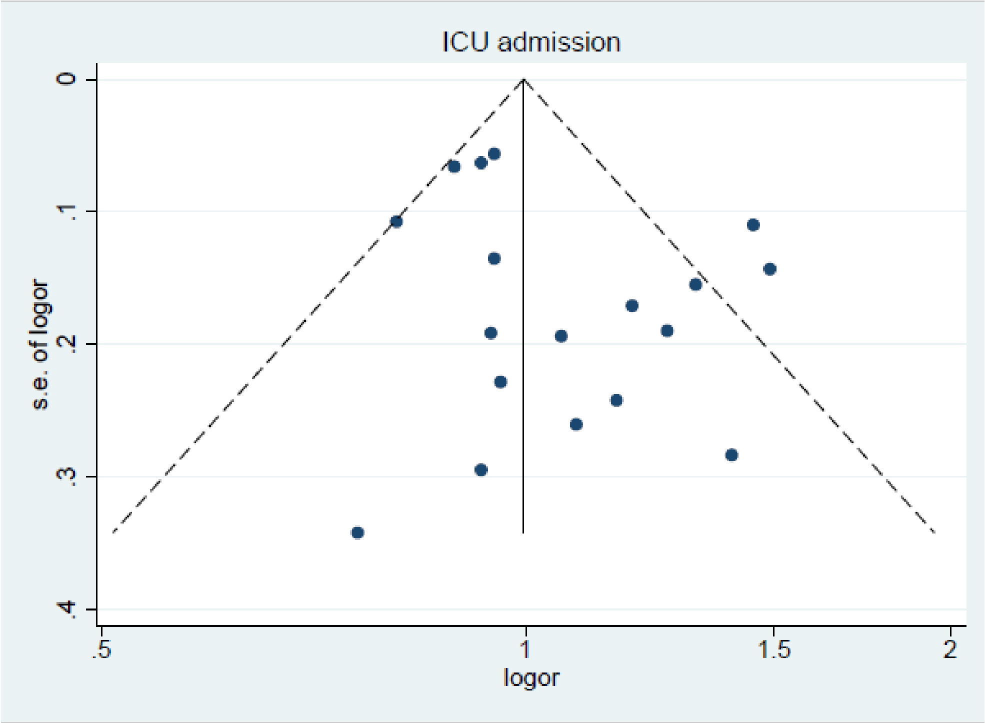

### Supplementary file 11

Supplementary file 11. Results of the influential analyses

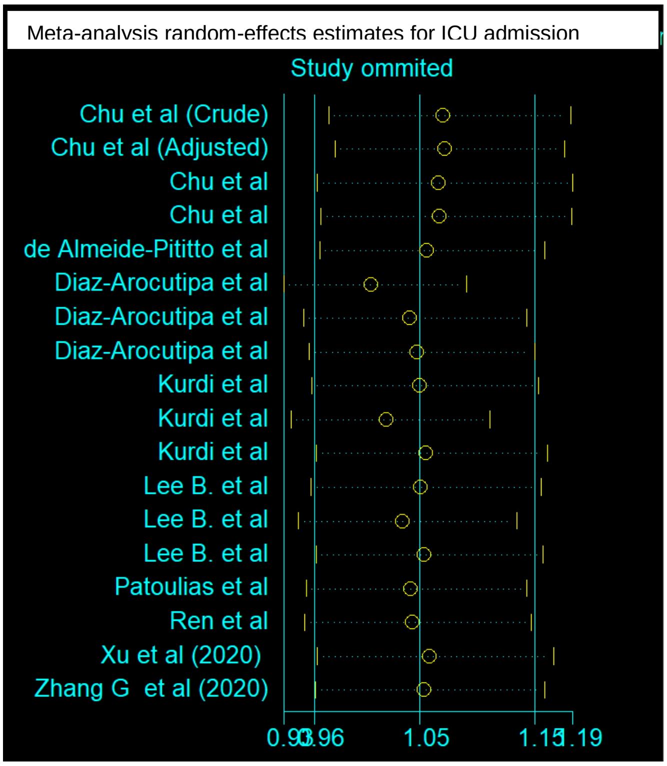


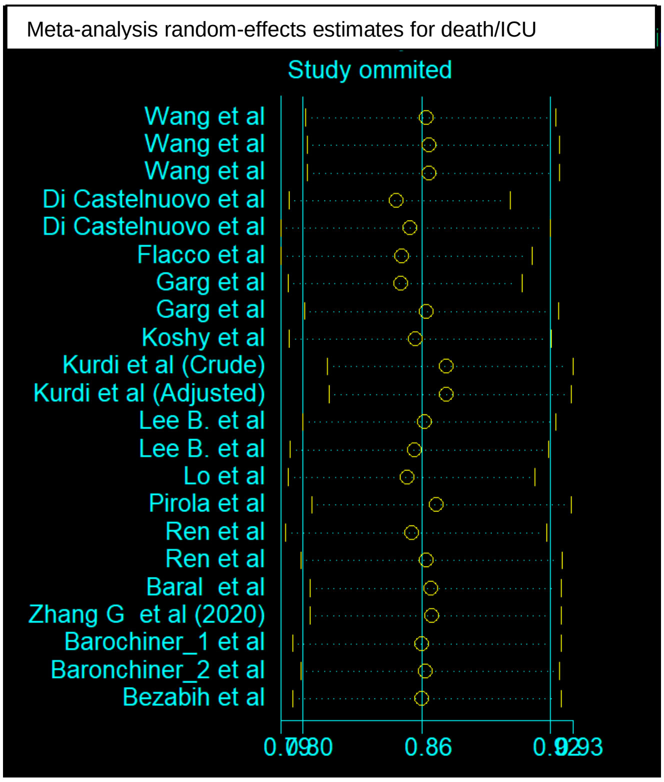

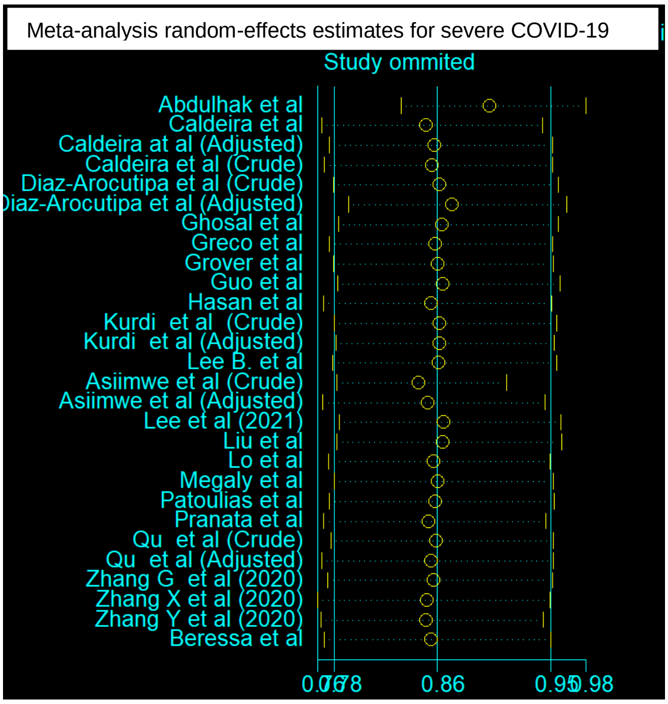

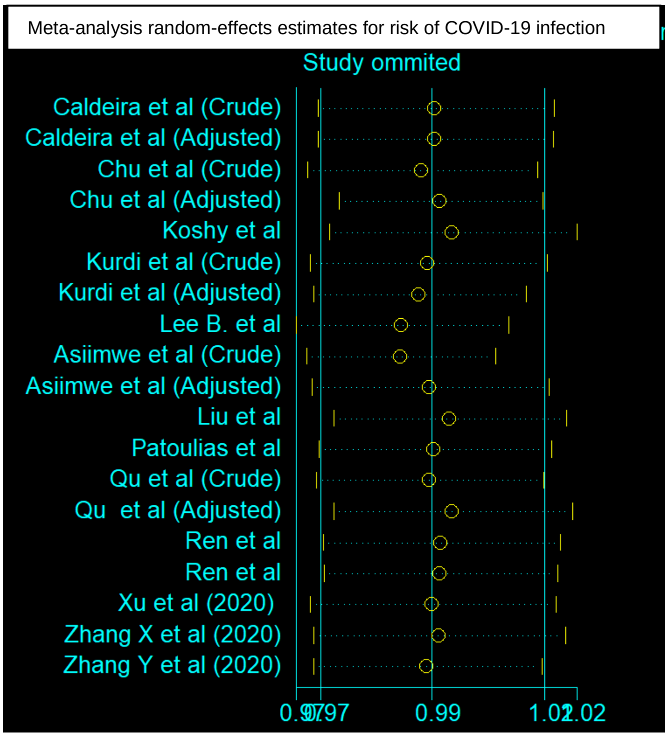

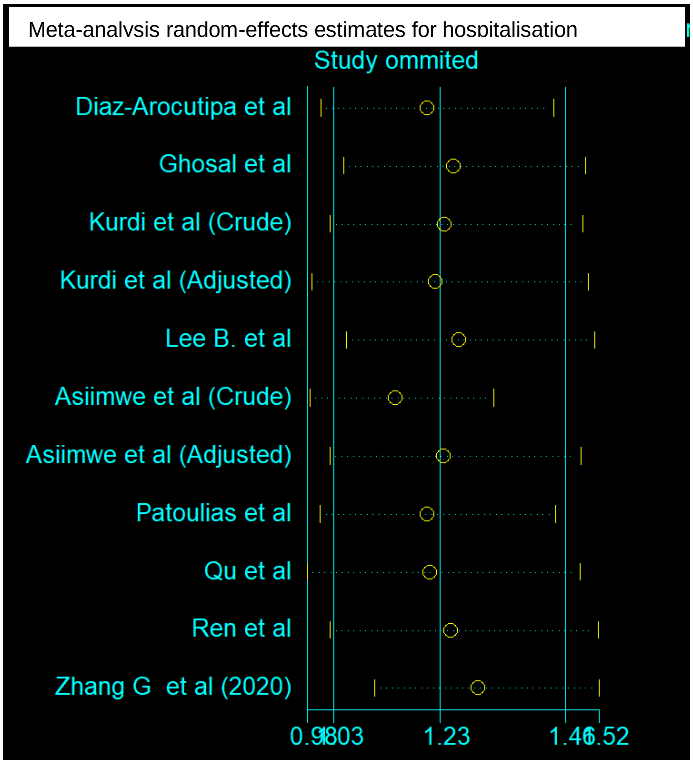
