## Supplementary file 3 for "An umbrella review and meta-analysis of the use of renin-angiotensin system drugs and COVID-19 outcomes: what do we know so far?"

**Review details (n=47)**

| **Review** | **Aim/objective** | **Peer review status** | **Databases searched** | **Month search conducted** | **Did review included pre-prints?** | **Study designs included in review** | **Studies included in review (n)** | **Did review adjust for retracted studies?** |
| --- | --- | --- | --- | --- | --- | --- | --- | --- |
| Abdulha (2020) | Therefore, herein we aimed to systematically review the exiting literature and perform a meta-analysis of the available studies examining the role of ACEIs/ARBs in the outcome of COVID-19. | Not peer reviewed | Medline, ClinicalTrial.org, MedRxiv | May-20 | Yes | All study types and design | 9 | No |
| Alamer (2020) | We re-estimated the Zhang et al. [5] pooled estimates for mortality in Covid-19 patients and mortality in Covid-19 patients with antihypertensive indication after excluding the Mehra et al. [1] data. | Unclear if peer reviewed- letter to the editor | Not reported | Not reported | No | All study types and design | Not reported | Yes |
| Asiimwe (2020) | To provide more comprehensive and up-to-date evidence, we have conducted a systematic review and meta-analysis to evaluate all the current evidence on the influence of cardiovascular drugs on COVID-19 clinical outcomes. Due to the rapidly evolving nature of this field, we will periodically update this baseline review for up to two years to reflect emerging evidence. | Not peer reviewed | MEDLINE, and Liverpool’s DISCOVER platform (which links, through EBSCOhost, to sources from >500 databases including MEDLINE, Google Scholar, Scopus, the Web of Science and Cochrane Central Register of Controlled Trial libraries), bioRxiv and medRxiv, Text S1, COVID-19 Clinical Trials registry, World Health Organization database of COVID-19 publications, ClinicalTrials.gov, International Clinical Trials Registry Platform | Jul-20 | Yes | Observational (e.g. cohorts/case-series and case-control studies) and interventional (e.g. randomised controlled trials) studies | 178 | No |
| Baral (2020) | We, thus, conducted an up-to-date systematic review and meta-analysis of RAAS blockers in patients with COVID-19. | Peer reviewed | PubMed, Embase, Medrxiv | May-20 | Yes | All study types and design | 20 | Yes |
| Barochiner -a (2020) | Therefore, in this article, we conducted a systematic literature search to determine a possible association between the use of ACEI/ARB in hypertensives who become infected with COVID-19 and the progression of the disease to severe forms or death. | Peer reviewed | Medline, Cochrane Library, Google Scholar, medRxiv, bioRxiv | Jun-20 | Yes | Comparative studies - RCTs or non-RCTs | 18 | No |
| Baronchiner - b (2020) | Therefore, in this article, we conducted a systematic literature search to determine a possible association between the use of ACEI/ARB and the progression of COVID-19 to severe forms or death. | Not peer reviewed | Medline, Cochrane Library, Google Scholar, MedRxiv and BioRxiv, | May-20 | Yes | Comparative studies - RCTs or non-RCTs | 15 | No |
| Beressa (2020) | This study aimed to systematically synthesis the evidence on the effect of RAAS inhibitors on the outcome of COVID_19 patients with hypertension. | Not peer reviewed | PubMed, Cochrane Central Register of controlled trials, clinical trial.gov, CINAHIL, Google scholar | Jun-20 | Yes | RCTs (including cluster RCTs), observational studies, and prospective, retrospective comparative cohort studies and case-control studies | 9 | No |
| Bezabih (2020) | Therefore, our primary objective was to perform a systematic review and meta-analysis to estimate the overall risk of poor COVID-19 outcomes in patients receiving RAAS inhibitors compared to non-RAAS inhibitor antihypertensive agents. As secondary objective, we also compared the of risk developing poor clinical outcomes between specific classes of antihypertensives (ACEIs, ARBs, beta-blockers (BBs), calcium channel blockers (CCBs), and thiazides). | Not peer reviewed | PubMed, EMBASE | May-20 | No | Cohort (prospective or retrospective) studies, case series and editorials/letters. | 7 | No |
| Cai (2020) | This systematic review and meta-analysis sought to quantify the effect of ACEI/ARB on the risks of in-hospital mortality in COVID-19 patients. | Peer reviewed | PubMed, EMBASE, clinicaltrials.gov, Google Scholar | May-20 | No | All study types and design | 9 | No |
| Caldeira (2020) | In this systematic reviewwe aimed to assess the risk of infection by SARS-CoV-2 and the riskof mortality or respiratory complications in patients with symp-tomatic disease of SARS-CoV-2 (COVID-19) related to previoususe of ACEi or ARBs. | Peer reviewed | MEDLINE, CENTRAL, PsycINFO, Web of Science Core collection, medRxiv | Jun-20 | Yes | All controlled studies (RCTs, cohort/nested case-control/case-control studies) | 27 | No |
| Chan (2020) | In this study, we aimed to clarify the association between the use of ACE inhibitors or ARBs and the risk of SARS-CoV-2 infection. | Peer reviewed | MEDLINE (Ovid), CINAHL (Ovid), Cochrane Database of Systematic Reviews, PubMed, Embase, medRxiv, Cnki.net, WHO database of COVID publications, ClinicalTrials.gov | Jun-20 | Yes | All study types and design | 7 | No |
| Chu (2020) | A systematic literature search was conducted to identify studies investigating the association between ACEIs or ARBs and pneumonia or COVID-19 | Peer reviewed | PubMed, Embase (OVID), the Cochrane Central Register of Controlled Trials (CENTRAL), ClinicalTrials.gov | Sep-20 | No | RCTs, cohort studies, case-control studies, nested case-control studies, case-crossover studies | 83 | Unclear |
| de Almeide-Pititto (2020) | The aim of this study is to evaluate the association of diabetes, hypertension, cardiovascular disease and ACEI/ ARBs exposure with severity–intensive care unit treatment or mechanical ventilation necessity or O2 saturation | Peer reviewed | PubMed, Cochrane Library, SciELO | May-20 | No | Observational studies (cross-sectional, self-controlled case series, retrospective cohort studies) | 40 | No |
| Di Castelnuovo (2020) | Against this controversial background, in March 2020, we launched a large multicenter study in Italy aimed at investigating the role of RAAS inhibitors in COVID-19 patients. We here present the findings of this collaborative project, supported by a set of related meta-analyses | Peer reviewed | MEDLINE, EMBASE, Web of Science, Cochrane Central Register of Controlled Trials | Jul-20 | No | All study types and design | 19 | No |
| Diaz-Arocutipa (2020) | Recently, several studies that evaluated the effect of RAS inhibitors on COVID-19 have been published. Therefore, we performed a systematic review and meta-analysis to evaluate the association between ACEIs or ARBs use and clinical outcomes in COVID-19 patients. | Not peer reviewed | PubMed, Embase, Scopus, Web of Science, Cochrane Central Register of Controlled Trials, medRxiv, SSNR, clinicaltrials.gov | May-20 | Yes | RCTS and observational studies | 40 | No |
| Flacco (2020) | We thus carried out a meta-analysis to summarise the existing evidence from adjusted analyses on the association between RAAS inhibitors and COVID-19. | Peer reviewed | MEDLINE, S+B32+F20 | May-20 | Yes | Cohort or case-control design | 10 | Yes |
| Garg (2020) | Accordingly, we sought to study the cumulative evidence for association of ACEI/ARB use with risk of mortality and severe illness with Covid-19. | Not peer reviewed | MEDLINE, EMBASE | May-20 | No | All study types and design | 15 | No |
| Ghosal (2020) | We therefore did a meta-analysis of the studies reported till 21.04.2020 on the outcomes in patients with COVID-19 on ACEi/ARBs to strengthen the evidence base by determining the benefit-risk ratio of these medications in patients with COVID-19. | Not peer reviewed | Cochran Library, PubMed, Embase, CT.gov, medRxiv | Apr-20 | Yes | All study types and design (except case reports) | 6 | No |
| Greco (2020) | Accruing evidence is available that does not show a higher risk with ACEIs/ARBs in patients with COVID-19.5 However, available studies have limited statistical power to detect signals of increased mortality.6 In this context, a meta-analysis provides more precise estimates of the direction and strength of association between the use of ACEIs/ARBs and prognosis. Few meta-analyses have been published that do not show an increase in death with ACEIs/ARBs. However, new studies are now available that suggest the opportunity for an update.7,8 | Unclear if peer reviewed - correspondence | PubMed, Scopus, and web sources | Jun-20 | No | All study types and design | 14 | No |
| Grover (2020) | Therefore, the medical literature was systematically reviewed, and a meta-analysis was performed of the current clinical studies evaluating the safety and efficacy of ACEs and ARBs in COVID-19 patients. | Peer reviewed | MEDLINE, Google Scholar, MedRxiv, BioRxiv | May-20 | No | All study types and design | 16 | No |
| Guo (2020) | Thus, we performed a meta-analysis of the current studies to explore whether the use of ACE inhibitor/ARB was associated with disease severity and mortality in COVID-19 patients with hypertension. | Unclear if peer reviewed - research letter | Pubmed, Web of Science, Scopus, Embase | May-20 | No | All study types and design | 9 | No |
| Hasan (2020) | we systematically reviewed the data from available studies to date to critically examine the association of RAS inhibitors use with mortality and disease severity in COVID-19. | Peer reviewed | PubMed, Google Scholar, and two preprint repositories (medRxiv and SSRN) | Aug-20 | Yes | Observational (prospective or retrospective) studies | 59 | No |
| Kashour (2020) | Therefore, herein we aimed to systematically review the exiting literature and perform a meta-analysis of the available studies examining the role of ACEIs/ ARBs in the outcome of COVID-19. | Peer reviewed | Ovid MEDLINE(R) and Epub Ahead of Print, In-Process and Other Non-Indexed Citations and Daily, Ovid Embase, Ovid Cochrane Central Register of Controlled Trials, Ovid Cochrane Database of Systematic Reviews, Web of Science, Scopus, Medrxiv.org, Google scholar | Jun-20 | No | All study types and design | 9 | No |
| Kerneis (2020) | The aims of this review were to report the updated evidence to guide physicians’ clinical decision making, to present a pooled analysis of the published observational studies evaluating all-cause mortality of patients with COVID-19 according to treatment with RAS blockers and to provide the latest information on ongoing clinical research related to RAS blocker treatments in patients with COVID-19. | Peer reviewed | Pubmed, Embase | Jul-20 | No | All study types and design | 21 | No |
| Koshy (2020) | With the release of new evidence, we performed a systematic review and meta-analysis of studies to assess whether RAS inhibitors were associated with a risk of a positive test for COVID-19 as well as the likelihood of severe illness or mortality due to the disease. | Peer reviewed | MEDLINE, PubMed and EMBASE | May-20 | No | All study types and design | 7 | Yes |
| Kurdi (2020) | Consequently, we sought to undertake an updated and comprehensive evaluation of effect of ACEIs/ARBs use on all reported COVID-19-related outcomes, including exploration of any class differences, through a systematic review of the literature coupled with a meta-analysis. | Peer reviewed | Embase, Medline (OVID), Scopus, Cochrane library, and medRxiv | May-20 | Yes | All study types and design | 27 | No |
| Lee (2021) | To help resolve this issue, we conducted a systematic review and meta-analysis on recently published studies to investigate the effects of ACEI/ARBs on the risk of mortality and severe disease in patients diagnosed with COVID-19. | Peer reviewed | Medline, Embase, Cochrane Central Register of Controlled Trials, MedRxiv, Social Science Research N etwork and Peer J databases | May-20 | Yes | Any human clinical comparative study (except for those with a cross-sectional design) | 20 | No |
| Lee b (2020) | It is therefore important to understand whether the type of RAS blocker matters in relation to CoViD-19, especially as these drugs can generally be used interchangeably in clinical practice, and patients could be switched to the safer alternative. Consequently, we have undertaken a systematic review and meta-analysis of non-randomized, observational, studies to address these questions which are relevant to millions of patients with hypertension, diabetes, cardiovascular, and renal disease treated with RAS blockers worldwide. | Peer reviewed | PubMed, EMBASE, Web of Science, Google Scholar, medRxiv, SSRN preprint | Aug-20 | Yes | Non-randomized observational studies (cohort, case–control, case series) | 92 | No |
| Liu (2020) | in this article, we will review the current evidence and assess the clinical prognosis of COVID-19 patients with or without hypertension treated with ACEs/ARBs | Peer reviewed | PubMed, Embase, medRxiv, bioRxiv | May-20 | Yes | RCTs, case-control studies, cohort studies | 11 | No |
| Lo (2020) | Therefore, this systematic review and meta-analysis sought to investigate the impact of ACEI/ ARB usage on mortality and disease severity in patients positive for COVID-19 among different locations worldwide. | Peer reviewed | PubMed, CINAHL, pre-print servers | Jul-20 | Yes | Prospective and retrospective studies (excluding single case reports and case series) | 21 | Yes |
| Megaly (2020) | Data on the outcomes of ACEIs/ARBs in patients with COVID-19 is limited; we, therefore, performed the current meta-analysis to fill that gap in the literature | Unclear if peer reviewed - letter | Embase, Medline, Cochrane databases | May-20 | No | All study types and design | 4 | No |
| Nunes (2020) | The aim of the study was to use currently available data to tentatively evaluate if a relation exists between ACEi use and patient mortality in this context. | Peer reviewed | Medline(PubMed), ISI Web of Knowledge, medRxiv | Jun-20 | Yes | All study types and design (except case reports) | 7 | Yes |
| Patoulias (2020) | Towards this end, we conducted a systematic review and meta-analysis of clinical studies assessing the association of RAS inhibitors with COVID-19 infectivity, severity (need for hospitalization, admission to intensive care unit (ICU)), and mortality. We also sought to unveil potential differences between angiotensin-converting enzyme (ACE) inhibitors and angiotensin receptor blockers (ARBs) in these outcomes, as well as potential disparities in different continents. | Peer reviewed | PubMed, Cochrane Central Register of Controlled Trials (CENTRAL), clinicaltrials.gov, medRxiv | May-20 | Yes | RCTs or observational studies | 25 | Yes |
| Pirola (2020) | the primary objective of the current study is to provide a quantitative estimation of the effect of ACEIs and/or ARBs, alone or ACEIs/ARBs (undistinct drug) on COVID-19 severe clinical illness in patients with arterial hypertension by a meta-analysis | Peer reviewed | PubMed, Ovid-Medline, Google Scholar | May-20 | No | Observational studies | 16 | No |
| Pranata (2020) | Therefore, to investigate the association between the administration of ACEI/ARB and the outcomes of hypertensive COVID-19 patients, a systematic review and meta-analysis were performed. | Peer reviewed | PubMed, EuropePMC, PRoQuest, Cochrane Central Databases, Google Scholar, preprint servers | Jun-20 | Yes | All study types and design | 15 | Yes |
| Qu (2020) | The goal of this meta-analysis is to clarify the effects of ACEIs and/or ARBs on the infection risk, severity, and mortality of COVID-19, hoping to shed more light in the prevention and treatment of the current ongoing pandemic. | Not peer reviewed | PubMed, Web of Science, Cochrane Library, China National Knowledge Infrastructure databased, medRxiv | May-20 | Yes | Cohort or case-control design | 22 | Yes |
| Ren (2020) | To this end, we designed a meta-analysis to address whether there is an association of antihypertensive medications and the incidence and severity of COVID-19 | Peer reviewed | PubMed, MEDLINE, The Cochrane Library, the Cochrane Central Register of Controlled Trials (CENTRAL), ClinicalTrials.gov, medRxiv | Jul-20 | Yes | RCTs, case-control studies, cohort studies | 53 | No |
| Ssentongo (2020) | In this this systematic review and meta-analysis, the authors aim to delineate the association of RAAS-inhibitors use and mortality in patients with COVID-19 | Peer reviewed | PubMed (MEDLINE), SCOPUS, OVID, Cochrane Library databases, medRxiv | Sep-20 | Yes | All study types and design (except case reports and case series with <10 patients) | 14 | No |
| Tleyjeh (2020) | Therefore, we systematically reviewed the literature and performed a meta-analysis of the association between prior use of ACEIs and ARBs and development of COVID-19 | Peer reviewed | Ovid MEDLINE(R) and Epub Ahead of Print, In-Process & Other Non-Indexed Citations and Daily, Ovid Embase, Cochrane Central Register of Controlled Trials, Cochrane Database of Systematic Reviews, Web of Science, Scopus. | Jun-20 | Yes | All study types and design | 10 | No |
| Usman (2020) | Thus, the effect of RAAS inhibitors on susceptibility and prognosis of COVID-19 continues to be the subject of much debate.1 Individual observational studies in the area have yielded equivocal results; hence, we sought to conduct a meta-analysis of all available data to provide greater insight. | Unclear if peer reviewed - readers comment | PubMed and Scopus | May-20 | No | All study types and design | 8 | No |
| Wang (2020) | We conducted this updated meta‐analysis not only to investigatethe effects of ACEI/ARB on mortality as well as other outcomevariables such as admission to intensive care unit (ICU) and rate of ventilatory support in COVID‐19 patients with hypertension, but also to explore potential modulators. | Peer reviewed | PubMed, Cochrane Library, Embase, medRxiv, bioRxiv | Oct-20 | Yes | Observational studies (case-control or cohort design) | 26 | No |
| Xu (2020) | In this systematic review, we aim to provide a comprehensive summary of current evidence to answer 2 major clinical questions: (1) Does prior treatment with ACEIs/ARBs alter the susceptibility of SARS-CoV-2 infection in the general population and (2) Does prior treatment with ACEIs/ARBs affect the risk of mortality and severe outcomes in patients with COVID-19? This information is critical to guide the evidence-based management of ACEIs/ARBs in patients during the COVID-19 pandemic. | Peer reviewed | MEDLINE (via Ovid), Embase, The Cochrane Central Register of Controlled Trials (CENTRAL), China National Knowledge Infrastructure (CNKI), Wanfang database, SinoMed, World Health Organization (WHO) COVID-19 database (global literature on coronavirus disease), and the Cochrane COVID-19 study register, preprint servers (MedRxiv, BioRxiv, SSRN), | May 2020, updated July 2020 | Yes | RCTs, nonrandomized clinical trials, cohort studies, and case-control studies | 49 | Yes |
| Xue (2020) | Therefore, we performed a meta-analysis on the effects of ACEI/ARB on disease symptoms and laboratory tests in hypertensive patients infected with COVID-19 virus and those who did not use ACEI/ARB. | Peer reviewed | Pubmed, Embase, EuropePMC, medRxiv (https://www.medrxiv.org), SSRN (https://www.ssrn.com), and CNKI databases | Apr-20 | No | All study types and design | 6 | No |
| Yokoyama (2020) | Therefore, investigation of the effect of ACEI/ARB use on patients with COVID‐19 is still warranted, hence we conducted two meta‐analyses to compare mortality and susceptibility of SARS‐CoV‐2 infection between patients treated and those not treated with ACEI and/or ARB. | Peer reviewed | Pubmed and EMBASE | Jun-20 | No | Observational studies | 17 | No |
| Zhang G (2020) | It is important to determine whether the RAAS inhibitor (ACEI/ ARB) use is associated with increased risks of contracting COVID‐19 and developing severe disease. Thus, we performed a meta‐analysis of the available studies to explore whether the use of RAAS inhibitors was associated with severe disease and mortality in COVID‐19 patients. | Peer reviewed | PubMed, Cochrane Library, Web of Science, EMBASE, medRxiv, and bioRxiv databases | Oct-20 | Yes | Observational cohort studies | 25 | No |
| Zhang X (2020) | Thus, the aims of this work were as follows: (1) to determine whether ACEI/ARB use is associated with an increase in likelihood of viral infectivity; (2) to investigate whether there are differences in severity and mortality between ACRI/ARB users and non- ACRI/ARB users; (3) to evaluate, in particular, whether ACEI/ARB exposure was associated with a lower risk of mortality when compared to non-ACEI/ ARB antihypertensive drug exposure. Our findings provide vital guidance for current clinical work on the prevention and treatment of COVID-19 infection. | Peer reviewed | PubMed, Embase, and Cochrane Library | May-20 | No | Case-control, case-crossover, self-controlled case series, or cohort study | 12 | No |
| Zhang Y (2020) | We therefore undertook a systematic review and meta-analysis of all existing clinical data to provide the most definitive report to date on the association between RAS inhibitors and the susceptibility to SARS-CoV-2 infection (18 studies), the severity of the resulting disease (29 studies) and mortality (30 studies) due to COVID-19. | Not peer reviewed | PubMed, Web of Science, OVID Embase, MedRxiv, BioRxiv | Jul-20 | Yes | All study types and design | 58 | No |

ACEI = Angiotensin Converting Enzyme Inhibitors; ARB = Angiotensin Receptor Blockers; RAAS = Renin–angiotensin–aldosterone system; RCT = Randomised Control Trial
