## Supplementary file 4 for "An umbrella review and meta-analysis of the use of renin-angiotensin system drugs and COVID-19 outcomes: what do we know so far?"

**Details of all meta-analyses (n=213)**

| **Review** | **Outcome** | **Studies included in meta-analysis (n)** | **Exposure** | **Ratio used** | **Unadjusted Ratio (95% CI)** | **I2 (%)** | **Adjusted Ratio (95% CI)** | **I2 (%)** | **Total patients (n)** | **Patients COVID-19 positive (n)** | **Patients on ACEI / ARB / RAAS inhibitors (n)** | **Criteria for COVID-19 diagnosis** | **Location of patient cohort** | **Population character-istics** | **Other comorbidities** | **Age - years** | **Gender - % male** |
| --- | --- | --- | --- | --- | --- | --- | --- | --- | --- | --- | --- | --- | --- | --- | --- | --- | --- |
| Abdulha (2020) | Severe disease | 5 | ACEIs/ARBs | Odds ratio | 0.32 (0.22-0.46) | 32% | NR | NR | 11,242 | 11,242 | NR / NR / 1594 | NR | NR | NR | NR | NR | 6,740 (60%) estimated |
| Abdulha (2020) | Mortality | 4 | ACEIs/ARBs | Odds ratio | 0.33 (0.22-0.49) | 0% | NR | NR | 11,183 | 11,183 | NR / NR / 1591 | NR | NR | NR | NR | NR | 6,708 (60%) estimated |
| Alamer (2020) | Mortality | 5 | ACEIs/ARBs | Odds ratio | 0.66 (0.42-1.04) | 30% | NR | NR | NR | NR | NR / NR / NR | NR | NR | All patients on ACEIs / ARBs for HTN | NR | NR | NR |
| Alamer (2020) | Mortality | 7 | ACEIs/ARBs | Odds ratio | 0.77 (0.5-1.19) | 34% | NR | NR | NR | NR | NR / NR / NR | NR | NR | NR | NR | NR | NR |
| Asiimwe (2020) | All-cause mortality | 40 | ACEIs/ARBs | Odds ratio | 1.25 (0.98-1.58) | 85% | 0.86 (0.64-1.15) | 4% | 43,099 | 43,099 | N/A / N/A / 11340 | NR | NR | NR | NR | NR | NR |
| Asiimwe (2020) | COVID-19 infection risk | 24 | ACEIs/ARBs | Odds ratio | 1.15 (1.02-1.3) | 93% | 1.01 (0.93-1.1) | 0% | 1,362,182 | 1,362,182 | N/A / N/A / 275913 | NR | NR | NR | NR | NR | NR |
| Asiimwe (2020) | Severe disease | 60 | ACEIs/ARBs | Odds ratio | 1.5 (1.27-1.77) | 81% | 1.04 (0.76-1.42) | 65% | 45,394 | 45,394 | N/A / N/A / 9673 | NR | NR | NR | NR | NR | NR |
| Asiimwe (2020) | Hospitalis-ation | 20 | ACEIs/ARBs | Odds ratio | 2.25 (1.7-2.98) | 91% | 1.16 (0.8-1.68) | 53% | 27,413 | 27,413 | N/A / N/A / 4838 | NR | NR | NR | NR | NR | NR |
| Asiimwe (2020) | Length of hospitaliz-ation | 11 | ACEIs/ARBs | Other - Mean difference (days) with SI | -0.45 (-1.33-0.43) | 30% | NR | NR | 2,510 | 2,510 | N/A / N/A / 742 | NR | NR | NR | NR | NR | NR |
| Baral (2020) | Death/critical outcomes | 16 | ACEIs/ARBs | Odds ratio | 0.671 (0.435-1.034) | 9434% | NR | NR | 16099 | 16099 | N/A / N/A / 5996 | NR | NR | NR | NR | NR | NR |
| Baral (2020) | Mortality | 12 | ACEIs/ARBs | Odds ratio | 0.857 (0.634-1.16) | 6479% | NR | NR | 7492 | 7492 | N/A / N/A / 1985 | NR | NR | NR | NR | NR | NR |
| Baral (2020) | Death/critical outcomes | 8 | ARBs | Odds ratio | 0.946 (0.735-1.218) | 5999% | NR | NR | 11308 | 11308 | N/A / 2540 / N/A | NR | NR | NR | NR | NR | NR |
| Baral (2020) | Death/critical outcomes | 8 | ACEIs | Odds ratio | 1.008 (0.822-1.235) | 3826% | NR | NR | 11137 | 11137 | 2479 / N/A / N/A | NR | NR | NR | NR | NR | NR |
| Baro-chiner (2020) | Composite outcome (death, admission to ICU, mechanical ventilation requirement or progression to severe or critical pneumonia) | 15 | ACEIs/ARBs | Risk ratio | 0.81 (0.63-1.04) | 82.00% | NR | NR | 21614 | 21614 | NR / NR / 6477 | NR | NR | NR | NR | NR | 12752 |
| Baro-chiner (2020) | Composite of death, admission to ICU, mechanical ventilation, progression to severe / critical pneumonia | 18 | ACEIs/ARBs | Risk ratio | 0.84 (0.73-0.95) | 65% | NR | NR | 17,311 | 17,311 | N/A / N/A / 8328 | NR | NR | All HTN patients | NR | NR | NR |
| Beressa (2020) | Mortality | 7 | ACEIs/ARBs | Risk ratio | 0.73 (0.63-0.85) | 0.00% | NR | NR | 3572 | 3572 | NR / NR / 1207 | NR | NR | All HTN patients | NR | NR | NR |
| Beressa (2020) | Disease severity | 9 | ACEIs/ARBs | Risk ratio | 0.92 (0.74-1.14) | 64.00% | NR | NR | 3,823 | 3,823 | NR / NR / 1416 | NR | NR | All HTN patients | NR | NR | NR |
| Beressa (2020) | Long term hospitalis-ation | 2 | ACEIs/ARBs | Other - Weighted mean difference (days) | -2.33 (-5.6-0.75) | 0.00% | NR | NR | 201 | 201 | NR / NR / 71 | NR | NR | All HTN patients | NR | NR | NR |
| Bezabih (2020) | Poor patient outcomes | 3 | ACEIs | Odds ratio | 0.77 (0.63-0.93) | NR | NR | NR | 5,539 | 5,539 | NR / NR / 1366 | NR | NR | All HTN patients | NR | NR | NR |
| Bezabih (2020) | Poor patient outcomes | 7 | ACEIs/ARBs | Odds ratio | 0.84 (0.73-0.96) | NR | NR | NR | 6,997 | 6,997 | NR / NR / 2858 | NR | NR | All HTN patients | NR | NR | NR |
| Bezabih (2020) | Poor patient outcomes | 3 | ARBs | Odds ratio | 1.13 (0.95-1.35) | NR | NR | NR | 5,539 | 5,539 | NR / NR / 1209 | NR | NR | All HTN patients | NR | NR | NR |
| Cai (2020) | In-hospital mortality | 9 | ACEIs/ARBs | Odds ratio | 1.06 (0.75-1.5) | 25.00% | NR | NR | 3,949 | 3,949 | NR / NR / 881 | NR | in-hospital | Patients with and without HTN | Mixture of patients with HTN, DM, CAD, CCF, CKD, CVA, RD | NR | NR |
| Caldeira (2020) | Risk of severe disease associated with acei/ARB among patients with COVID-19 | 17 | ACEis/ARBs | Odds ratio | 0.9 (0.74-1.11) | 55% | 0.88 (0.63-1.22) | 68% | NR | NR | NR / NR / NR | NR | NR | NR | NR | NR | NR |
| Caldeira (2020) | All-cause mortality | 17 | ACEis/ARBs | Odds ratio | 0.91 (0.74-1.11) | 20% | 0.9 (0.68-1.18) | 27% | NR | NR | NR / NR / NR | NR | NR | NR | NR | NR | NR |
| Caldeira (2020) | COVID-19 infection risk | 6 | ACEis/ARBs | Odds ratio | 0.99 (0.91-1.11) | 36% | 0.99 (0.89-1.11) | 36% | NR | NR | NR / NR / NR | NR | NR | NR | NR | NR | NR |
| Caldeira (2020) | All-cause mortality | 3 | ARBs | Odds ratio | 0.8 (0.47-1.35) | 0% | NR | NR | NR | NR | NR / NR / NR | NR | NR | NR | NR | NR | NR |
| Caldeira (2020) | All-cause mortality | 4 | ACEis | Odds ratio | 0.85 (0.4-1.78) | 0% | NR | NR | NR | NR | NR / NR / NR | NR | NR | NR | NR | NR | NR |
| Caldeira (2020) | Risk of severe disease associated with acei/ARB compared with population controls | 2 | ACEis | Odds ratio | 0.91 (0.72-1.14) | 0% | NR | NR | NR | NR | NR / NR / NR | NR | NR | NR | NR | NR | NR |
| Caldeira (2020) | COVID-19 infection risk | 7 | ACEis | Odds ratio | 0.94 (0.87-1.02) | 0% | NR | NR | NR | NR | NR / NR / NR | NR | NR | NR | NR | NR | NR |
| Caldeira (2020) | Risk of severe disease associated with acei/ARB compared with population controls | 2 | ARBs | Odds ratio | 1.01 (0.67-1.5) | 69% | NR | NR | NR | NR | NR / NR / NR | NR | NR | NR | NR | NR | NR |
| Caldeira (2020) | COVID-19 infection risk | 6 | ARBs | Odds ratio | 1.01 (0.93-1.1) | 11% | NR | NR | NR | NR | NR / NR / NR | NR | NR | NR | NR | NR | NR |
| Caldeira (2020) | Risk of severe disease associated with acei/ARB among patients with COVID-19 | 4 | ACEis | Odds ratio | 1.05 (0.64-1.7) | 63% | NR | NR | NR | NR | NR / NR / NR | NR | NR | NR | NR | NR | NR |
| Caldeira (2020) | Risk of severe disease associated with acei/ARB compared with population controls | 1 | ACEis/ARBs | Odds ratio | 1.08 (0.79-1.47) | n/a | NR | NR | NR | NR | NR / NR / NR | NR | NR | NR | NR | NR | NR |
| Caldeira (2020) | Risk of severe disease associated with acei/ARB among patients with COVID-19 | 6 | ARBs | Odds ratio | 1.32 (0.75-2.3) | 86% | NR | NR | NR | NR | NR / NR / NR | NR | NR | NR | NR | NR | NR |
| Chan (2020) | Positive SARS-cov-2 infection | 7 | ACEis | Odds ratio | 1.21 (1.01-1.45) | >75% | 0.95 (0.86-1.05) | 34.70% | 164,084 | 22,114 | 14921 / 22685 / NR | NR | NR | NR | NR | NR | NR |
| Chan (2020) | Severe infection or mortality | 4 | ARBs | Odds ratio | NR | NR | 0.99 (0.83-1.18) | 18.50% | NR | NR | NR / NR / NR | NR | NR | NR | NR | NR | NR |
| Chan (2020) | Severe infection or mortality | 4 | ACEis | Odds ratio | NR | NR | 1 (0.8-1.26) | 51.60% | NR | NR | NR / NR / NR | NR | NR | NR | NR | NR | NR |
| Chan (2020) | Positive SARS-cov-2 infection | 7 | ARBs | Odds ratio | 1.25 (0.99-1.57) | >75% | 1.05 (0.97-1.14) | 34.90% | NR | NR | NR / NR / NR | NR | NR | NR | NR | NR | NR |
| Chu (2020) | All-cause mortality | 34 | ACEIs/ARBs | Odds ratio | 0.76 (0.59-0.99) | 88.0% | 0.81 (0.65-0.99) | 73.10% | 67,644 | 67,644 | NR / NR / NR | NR | NR | NR | NR | NR | NR |
| Chu (2020) | COVID-19 related severe adverse clinical outcomes | 40 | ACEis/ARBs | Odds ratio | 0.89 (0.78-1.01) | 71.0% | 0.81 (0.65-0.99) | 73.10% | 78,960 | 78,960 (?) | NR / NR / NR | NR | NR | NR | NR | NR | NR |
| Chu (2020) | COVID-19 infection risk | 11 | ACEis/ARBs | Odds ratio | 1.04 (0.94-1.14) | 71% | 0.87 (0.77-0.98) | 80.80% | 143,696 | NR | NR / NR / NR | detection of SARS-CoV-2 RNA in the patient's upper or lower respiratory tract system | NR | NR | NR | NR | NR |
| Chu (2020) | COVID-19 infection risk | 11 | ACEIs | Odds ratio | 0.87 (0.78-0.97) | 74% | NR | NR | NR | NR | NR / NR / NR | NR | NR | NR | NR | NR | NR |
| Chu (2020) | COVID-19 related severe adverse clinical outcomes | 40 | ARBs | Odds ratio | 0.93 (0.82-1.05) | 59.4% | NR | NR | NR | NR | NR / NR / NR | NR | NR | NR | NR | NR | NR |
| Chu (2020) | COVID-19 related severe adverse clinical outcomes | 40 | ACEis | Odds ratio | 0.95 (0.85-1.06) | 53.0% | NR | NR | NR | NR | NR / NR / NR | NR | NR | NR | NR | NR | NR |
| [de Almeide-Pititto (2020)](https://doi.org/10.1186/s13098-020-00586-4) | Severity of COVID-19 infection | 9 | ACEIs/ARBs | Odds ratio | 0.76 (0.39-1.49) | 95.8% | NR | NR | 10,900 | 10,900 | NR / NR / 4765 | NR | NR | Presence of diabetes mellitus, HTN, or CV disease | NR | NR | NR |
| Di Castelnuovo (2020) | Mortality or severe illness | 12 | ACEIs/ARBs | Odds ratio | 0.9 (0.8-1.01) | 5% | NR | NR | NR | NR | NR / NR / NR | NR | NR | HTN COVID-19 patients | NR | NR | NR |
| Di Castelnuovo (2020) | Mortality or severe illness | 9 | ACEIs/ARBs | Odds ratio | 1.18 (0.96-1.46) | 78% | NR | NR | NR | NR | NR / NR / NR | NR | NR | COVID-19 patients | NR | NR | NR |
| Diaz-Arocutipa (2020) | COVID-19 severity | 18 | ACEis/ARBs | Odds ratio | 0.79 (0.59-1.07) | 77% | 0.56 (0.37-0.87) | 0% | 11,870 | 11,870 | NR / NR / 5431 | diagnosed by diagnosed by RT-PCR | NR | COVID-19 patients (≥18 years) | NR | NR | NR |
| Diaz-Arocutipa (2020) | COVID-19 severity | NR | ACEis | Odds ratio | 1.1 (0.55-2.18) | 77% | 0.66 (0.37-1.18) | 61% | NR | NR | NR / NR / NR | NR | NR | NR | NR | NR | NR |
| Diaz-Arocutipa (2020) | All-cause mortality | 22 | ACEis/ARBs | Odds ratio | 1.11 (0.77-1.6) | 87% | 0.83 (0.49-1.38) | 53% | 23,059 | 23,059 | NR / NR / 4671 | diagnosed by diagnosed by RT-PCR | NR | COVID-19 patients (≥18 years) | NR | NR | NR |
| Diaz-Arocutipa (2020) | COVID-19 severity | NR | ARBs | Odds ratio | 1 (0.77-1.29) | 77% | 0.97 (0.79-1.2) | 0% | NR | NR | NR / NR / NR | NR | NR | NR | NR | NR | NR |
| Diaz-Arocutipa (2020) | All-cause mortality | NR | ACEis | Odds ratio | 1.18 (0.83-1.66) | 87% | 0.97 (0.83-1.13) | 0% | NR | NR | NR / NR / NR | NR | NR | NR | NR | NR | NR |
| Diaz-Arocutipa (2020) | All-cause mortality | NR | ARBs | Odds ratio | 1.79 (1.07-3) | 87% | 1.14 (0.98-1.34) | 0% | NR | NR | NR / NR / NR | NR | NR | NR | NR | NR | NR |
| Diaz-Arocutipa (2020) | Length of hospital stay | 5 | ACEIs/ARBs | Other - Mean Difference (days) | -0.96 (-2.50-0.57) | NR | NR | NR | NR | NR | NR / NR / NR | diagnosed by RT-PCR | NR | COVID-19 patients (≥18 years) | NR | NR | NR |
| Diaz-Arocutipa (2020) | ICU admission | NR | ACEis | Odds ratio | 1.16 (0.72-1.86) | NR | NR | NR | NR | NR | NR / NR / NR | NR | NR | NR | NR | NR | NR |
| Diaz-Arocutipa (2020) | ICU admission | NR | ARBs | Odds ratio | 1.26 (0.87-1.83) | NR | NR | NR | NR | NR | NR / NR / NR | NR | NR | NR | NR | NR | NR |
| Diaz-Arocutipa (2020) | Mechanical ventilation | 7 | ACEIs/ARBs | Odds ratio | 1.39 (0.99-1.94) | NR | NR | NR | 6,533 | 6,533 | NR / NR / NR | diagnosed by RT-PCR | NR | COVID-19 patients (≥18 years) | NR | NR | NR |
| Diaz-Arocutipa (2020) | ICU admission | 6 | ACEis/ARBs | Odds ratio | 1.45 (1.17-1.8) | NR | NR | NR | 8,884 | 8,884 | NR / NR / NR | diagnosed by RT-PCR | NR | COVID-19 patients (≥18 years) | NR | NR | NR |
| Diaz-Arocutipa (2020) | Hospitalis-ation | NR | ARBs | Odds ratio | 1.48 (0.95-2.31) | NR | NR | NR | NR | NR | NR / NR / NR | NR | NR | NR | NR | NR | NR |
| Diaz-Arocutipa (2020) | Hospitalis-ation | NR | ACEis | Odds ratio | 1.63 (0.94-2.83) | NR | NR | NR | NR | NR | NR / NR / NR | NR | NR | NR | NR | NR | NR |
| Diaz-Arocutipa (2020) | Hospitali-sation | 4 | ACEis/ARBs | Odds ratio | 1.83 (0.95-3.52) | NR | NR | NR | 5,048 | 5,048 | NR / NR / NR | diagnosed by RT-PCR | NR | COVID-19 patients (≥18 years) | NR | NR | NR |
| Diaz-Arocutipa (2020) | Hospital discharge | 3 | ACEIs/ARBs | Odds ratio | 2.27 (0.96-5.35) | NR | NR | NR | 301 | 301 | NR / NR / NR | diagnosed by diagnosed by RT-PCR | NR | COVID-19 patients (≥18 years) | NR | NR | NR |
| Flacco (2020) | Lethal COVID-19 | 4 | ACEIs/ARBs | Odds ratio | 0.88 (0.68-1.14) | 24% | NR | NR | 2,412 | 2,412 | NR / NR / 921 | laboratory confirmed PCR assay of nasal or pharyngeal swab specimens | NR | All HTN patients | NR | NR | NR |
| Flacco (2020) | Severe / lethal COVID-19 | 5 | ACEIs | Odds ratio | 0.9 (0.65-1.26) | 80% | NR | NR | 6,083 | 6,083 | 2002 / NR / NR | laboratory confirmed PCR assay of nasal or pharyngeal swab specimens | NR | All HTN patients | NR | NR | NR |
| Flacco (2020) | Severe/lethal COVID-19 | 5 | ARBs | Odds ratio | 0.92 (0.75-1.12) | 25% | NR | NR | 6,130 | 6,130 | NR / 1786 / NR | laboratory confirmed PCR assay of nasal or pharyngeal swab specimens | NR | All HTN patients | NR | NR | NR |
| Flacco (2020) | Severe / lethal COVID-19 | 5 | ACEIs/ARBs | Odds ratio | 1 (0.84-1.18) | 50% | NR | NR | 17,345 | 17,345 | NR / NR / 7465 | laboratory confirmed PCR assay of nasal or pharyngeal swab specimens | NR | All HTN patients | NR | NR | NR |
| Garg (2020) | In-hospital mortality | 6 | ACEIs/ARBs | Odds ratio | 0.64 (0.45-0.89) | 1% | NR | NR | 1,895 | 1,895 | NR / NR / 493 | NR | NR | COVID-19 patients with HTN | NR | NR | NR |
| Garg (2020) | Severe / critical illness | 6 | ACEIs/ARBs | Odds ratio | 0.76 (0.52-1.12) | 59% | NR | NR | 3,776 | 3,776 | NR / NR / 1415 | NR | NR | COVID-19 patients with HTN | NR | NR | NR |
| Garg (2020) | In-hospital mortality | 10 | ACEIs/ARBs | Odds ratio | 1.03 (0.69-1.55) | 65% | NR | NR | 13,149 | 13,149 | NR / NR / 2071 | NR | NR | COVID-19 patients | NR | NR | NR |
| Garg (2020) | Severe / critical illness | 11 | ACEIs/ARBs | Odds ratio | 1.18 (0.91-1.54) | 77% | NR | NR | 14,190 | 14,190 | NR / NR / 5175 | NR | NR | COVID-19 patients | NR | NR | NR |
| Ghosal (2020) | Mortality | 4 | ACEIs/ARBs | Odds ratio | 0.571 (0.373-0.875) | 0% | NR | NR | 1,691 | 1,691 | NR / NR / 374 | NR | NR | COVID-19 patients | NR | NR | NR |
| Ghosal (2020) | Severity of infection | 6 | ACEIs/ARBs | Odds ratio | 0.622 (0.314-1.233) | 70.4% | NR | NR | 1,942 | 1,942 | NR / NR / 423 | NR | NR | COVID-19 patients | NR | NR | NR |
| Ghosal (2020) | Hospitalis-ation | 2 | ACEIs/ARBs | Odds ratio | 0.81 (0.422-1.553) | 0% | NR | NR | 201 | 201 | NR / NR / 71 | NR | NR | COVID-19 patients | NR | NR | NR |
| Greco (2020) | Severe disease | 9 | ACEIs/ARBs | Odds ratio | 0.88 (0.6-1.31) | NR | NR | NR | NR | NR | NR / NR / NR | NR | NR | NR | NR | NR | NR |
| Greco (2020) | All-cause death | 13 | ACEIs/ARBs | Odds ratio | 0.95 (0.57-1.58) | 76.00% | NR | NR | 10,014 | 10,014 | NR / NR / 1839 | NR | NR | NR | NR | NR | NR |
| Grover (2020) | Clinical severity | 4 | ACEIs/ARBs | Odds ratio | 0.806 (0.412-1.579) | 50.52% | NR | NR | 2322 | 2322 | NR / NR / 208 | NR | NR | NR | Mixture of patients with HTN, CHD, HT, AV block, CV disease, HF; CKD, cerebrovascular disease, RD, kidney disease | NR | NR |
| Grover (2020) | Mortality | 6 | ACEIs/ARBs | Odds ratio | 0.863 (0.531-1.405) | 79.12% | NR | NR | 16645 | 16645 | NR / NR / 1933 | NR | NR | NR | Mixture of patients with HTN, CHD, HT, AV block, CAD, HF, asthma, COPD, CKD, end-stage renal disease, cerebrovascular disease, COPD, RD, kidney disease | NR | NR |
| Guo (2020) | Mortality | 6 | ACEIs/ARBs | Odds ratio | 0.57 (0.38-0.84) | 0.00% | NR | NR | 1,786 | 1,786 | NR / NR / 400 | NR | NR | All HTN patients | NR | NR | NR |
| Guo (2020) | Disease severity | 6 | ACEIs/ARBs | Odds ratio | 0.71 (0.46-1.08) | 59.00% | NR | NR | 2604 | 3780 | NR / NR / 1247 | NR | NR | All HTN patients | NR | NR | NR |
| Hasan (2020) | Mortality estimates | 12 | ACEIs/ARBs | Odds ratio | NR | NR | 0.73 (0.56-0.95) | NR | 18749 | 18749 | NR / NR / NR | NR | NR | NR | NR | NR | NR |
| Hasan (2020) | Severe/critical illness | 13 | ACEIs/ARBs | Odds ratio | NR | NR | 0.91 (0.75-1.1) | 66.00% | 7446 | 7446 | NR / NR / NR | NR | NR | NR | NR | NR | NR |
| Kahsour (2020) | Mortality | 6 | ACEIs/ARBs | Pooled adjusted relative risk | NR | NR | 0.63 (0.42-0.94) | 65.00% | 9573 | 9573 | NR / NR / 2266 | NR | NR | NR | NR | NR | NR |
| Kahsour (2020) | Mortality | 3 | ACEIs | Pooled adjusted relative risk | NR | NR | 0.78 (0.58-1.04) | 0.00% | 5226 | 5226 | NR / NR / NR | NR | NR | NR | NR | NR | NR |
| Kahsour (2020) | Mortality | 3 | ARBs | Pooled adjusted relative risk | NR | NR | 0.97 (0.73-1.3) | 0.00% | 5226 | 5226 | NR / NR / NR | NR | NR | NR | NR | NR | NR |
| Kerneis (2020) | All-cause mortality | 21 | ACEIs/ARBs | Odds ratio | 1 (0.69-1.45) | 84.00% | NR | NR | 11539 | 11539 | NR / NR / 3419 | NR | NR | NR | NR | NR | NR |
| Koshy (2020) | Risk of mortality or severe illness | 6 | ACEIs/ARBs | Odds ratio | 0.89 (0.73-1.07) | 38.00% | NR | NR | NR | NR | NR / NR / NR | NR | NR | NR | NR | NR | NR |
| Koshy (2020) | Risk of mortality or severe illness | 4 | ARBs | Odds ratio | 0.93 (0.79-1.1) | 0.00% | NR | NR | NR | NR | NR / NR / NR | NR | NR | NR | NR | NR | NR |
| Koshy (2020) | Likelihood of a positive COVID-19 test | 3 | ACEIs | Odds ratio | 0.93 (0.86-1.02) | 0.00% | NR | NR | NR | NR | NR / NR / NR | NR | NR | NR | NR | NR | NR |
| Koshy (2020) | Risk of mortality or severe illness | 4 | ACEIs | Odds ratio | 0.94 (0.79-1.11) | 0.00% | NR | NR | NR | NR | NR / NR / NR | NR | NR | NR | NR | NR | NR |
| Koshy (2020) | Likelihood of a positive COVID-19 test | 3 | ACEIs/ARBs | Odds ratio | 0.97 (0.97-1.05) | 0.00% | NR | NR | 68097 | 68097 | NR / NR / NR | NR | NR | NR | NR | NR | NR |
| Koshy (2020) | Likelihood of a positive COVID-19 test | 3 | ARBs | Odds ratio | 1.01 (0.91-1.12) | 56.00% | NR | NR | NR | NR | NR / NR / NR | NR | NR | NR | NR | NR | NR |
| Kurdi (2020) | Severe pneumonia | 2 | ACEIs/ARBs | Odds ratio | 1.285 (0.237-6.958) | 57.50% | 0.41 (0.05-3.275) | NR | 838 | 838 | NR / NR / 44 | NR | NR | NR | NR | NR | NR |
| Kurdi (2020) | Severe COVID-19 | 6 | ACEIs/ARBs | Odds ratio | 0.782 (0.529-1.154) | 43.30% | 0.48 (0.108-2.13) | NR | 14610 | 14610 | NR / NR / 2509 | NR | NR | NR | NR | NR | NR |
| Kurdi (2020) | Death/ICU admission | 3 | ACEIs/ARBs | Odds ratio | 0.67 (0.524-0.857) | 0.00% | 0.63 (0.471-0.842) | NR | 3635 | 3635 | NR / NR / 1116 | NR | NR | NR | NR | NR | NR |
| Kurdi (2020) | Mortality | 11 | ACEIs/ARBs | Odds ratio | 0.973 (0.746-1.269) | 6550.00% | 0.973 (0.26-1.66) | NR | 16252 | 16252 | NR / NR / 1722 | NR | NR | NR | NR | NR | NR |
| Kurdi (2020) | Hospitalisation | 4 | ARBs | Odds ratio | 0.907 (0.74-1.112) | 0.00% | 1 (0.702-1.424) | NR | 4579 | 4579 | NR / NR / NR | NR | NR | NR | NR | NR | NR |
| Kurdi (2020) | Hospitalisation | 4 | ACEIs | Odds ratio | 1.077 (0.791-1.465) | 63.70% | 1.17 (0.9-1.52) | NR | 4579 | 4579 | NR / NR / NR | NR | NR | NR | NR | NR | NR |
| Kurdi (2020) | COVID-19 infection risk | 3 | ACEIs | Odds ratio | 1.133 (0.91-1.42) | 0.00% | 1.18 (0.867-1.605) | NR | 2215 | NR | NR / NR / NR | NR | NR | NR | NR | NR | NR |
| Kurdi (2020) | COVID-19 infection risk | 7 | ACEIs/ARBs | Odds ratio | 1.014 (0.935-1.099) | 0.00% | 1.19 (0.962-1.473) | NR | 35178 | NR | NR / NR / 3301 | NR | NR | NR | NR | NR | NR |
| Kurdi (2020) | COVID-19 infection risk | 2 | ARBs | Odds ratio | 0.557 (0.107-2.895) | 97.90% | 1.29 (0.93-1.79) | NR | 1927 | NR | NR / NR / NR | NR | NR | NR | NR | NR | NR |
| Kurdi (2020) | Hospitalis-ation | 5 | ACEIs/ARBs | Odds ratio | 1.143 (0.806-1.65) | 74.50% | 1.3 (1.113-1.518) | NR | 23587 | 23587 | NR / NR / 1395 | NR | NR | NR | NR | NR | NR |
| Kurdi (2020) | Severe COVID-19 | 3 | ARBs | Odds ratio | 0.506 (0.247-1.036) | 18.00% | NR | NR | 1252 | 1252 | NR / NR / NR | NR | NR | NR | NR | NR | NR |
| Kurdi (2020) | Severe COVID-19 | 3 | ACEIs | Odds ratio | 0.718 (0.264-1.955) | 0.00% | NR | NR | 1252 | 1252 | NR / NR / NR | NR | NR | NR | NR | NR | NR |
| Kurdi (2020) | Death / ICU admission | 2 | ARBs | Odds ratio | 0.83 (0.65-1.061) | 0.00% | NR | NR | 7875 | 7875 | NR / NR / NR | NR | NR | NR | NR | NR | NR |
| Kurdi (2020) | Death / ICU admission | 2 | ACEIs | Odds ratio | 0.888 (0.694-1.136) | 0.00% | NR | NR | 7875 | 7875 | NR / NR / NR | NR | NR | NR | NR | NR | NR |
| Kurdi (2020) | ICU admission | 2 | ACEIs | Odds ratio | 0.945 (0.65-1.376) | 4.90% | NR | NR | 699 | 699 | NR / NR / NR | NR | NR | NR | NR | NR | NR |
| Kurdi (2020) | Ventilator use | 2 | ARBs | Odds ratio | 0.985 (0.084-11.57) | 88.60% | NR | NR | 411 | 411 | NR / NR / NR | NR | NR | NR | NR | NR | NR |
| Kurdi (2020) | Ventilator use | 2 | ACEIs | Odds ratio | 1.014 (0.03-34.758) | 64.70% | NR | NR | 411 | 411 | NR / NR / NR | NR | NR | NR | NR | NR | NR |
| Kurdi (2020) | Mortality | 2 | ACEIs | Odds ratio | 1.049 (0.751-1.464) | 26.30% | NR | NR | 2674 | 2674 | NR / NR / NR | NR | NR | NR | NR | NR | NR |
| Kurdi (2020) | ICU admission | 6 | ACEIs/ARBs | Odds ratio | 1.086 (0.652-1.809) | 84.40% | NR | NR | 26830 | 26830 | NR / NR / 1371 | NR | NR | NR | NR | NR | NR |
| Kurdi (2020) | ICU / ventilator use | 3 | ACEIs | Odds ratio | 1.149 (0.554-2.382) | 75.20% | NR | NR | 1263 | 1263 | NR / NR / NR | NR | NR | NR | NR | NR | NR |
| Kurdi (2020) | Mortality | 2 | ARBs | Odds ratio | 1.181 (0.983-1.418) | 0.60% | NR | NR | 2674 | 2674 | NR / NR / NR | NR | NR | NR | NR | NR | NR |
| Kurdi (2020) | ICU / ventilator use | 7 | ACEIs/ARBs | Odds ratio | 1.225 (0.836-1.795) | 83.20% | NR | NR | 27394 | 27394 | NR / NR / 1387 | NR | NR | NR | NR | NR | NR |
| Kurdi (2020) | ICU / ventilator use | 3 | ARBs | Odds ratio | 1.467 (0.907-2.737) | 66.20% | NR | NR | 1263 | 1263 | NR / NR / NR | NR | NR | NR | NR | NR | NR |
| Kurdi (2020) | ICU admission | 2 | ARBs | Odds ratio | 1.49 (1.126-1.973) | 0.00% | NR | NR | 699 | 699 | NR / NR / NR | NR | NR | NR | NR | NR | NR |
| Kurdi (2020) | Ventilator use | 4 | ACEIs/ARBs | Odds ratio | 1.492 (0.804-2.77) | 80.70% | NR | NR | 25147 | 25147 | NR / NR / 750 | NR | NR | NR | NR | NR | NR |
| Lee (2020) | Case-fatality | 21 | ACEIs/ARBs | Odds ratio | 0.75 (0.61-0.92) | 63.70% | NR | NR | 18735 | 18735 | NR / NR / 9227 | NR | NR | All HTN patients | NR | NR | NR |
| Lee (2020) | ITU admission / death | 3 | ACEIs/ARBs | Odds ratio | 0.76 (0.47-1.23) | 68.20% | NR | NR | 1443 | 1443 | NR / NR / 1092 | NR | NR | All HTN patients | NR | NR | NR |
| Lee (2020) | Severe / critical covid-19 | 14 | ACEIs/ARBs | Odds ratio | 0.8 (0.58-1.1) | 58.70% | NR | NR | 2564 | 2564 | NR / NR / 878 | NR | NR | All HTN patients | NR | NR | NR |
| Lee (2020) | Severe/critical covid-19 | 5 | ARBs | Odds ratio | 0.82 (0.52-1.31) | 59.80% | NR | NR | 1228 | 1228 | NR / 349 / NR | NR | NR | All HTN patients | NR | NR | NR |
| Lee (2020) | ITU admission / death | 3 | ACEIs | Odds ratio | 0.84 (0.65-1.1) | 16.60% | NR | NR | 1443 | 1443 | 551 / NR / NR | NR | NR | All HTN patients | NR | NR | NR |
| Lee (2020) | Hospital admission | 6 | ACEIs/ARBs | Odds ratio | 0.9 (0.62-1.31) | 83.30% | NR | NR | 5925 | 5925 | NR / NR / 3734 | NR | NR | All HTN patients | NR | NR | NR |
| Lee (2020) | Case-fatality | 9 | ARBs | Odds ratio | 0.91 (0.71-1.17) | 37.90% | NR | NR | 11658 | 11658 | NR / 2959 / NR | NR | NR | All HTN patients | NR | NR | NR |
| Lee (2020) | ITU / ventilation/death | 13 | ACEIs/ARBs | Odds ratio | 0.91 (0.72-1.15) | 59.30% | NR | NR | 13163 | 13163 | NR / NR / 6948 | NR | NR | All HTN patients | NR | NR | NR |
| Lee (2020) | ITU admission | 5 | ACEIs | Odds ratio | 0.93 (0.52-1.65) | 71.10% | NR | NR | 2328 | 2328 | 375 / NR / NR | NR | NR | All HTN patients | NR | NR | NR |
| Lee (2020) | Hospital admission | 6 | ARBs | Odds ratio | 0.94 (0.68-1.29) | 76.20% | NR | NR | 19607 | 19607 | NR / 7297 / NR | NR | NR | All HTN patients | NR | NR | NR |
| Lee (2020) | Hospital admission | 6 | ACEIs | Odds ratio | 0.95 (0.69-1.3) | 74.40% | NR | NR | 20915 | 20915 | 9641 / NR / NR | NR | NR | All HTN patients | NR | NR | NR |
| Lee (2020) | ITU admission / death | 3 | ARBs | Odds ratio | 0.97 (0.77-1.22) | 0.00% | NR | NR | 1443 | 1443 | NR / 572 / NR | NR | NR | All HTN patients | NR | NR | NR |
| Lee (2020) | Case-fatality | 9 | ACEIs | Odds ratio | 0.97 (0.86-1.09) | 0.00% | NR | NR | 11481 | 11481 | 2889 / NR / NR | NR | NR | All HTN patients | NR | NR | NR |
| Lee (2020) | ITU / ventilation / death | 7 | ARBs | Odds ratio | 1.01 (0.89-1.14) | 6.00% | NR | NR | 24854 | 24854 | NR / 10191 / NR | NR | NR | All HTN patients | NR | NR | NR |
| Lee (2020) | Likelihood of a positive COVID-19 test | 3 | ARBs | Odds ratio | 1.03 (0.92-1.16) | 0.00% | NR | NR | 11774 | 3340 | NR / 2217 / NR | NR | NR | All HTN patients | NR | NR | NR |
| Lee (2020) | ITU admission | 13 | ACEIs/ARBs | Odds ratio | 1.06 (0.73-1.56) | 81.00% | NR | NR | 7218 | 7218 | NR / NR / 3773 | NR | NR | All HTN patients | NR | NR | NR |
| Lee (2020) | Likelihood of a positive COVID-19 test | 3 | ACEIs | Odds ratio | 1.06 (0.94-1.2) | 0.00% | NR | NR | 11774 | 3340 | 2228 / NR / NR | NR | NR | All HTN patients | NR | NR | NR |
| Lee (2020) | Likelihood of a positive COVID-19 test | 4 | ACEIs/ARBs | Odds ratio | 1.06 (0.99-1.14) | 0.00% | NR | NR | 60141 | 5487 | NR / NR / 35944 | NR | NR | All HTN patients | NR | NR | NR |
| Lee (2020) | ITU / ventilation / death | 7 | ACEIs | Odds ratio | 1.07 (0.95-1.21) | 6.50% | NR | NR | 26162 | 26162 | 10926 / NR / NR | NR | NR | All HTN patients | NR | NR | NR |
| Lee (2020) | Severe / critical covid-19 | 4 | ACEIs | Odds ratio | 1.1 (0.64-1.89) | 20.90% | NR | NR | 1228 | 1228 | 98 / NR / NR | NR | NR | All HTN patients | NR | NR | NR |
| Lee (2020) | ITU admission | 5 | ARBs | Odds ratio | 1.32 (0.97-1.78) | 14.50% | NR | NR | 2328 | 2328 | NR / 429 / NR | NR | NR | All HTN patients | NR | NR | NR |
| Lee (2021) | All-cause mortality | 11 | ACEIs/ARBs | Odds ratio | 0.52 (0.37-0.72) | 87.00% | NR | NR | 12239 | 12239 | NR / NR / 2385 | High-throughput sequencing or real-time RT-PCR assay using upper or lower respiratory tract specimens | NR | NR | NR | NR | NR |
| Lee (2021) | Severe disease | 13 | ACEIs/ARBs | Odds ratio | 0.68 (0.44-1.07) | 95.00% | NR | NR | 12848 | 12848 | NR / NR / 4658 | High-throughput sequencing or real-time reverse RT-PCR assay using upper or lower respiratory tract specimens | NR | NR | NR | NR | NR |
| Liu (2020) | Mortality | 7 | ACEIs/ARBs | Odds ratio | 0.52 (0.35-0.79) | 46% | NR | NR | 11,509 | 11,509 | NR / NR / NR | NR | NR | COVID-19 patients | NR | NR | NR |
| Liu (2020) | Disease severity | 8 | ACEIs/ARBs | Odds ratio | 0.75 (0.59-0.96) | 54% | NR | NR | NR | NR | NR / NR / NR | NR | NR | NR | NR | NR | NR |
| Liu (2020) | COVID-19 infection risk | 2 | ACEIs/ARBs | Odds ratio | 0.95 (0.89-1.02) | 0% | NR | NR | 10,629 | NR | NR / NR / NR | NR | NR | NR | NR | NR | NR |
| Lo (2020) | Severe disease | 12 | ACEIs/ARBs | Odds ratio | 0.94 (0.59-1.5) | 89% | NR | NR | 8,068 | 8,068 | NR / NR / 2439 | NR | NR | Adults with confirmed COVID-19 | NR | NR | NR |
| Lo (2020) | Combined outcome | 21 | ACEIs/ARBs | Odds ratio | 1.09 (0.8-1.48) | 92% | NR | NR | 24,191 | 24,191 | NR / NR / 9027 | NR | NR | Adults with confirmed COVID-19 | NR | NR | NR |
| Lo (2020) | Mortality | 17 | ACEIs/ARBs | Odds ratio | 1.29 (0.89-1.87) | 91% | NR | NR | 21,603 | 21,603 | NR / NR / 7907 | NR | NR | Adults with confirmed COVID-19 | NR | NR | NR |
| Megaly (2020) | Severity of illness | 4 | ACEIs/ARBs | Odds ratio | 0.73 (0.24-2.24) | 63% | NR | NR | 3,267 | 3,267 | NR / NR / 534 | NR | NR | Patients with COVID-19 infection | NR | NR | NR |
| Megaly (2020) | All-cause death | 4 | ACEIs/ARBs | Odds ratio | 0.75 (0.36-1.57) | 62% | NR | NR | 3,267 | 3,267 | NR / NR / 534 | NR | NR | Patients with COVID-19 infection | NR | NR | NR |
| Megaly (2020) | Length of stay | 4 | ACEIs/ARBs | Other - Mean Difference (days) | 1.7 [days] (-0.7-4) | 92% | NR | NR | 3,267 | 3,267 | NR / NR / 534 | NR | NR | Patients with COVID-19 infection | NR | NR | NR |
| Nunes (2020) | Mortality | 5 | ACEIs | Odds ratio | 1.48 (1.019-2.15) | NR | NR | NR | 6,117 | 6,117 (?) | 944 / NR / NR | NR | NR | NR | NR | NR | NR |
| Patoulias (2020) | Development of severe or critial illness | 15 | ACEIs/ARBs | Odds ratio | 0.86 (0.64-1.16) | 90% | NR | NR | 20,256 | 20,256 | NR / NR / 8530 | NR | NR | NR | NR | NR | NR |
| Patoulias (2020) | COVID-19 infection risk | 11 | ACEIs/ARBs | Odds ratio | 0.99 (0.83-1.17) | 93% | NR | NR | 593,079 | 18,699 (?) | NR / NR / 95789 | NR | NR | NR | NR | NR | NR |
| Patoulias (2020) | SARS-cov-2-related death | 11 | ACEIs/ARBs | Odds ratio | 1.06 (0.77-1.47) | 83% | NR | NR | 15,668 | 15,668 | NR / NR / 5258 | NR | NR | NR | NR | NR | NR |
| Patoulias (2020) | ICU admission | 6 | ACEIs/ARBs | Odds ratio | 1.4 (0.8-2.43) | 86% | NR | NR | 4,560 | 4,560 | NR / NR / 1110 | NR | NR | NR | NR | NR | NR |
| Patoulias (2020) | Hospital admission | 5 | ACEIs/ARBs | Odds ratio | 1.74 (0.95-3.17) | 96% | NR | NR | 7,678 | 7,678 (?) | NR / NR / 2810 | NR | NR | NR | NR | NR | NR |
| Pirola (2020) | Critical or fatal versus on-critical disease | 16 | ACEIs/ARBs | Odds ratio | 0.768 (0.651-0.907) | 63% | NR | NR | 24,676 | 24,676 | NR / NR / 24676 | NR | NR | Hospital-ised patients with confirmed COVID-19 infection and arterial HTN prior to hospitalis-ation | NR | NR | NR |
| Pranata (2020) | Mortality in patients with COVID-19 pneumonia | 11 | ACEIs/ARBs | Odds ratio | 0.73 (0.38-1.4) | 81% | 0.83 (0.54-1.27) | 0% | 4,964 | 4,964 | NR / NR / 1655 | NR | NR | COVID-19 patients with HTN | NR | NR | NR |
| Pranata (2020) | Severe COVID-19 | 9 | ACEIs/ARBs | Odds ratio | 1.03 (0.73-1.45) | 65% | NR | NR | 3,547 | 3,547 | NR / NR / 1485 | NR | NR | COVID-19 patients with HTN | NR | NR | NR |
| Qu (2020) | COVID-19 infection risk | 7 | ARBs | Odds ratio | 1.07 (0.76-1.5) | 96.0% | 0.73 (0.49-1.08) | 95% | NR | NR | NR / NR / NR | NR | NR | NR | NR | NR | NR |
| Qu (2020) | Severity of COVID-19 infection | 5 | ACEiS | Odds ratio | 1.01 (0.63-1.6) | 75.0% | 0.9 (0.72-1.14) | 0% | NR | NR | NR / NR / NR | NR | NR | NR | NR | NR | NR |
| Qu (2020) | Severity of COVID-19 infection | 5 | ACEis/ARBs | Odds ratio | 0.86 (0.57-1.31) | 86.0% | 0.9 (0.77-1.05) | 27.60% | NR | NR | NR / NR / NR | NR | NR | Patients diagnosed with COVID-19 | NR | NR | NR |
| Qu (2020) | Severity of COVID-19 infection | 10 | ARBs | Odds ratio | 0.75 (0.41-1.39) | 92.0% | 0.91 (0.74-1.13) | 46% | NR | NR | NR / NR / NR | NR | NR | NR | NR | NR | NR |
| Qu (2020) | COVID-19 infection risk | 4 | ACEiS | Odds ratio | 1.27 (0.95-1.69) | 92.0% | 0.94 (0.86-1.01) | 0% | NR | NR | NR / NR / NR | NR | NR | NR | NR | NR | NR |
| Qu (2020) | COVID-19 infection risk | 4 | ACEis/ARBs | Odds ratio | 1.1 (0.84-1.43) | 89.0% | 0.96 (0.91-1.01) | 6% | NR | NR | NR / NR / NR | NR | NR | Patients diagnosed with COVID-19 | NR | NR | NR |
| Qu (2020) | Risk of hospital admission | 2 | ACEiS | Odds ratio | NR | NR | 1.29 (1.07-1.57) | 62% | NR | NR | NR / NR / NR | NR | NR | NR | NR | NR | NR |
| Qu (2020) | Risk of hospital admission | 3 | ACEis/ARBs | Odds ratio | NR | NR | 1.38 (1.21-1.57) | 35% | NR | NR | NR / NR / NR | NR | NR | Patients diagnosed with COVID-19 | NR | NR | NR |
| Qu (2020) | Risk of hospital admission | 4 | ARBs | Odds ratio | NR | NR | 1.69 (1.46-1.96) | 0% | NR | NR | NR / NR / NR | NR | NR | NR | NR | NR | NR |
| Ren (2020) | Mortality | 14 | ACEIs/ARBs | Odds ratio | 0.77 (0.66-0.91) | 47% | NR | NR | NR | NR | NR / NR / NR | NR | NR | All HTN patients | NR | NR | NR |
| Ren (2020) | Severity | 10 | ACEIs/ARBs | Odds ratio | 0.81 (0.66-0.99) | 50.5% | NR | NR | NR | NR | NR / NR / NR | NR | NR | All HTN patients | NR | NR | NR |
| Ren (2020) | Mortality | 26 | ACEIs/ARBs | Odds ratio | 0.92 (0.74-1.13) | 84.7% | NR | NR | NR | NR | NR / NR / NR | NR | NR | NR | NR | NR | NR |
| Ren (2020) | Severity | 24 | ACEIs/ARBs | Odds ratio | 0.92 (0.77-1.11) | 83.1% | NR | NR | NR | NR | NR / NR / NR | NR | NR | NR | NR | NR | NR |
| Ren (2020) | Incidence | 13 | ACEIs/ARBs | Odds ratio | 0.96 (0.86-1.08) | 88.4% | NR | NR | NR | NR | NR / NR / NR | NR | NR | NR | NR | NR | NR |
| Ren (2020) | Incidence | 3 | ACEIs/ARBs | Odds ratio | 0.97 (0.89-1.06) | 33.6% | NR | NR | NR | NR | NR / NR / NR | NR | NR | All HTN patients | NR | NR | NR |
| Ren (2020) | Hospitalisation | 13 | ACEIs/ARBs | Odds ratio | 1.09 (0.91-1.31) | 70.3% | NR | NR | NR | NR | NR / NR / NR | NR | NR | NR | NR | NR | NR |
| Ren (2020) | ICU admission | 8 | ACEIs/ARBs | Odds ratio | 1.19 (0.85-1.66) | 56.6% | NR | NR | NR | NR | NR / NR / NR | NR | NR | NR | NR | NR | NR |
| Ssen-tongo (2020) | Mortality in patients with HTN hospitalised for COVID-19 | 7 | ACEIs/ARBs | Risk ratio | 0.65 (0.45-0.94) | 80% | NR | NR | NR | NR | NR / NR / NR | NR | NR | HTN patients hospitalised with COVID-19 | NR | NR | NR |
| Ssen-tongo (2020) | Mortality in patients hospitalised for COVID-19 | 12 | ACEIs/ARBs | Risk ratio | 0.77 (0.63-0.95) | NR | NR | NR | NR | NR | NR / NR / NR | NR | NR | Patients hospitalised with COVID-19 | NR | NR | NR |
| Tleyjeh (2020) | COVID-19 infection risk | 9 | ACEIs | Odds ratio | 0.98 (0.91-1.05) | 15% | NR | NR | NR | NR | NR / NR / NR | NR | NR | NR | NR | NR | NR |
| Tleyjeh (2020) | COVID-19 infection risk | 8 | ARBs | Odds ratio | 1.04 (0.98-1.1) | 0% | NR | NR | NR | NR | NR / NR / NR | NR | NR | NR | NR | NR | NR |
| Usman (2020) | Risk of mortality | 5 | ACEIs/ARBs | Odds ratio | 0.74 (0.34-1.58) | 65.00% | NR | NR | NR | NR | NR / NR / NR | NR | NR | All HTN patients | NR | NR | NR |
| Usman (2020) | COVID-19 infection risk | 3 | ACEIs | Odds ratio | 0.96 (0.88-1.04) | 0.00% | NR | NR | NR | NR | NR / NR / NR | NR | NR | All HTN patients | NR | NR | NR |
| Usman (2020) | COVID-19 infection risk | 3 | ARBs | Odds ratio | 0.99 (0.91-1.08) | 5.00% | NR | NR | NR | NR | NR / NR / NR | NR | NR | All HTN patients | NR | NR | NR |
| Wang (2020) | Risk of mortality | 26 | ACEIs/ARBs | Odds ratio | 0.624 (0.457-0.852) | 74% | NR | NR | 16,307 | 16,307 | NR / NR / 8104 | NR | NR | All HTN patients | NR | 66.7 (mean) | 47.2% |
| Wang (2020) | Clinical indicators (risks of severe COVID-19, ICU, requiring ventilatory support) | 7 | ACEIs/ARBs | Odds ratio | 0.628 (0.475-1.978) | 0% | NR | NR | 1,734 | 1,734 | NR / NR / NR | NR | NR | NR | NR | NR | NR |
| Wang (2020) | Clinical indicators (risks of severe COVID-19, ICU, requiring ventilatory support) | 10 | ACEIs/ARBs | Odds ratio | 0.699 (0.444-1.098) | 80% | NR | NR | 2,309 | 2,309 | NR / NR / NR | NR | NR | NR | NR | NR | NR |
| Wang (2020) | Clinical indicators (risks of severe COVID-19, ICU, requiring ventilatory support) | 11 | ACEIs/ARBs | Odds ratio | 0.704 (0.46-1.078) | 51.5% | NR | NR | 1,912 | 1,912 | NR / NR / NR | NR | NR | NR | NR | NR | NR |
| Xu (2020) | COVID-19 mortality | 13 | ACEIs/ARBs | Odds ratio | NR | NR | 0.87 (0.66-1.14) | 69.00% | UNCLEAR | UNCLEAR | UNCLEAR / UNCLEAR / UNCLEAR | NR | NR | NR | NR | NR | NR |
| Xu (2020) | Severe outcomes of COVID-19 | 24 | ACEIs/ARBs | Odds ratio | NR | NR | 0.95 (0.73-1.24) | 65.00% | UNCLEAR | UNCLEAR | UNCLEAR / UNCLEAR / UNCLEAR | NR | NR | NR | NR | NR | NR |
| Xu (2020) | COVID-19 infection risk | 13 | ACEIs/ARBs | Odds ratio | NR | NR | 1 (0.94-1.05) | 0.00% | UNCLEAR | UNCLEAR | UNCLEAR / UNCLEAR / UNCLEAR | diagnosis via RT-PCR | NR | NR | NR | NR | NR |
| Xue (2020) | Creatinine | 3 | ACEIs/ARBs | Other - SMD (Std. Mean Difference) | 0.22 (0.04-0.41) | 0.00% | NR | NR | 530 | 530 | NR / NR / 215 | NR | NR | All HTN patients | NR | NR | NR |
| Xue (2020) | D-dimer | 5 | ACEIs/ARBs | Other - SMD (Std. Mean Difference) | -0.22 (-0.36--0.06) | 26.00% | NR | NR | 680 | 680 | NR / NR / 266 | NR | NR | All HTN patients | NR | NR | NR |
| Xue (2020) | Fever | 5 | ACEIs/ARBs | Odds ratio | 0.74 (0.55-0.98) | 0.00% | NR | NR | 1446 | 1446 | NR / NR / 339 | NR | NR | All HTN patients | NR | NR | NR |
| Xue (2020) | Dry cough | 5 | ACEIs/ARBs | Odds ratio | 0.87 (0.66-1.14) | 0.00% | NR | NR | 1376 | 1376 | NR / NR / 269 | NR | NR | All HTN patients | NR | NR | NR |
| Yokojama (2020) | In-hospital mortality rate | 10 | ACEIs/ARBs | Risk ratio | 0.66 (0.49-0.89) | 30.00% | NR | NR | 7596 | NR | NR / NR / 1948 | NR | in hospital | All HTN patients | NR | NR | NR |
| Yokojama (2020) | In-hospital mortality rate | 14 | ACEIs/ARBs | Risk ratio | 0.88 (0.64-1.2) | 73.00% | NR | NR | 11134 | NR | NR / NR / 2647 | NR | in hospital | NR | NR | NR | NR |
| Yokojama (2020) | COVID-19 infection risk | 3 | ACEIs | Odds ratio | 0.96 (0.88-1.04) | 0.00% | NR | NR | 58887 | NR | UNCLEAR / NR / NR | NR | NR | NR | NR | NR | NR |
| Yokojama (2020) | COVID-19 infection risk | 3 | ARBs | Odds ratio | 0.99 (0.91-1.08) | 5.00% | NR | NR | 58887 | NR | NR / UNCLEAR / NR | NR | NR | NR | NR | NR | NR |
| Zhang G (2020) | Length of hospital stay | NR | ACEIs/ARBs | Other - SMD (Std. Mean Difference) | 0.05 (-0.16-0.26) | 84.43% | NR | NR | NR | NR | NR / NR / NR | NR | In-patients or out-patients | General population + those with HTN | NR | NR | NR |
| Zhang G (2020) | Risk of mortality | 21 | ACEIs/ARBs | Odds ratio | 0.65 (0.46-0.85) | 73.37% | NR | NR | 10692 | NR | NR / NR / 4538 | NR | In-patients or out-patients | General population + those with HTN | NR | NR | NR |
| Zhang G (2020) | Severity/mortality | NR | ACEIs/ARBs | Odds ratio | 0.69 (0.43-0.95) | 22.90% | NR | NR | NR | NR | NR / NR / NR | NR | In-patients or out-patients | General population + those with HTN | NR | NR | NR |
| Zhang G (2020) | Acute respiratory distress syndrome | NR | ACEIs/ARBs | Odds ratio | 0.71 (0.46-0.95) | 0.00% | NR | NR | NR | NR | NR / NR / NR | NR | In-patients or out-patients | General population + those with HTN | NR | NR | NR |
| Zhang G (2020) | Hospitalisation | NR | ACEIs/ARBs | Odds ratio | 0.79 (0.6-0.98) | 0.00% | NR | NR | NR | NR | NR / NR / NR | NR | In-patients or out-patients | General population + those with HTN | NR | NR | NR |
| Zhang G (2020) | Mechanical ventilation | NR | ACEIs/ARBs | Odds ratio | 0.89 (0.61-1.16) | 3.19% | NR | NR | NR | NR | NR / NR / NR | NR | In-patients or out-patients | General population + those with HTN | NR | NR | NR |
| Zhang G (2020) | COVID‐19 severity | NR | ACEIs/ARBs | Odds ratio | 0.89 (0.63-1.15) | 38.55% | NR | NR | NR | NR | NR / NR / NR | NR | In-patients or out-patients | General population + those with HTN | NR | NR | NR |
| Zhang G (2020) | Transfer to the ICU | 14 | ACEIs/ARBs | Odds ratio | 0.96 (0.56-1.37) | 88.31% | NR | NR | NR | NR | NR / NR / NR | NR | In-patients or out-patients | General population + those with HTN | NR | NR | NR |
| Zhang G (2020) | Dialysis | NR | ACEIs/ARBs | Odds ratio | 1.24 (0.09-2.39) | 0.00% | NR | NR | NR | NR | NR / NR / NR | NR | In-patients or out-patients | General population + those with HTN | NR | NR | NR |
| Zhang X (2020) | Mortality | 8 | ACEIs/ARBs | Odds ratio | 0.66 (0.38-1.12) | 82.20% | 0.91 (0.51-1.61) | 33.40% | 23525 | 23525 | NR / NR / 2171 | NR | NR | NR | NR | NR | NR |
| Zhang X (2020) | COVID-19 severity | 7 | ACEIs/ARBs | Odds ratio | 0.95 (0.83-1.1) | 57.60% | NR | NR | 6161 | 6161 | NR / NR / 4237 | NR | NR | NR | NR | NR | NR |
| Zhang X (2020) | COVID-19 infection risk | 3 | ACEIs | Odds ratio | 0.98 (0.92-1.04) | 0.00% | NR | NR | NR | NR | NR / NR / NR | NR | NR | NR | NR | NR | NR |
| Zhang X (2020) | COVID-19 infection risk | 3 | ACEIs/ARBs | Odds ratio | 0.99 (0.95-1.04) | 0.00% | NR | NR | NR | NR | NR / NR / NR | NR | NR | NR | NR | NR | NR |
| Zhang X (2020) | COVID-19 infection risk | 3 | ACEIs | Odds ratio | 1.01 (0.95-1.07) | 8.90% | NR | NR | NR | NR | NR / NR / NR | NR | NR | NR | NR | NR | NR |
| Zhang Y (2020) | COVID-19 infection risk | 18 | ACEIs/ARBs | Odds ratio | 1.05 (0.9-1.21) | NR | 0.93 (0.85-1.02) | NR | NR | NR | NR / NR / NR | positive test result from a real-time RT-PCR assay of naso-pharyngeal swab samples | NR | NR | NR | NR | NR |
| Zhang Y (2020) | COVID-19 severity | 29 | ACEIs/ARBs | Odds ratio | 1.05 (0.81-1.36) | 86.00% | NR | NR | NR | NR | NR / NR / NR | NR | NR | NR | NR | NR | NR |
| Zhang Y (2020) | Mortality | 30 | ACEIs/ARBs | Odds ratio | 1.12 (0.88-1.44) | 88.00% | NR | NR | NR | NR | NR / NR / NR | NR | NR | NR | NR | NR | NR |

ACEI = angiotensin-converting enzyme inhibitors; ARB = angiotensin receptor blockers; AV = atrioventricular; CAD = coronary artery disease; CCF = congestive cardiac failure; CHD = coronary heart disease; CKD = chronic kidney disease; COPD = chronic obstructive pulmonary disease; CV = cardiovascular; CVA = cerebrovascular accidents; DM = diabetes mellitus; HF = heart failure; HT = hypothyroidism; HTN: hypertension / hypertensive; ICU = intensive care unit; ITU = intensive therapy unit; NR = not reported; RAAS = renin-angiotensin-aldosterone system; RD = respiratory disease; RT-PCR = reverse-transcriptase-polymerase-chain-reaction
