## Supplementary file 5 for "An umbrella review and meta-analysis of the use of renin-angiotensin system drugs and COVID-19 outcomes: what do we know so far?"

**AMSTAR 2 Quality Assessment Results**

| **Review** | **Overall confidence in the results of the review** | **1. PICO** | **2.**  **A priori protocol*** | **3. Inclusion criteria** | **4. Comprehensive literature search strategy** | 5. Duplicate study selection | **6. Duplicate data extraction** | **7. Excluded studies** | **8. Adequate detail** | **9. Conducted risk of bias (RoB)*** | **10. Sources of funding** | **11. Appropriate methods*** | **12. Assess impact of RoB** | **13. Discuss RoB results*** | **14. Explain / discuss hetero-geneity** | **15. Publication bias*** |
| --- | --- | --- | --- | --- | --- | --- | --- | --- | --- | --- | --- | --- | --- | --- | --- | --- |
| Abdulha | Critically low | Y | N | Y | PY | Y | Y | N | PY | Y | N | Y | N | Y | Y | N |
| Alamer | Critically low | Y | N | N | N | N | N | N | N | N | N | Y | N | N | N | N |
| Asiimwe | Moderate | Y | Y | Y | Y | N | Y | N | Y | Y | N | Y | Y | Y | Y | Y |
| Baral | Low | Y | N | Y | PY | N | N | N | Y | Y | N | Y | Y | Y | Y | Y |
| Barochiner-a | Low | Y | N | Y | PY | N | Y | N | PY | Y | N | Y | Y | Y | Y | Y |
| Barochiner-b | Low | Y | N | Y | PY | Y | Y | N | PY | Y | Y | Y | Y | Y | N | Y |
| Beressa | Low | Y | Y | Y | Y | Y | Y | N | N | Y | N | Y | Y | N | N | Y |
| Bezabih | Critically low | Y | N | Y | PY | N | N | N | PY | Y | N | N | N | Y | N | Y |
| Cai | Critically low | Y | N | Y | Y | Y | Y | N | PY | N | N | Y | Y | Y | Y | N |
| Caldeira | Moderate | Y | Y | Y | PY | Y | N | Y | PY | Y | N | Y | Y | Y | Y | Y |
| Chan | Moderate | Y | Y | N | PY | Y | Y | N | PY | Y | N | Y | Y | Y | Y | Y |
| Chu | Low | Y | N | Y | PY | Y | Y | N | Y | Y | N | Y | Y | Y | Y | Y |
| de Almeide-Pititto | Low | Y | Y | N | PY | N | Y | N | N | Y | N | Y | N | N | Y | Y |
| Di Castelnuovo | Critically low | Y | N | Y | PY | Y | N | N | Y | N | N | Y | Y | N | Y | Y |
| Diaz-Arocutipa | Moderate | Y | Y | Y | PY | Y | Y | N | PY | Y | N | Y | Y | Y | Y | Y |
| Flacco | Low | Y | N | Y | PY | Y | Y | N | PY | Y | N | Y | Y | Y | Y | Y |
| Garg | Critically low | Y | N | Y | PY | N | N | N | PY | N | N | Y | Y | Y | N | N |
| Ghosal | Critically low | Y | N | Y | PY | Y | N | N | N | N | N | Y | N | Y | Y | Y |
| Greco | Low | Y | Y | Y | N | N | N | N | N | Y | N | Y | Y | N | N | Y |
| Grover | Critically low | Y | N | Y | PY | N | N | N | N | Y | N | Y | Y | N | N | Y |
| Guo | Critically low | Y | N | Y | PY | Y | N | N | N | Y | N | Y | N | N | N | Y |
| Hasan | Moderate | Y | PY | Y | Y | Y | N | N | PY | Y | N | Y | Y | Y | Y | Y |
| Kashour | Critically low | Y | N | Y | N | Y | Y | N | PY | Y | N | Y | Y | Y | N | N |
| Kerneis | Critically low | Y | N | Y | PY | Y | Y | N | PY | N | N | Y | N | N | Y | N |
| Koshy | Critically low | Y | N | Y | PY | N | N | N | PY | Y | N | Y | Y | N | Y | N |
| Kurdi | Moderate | Y | PY | Y | PY | Y | N | N | PY | Y | N | Y | Y | Y | N | Y |
| Lee | Moderate | Y | Y | Y | Y | Y | Y | N | PY | Y | N | Y | Y | Y | Y | Y |
| Lee 2021 | Moderate | Y | Y | Y | PY | Y | N | Y | PY | Y | N | Y | Y | Y | Y | Y |
| Liu | Critically low | Y | N | Y | PY | Y | Y | N | N | Y | N | Y | Y | Y | N | N |
| Lo | Low | Y | N | Y | PY | N | Y | Y | PY | Y | N | Y | Y | Y | Y | Y |
| Megaly | Critically low | Y | N | Y | PY | N | Y | N | Y | Y | N | Y | N | N | N | Y |
| Nunes | Critically low | Y | N | Y | PY | N | N | N | PY | N | N | Y | N | Y | Y | N |
| Patoulias | Critically low | Y | N | Y | PY | Y | Y | N | N | Y | N | Y | N | N | Y | Y |
| Pirola | Low | Y | N | N | PY | N | N | N | N | Y | N | Y | Y | Y | Y | Y |
| Pranata | Low | Y | N | Y | PY | N | Y | N | PY | Y | N | Y | Y | Y | Y | Y |
| Qu | Low | Y | N | N | PY | Y | N | N | PY | Y | N | Y | Y | Y | N | Y |
| Ren | Low | Y | N | Y | PY | Y | N | N | PY | Y | N | Y | Y | Y | N | Y |
| Ssentongo | Critically low | Y | Y | Y | PY | Y | Y | N | N | Y | N | Y | Y | N | N | N |
| Tleyjeh | Critically low | Y | N | Y | PY | N | Y | N | PY | Y | N | Y | Y | Y | N | N |
| Usman | Critically low | Y | N | Y | PY | N | N | N | N | N | N | Y | N | N | N | N |
| Wang | Low | Y | Y | N | PY | Y | Y | N | PY | Y | N | Y | Y | N | Y | Y |
| Xu | Moderate | Y | Y | Y | Y | Y | Y | N | PY | Y | N | Y | Y | Y | N | Y |
| Xue | Critically low | Y | N | Y | PY | N | Y | N | N | Y | N | Y | Y | N | N | Y |
| Yokoyama | Critically low | Y | N | N | PY | Y | N | N | N | N | N | N | Y | Y | N | N |
| Zhang G | Moderate | Y | Y | N | PY | Y | Y | N | PY | Y | N | Y | Y | Y | Y | Y |
| Zhang X | Critically low | Y | N | N | PY | N | Y | N | PY | Y | N | Y | Y | Y | Y | N |
| Zhang Y | Low | Y | N | Y | PY | Y | Y | N | PY | Y | N | Y | Y | Y | Y | Y |

*Critical AMSTAR tool domains; Y = Yes; PY = Partial Yes; N = No
