## Supplementary file 2 for "An umbrella review and meta-analysis of the use of renin-angiotensin system drugs and COVID-19 outcomes: what do we know so far?"

**Irrelevant and excluded articles**

Article deems irrelevant at abstract/title stage (n=84) **(1-84)**

Articles excluded at full text review (n=26) (85-110)

- Duplicate review* (n=17) (85, 87-89, 91, 92, 95, 97, 98, 100-103, 105-107, 109)
- No meta-analysis conducted (n=5) (90, 93, 94, 99, 110)
- Abstract / conference proceeding (n=2) (104, 108)
- No results available (n=1) (86)
- Insufficient information available for meta-analysis (n=1) (96)

** For example, identified as a pre-print of a published review*
